# Artificial Intelligence for Automated, Highly Accurate, and Scalable Multimodal EHR Data Abstraction

**DOI:** 10.64898/2026.03.16.26348522

**Authors:** Georgios Margaritis, Periklis Petridis, Dimitris Bertsimas, Jordan Bloom, Robert Hagberg, Robert Habib, David M. Shahian, Agni Orfanoudaki

## Abstract

Electronic health records (EHRs) contain rich multimodal data but remain underutilized for populating clinical registries due to the time and cost of manual abstraction. We developed an AI-driven pipeline to automate data abstraction for variables in the Society of Thoracic Surgeons Adult Cardiac Surgery Database (ACSD). Models were developed using Mass General Brigham data and externally validated on Hartford HealthCare data. The pipeline processes ten clinical EHR sources, seven unstructured text types and three structured data types; each encoded using two language-model embeddings and term frequency–inverse document frequency. This approach yielded 30 source-specific models per target variable whose predictions were aggregated by an ensemble meta-learner, followed by a dual-threshold confidence framework that enforced registry-grade high accuracy standards and deferred uncertain predictions to human review. The developed pipeline achieved an overall accuracy exceeding 99% across 647 registry variables, while automatically completing 49.5% and 43.2% of variables at both sites, respectively. These results demonstrate that AI-assisted abstraction can substantially reduce clinical registry data collection burden while maintaining high accuracy.

## 1 Introduction

Comprehensive healthcare data are essential for clinical research, hospital operations, billing, regulatory compliance, and for measuring and improving provider quality. Ideally, these data should be available for all or most patients, at least in specifically defined populations; accessible with minimum data collection burden; clearly specified so that data from different sources are comparable; clinically relevant so they have face validity with providers; granular, so they capture important nuances in patient presentation, treatment, and outcomes; and highly accurate so that stakeholders trust them.

The electronic health record (EHR) serves as the primary repository of inpatient and out-patient care data [1]. However, EHR data are typically underutilized due to the predominance of unstructured information [2, 3]. Their use in clinical research, hospital operations, and quality measurement or improvement is limited by the burden of manual data abstraction to translate these largely unstructured EHR data to a standardized structure that is comparable across thousands of hospitals. This manual effort demands significant institutional resources and diverts clinical professionals from direct patient care [4]. This bottleneck limits scalability and hampers efforts to build learning health systems and equitable digital infrastructures.

A prime illustration of this challenge is the development and maintenance of national clinical registries. Disease- and procedure-specific national clinical registries are valuable data sources for healthcare quality measurement and improvement, public reporting for accountability and transparency, provider oversight by regulators and payers, clinical research, and policymaking [5]. These registries are curated by content experts as structured, standardized repositories, developed and published by healthcare professional organizations representing diverse healthcare professionals, thus obtaining high face validity across stakeholders.

Manual data abstraction is the normative gold standard for national registries to ensure high accuracy, with data typically collected by trained abstractors who adhere to standardized data specifications. Some registries, such as the Society of Thoracic Surgeons (STS) Adult Cardiac Surgery Database (ACSD) [6], conduct annual external audits to validate data accuracy. The STS ACSD also has extremely high national penetration, capturing data from 95% of US non–federal cardiac surgery programs and 97% of all coronary artery bypass grafting (CABG) cases billed to Medicare [7].

Although preferred and trusted by many stakeholders, successful, widely utilized clinical registries like the STS ACSD are the exception. Broader use of clinical registries is typically constrained by the human effort required for data abstraction, the potential for incomplete data entry if not carefully audited, inconsistencies in reporting across institutions, and substantial costs associated with manual abstraction [8, 9].

Advances in artificial intelligence (AI) [10] offer an opportunity to transform clinical registry curation by automating data extraction. Algorithm-driven approaches to populating clinical registry data fields would reduce burden and costs and potentially increase coding uniformity across centers [11], but only if accuracy is maintained or increased. They might also permit collecting additional granular data currently infeasible to gather manually. Recent proof-of-concept studies have demonstrated the feasibility of AI-assisted extraction for individual clinical domains, including cancer registries [12] and selected cardiac surgery variables [13], but comprehensive solutions remain a significant challenge.

The automated extraction of structured information from clinical narratives has progressed through several methodological generations, from foundational tools such as cTAKES [14], MetaMap [15], and MedCAT [16] to deep learning and, most recently, large language models (LLMs) [17, 18, 19]. Systematic reviews have documented this evolution across hundreds of studies [20, 21, 22]. Registry automation has advanced furthest in cancer surveillance, where DeepPhe-CR extracts key cancer characteristics with F1 scores of 0.79–1.00 [12], expert-curated evaluation benchmarks have been established for oncology [23], and industry-scale NLP/ML pipelines have been developed for oncology real-world data [24]. In surgical domains, NLP has shown promise for infection surveillance [25] and orthopedic registry data collection [26]. Despite this progress, most systems remain disease-specific or institution-specific, and comprehensive, multi-variable registry abstraction, particularly for surgical quality registries with hundreds of data elements, remains an unmet challenge.

Accurate automation of EHR data extraction could have implications beyond clinical registries, potentially streamlining clinical research, hospital operations, billing, and quality oversight [3, 4, 11]. However, published AI systems for EHR abstraction predominantly rely on proprietary models such as GPT-3.5 and GPT-4 [18, 27, 28, 29] and target narrow clinical domains, from prostatectomy pathology [30] and spinal surgery operative notes [31] to oncology medication records [32], cancer registry curation [33], and social determinants of health extraction [34], with highly tailored designs that extract limited variables from single note types [35, 36, 37], raising concerns about reproducibility, transparency, and generalizability. More broadly, fewer than 10% of clinical AI models undergo external validation [38, 39, 40], and cross-site deployment often requires site-specific adaptation due to institutional variation in documentation practices [41, 42, 43]. Critically, few approaches have been tested against gold standard registries or validated across independent health systems, leaving unanswered whether AI can meet the stringent accuracy requirements of clinical and regulatory applications. Although commercial clinical data abstraction services exist for registries such as the STS ACSD (e.g., Q-Centrix [44] and Carta Healthcare [45]), these platforms combine human reviewers with proprietary software and may lack independent, published, peer-reviewed validation of their automated components.

Building on past small–scale efforts [19], this study develops and externally validates across Mass General Brigham (MGB) and Hartford HealthCare (HHC) a comprehensive framework for automated population of the STS ACSD. The proposed approach combines large language models with traditional NLP feature extraction to derive structured clinical variables from multimodal sources (clinical notes and structured tabular data) while ensuring high fidelity against manually abstracted registry data through confidence-based quality control mechanisms. Figure 1 provides an illustration of our pipeline. Tested across two major healthcare systems, the pipeline demonstrates that AI-assisted abstraction can achieve performance comparable to or exceeding human data collection while substantially reducing manual burden. Beyond registries, the same methodology can potentially generalize to any healthcare application that requires converting unstructured clinical documentation into standardized, structured data.

**Fig. 1:**
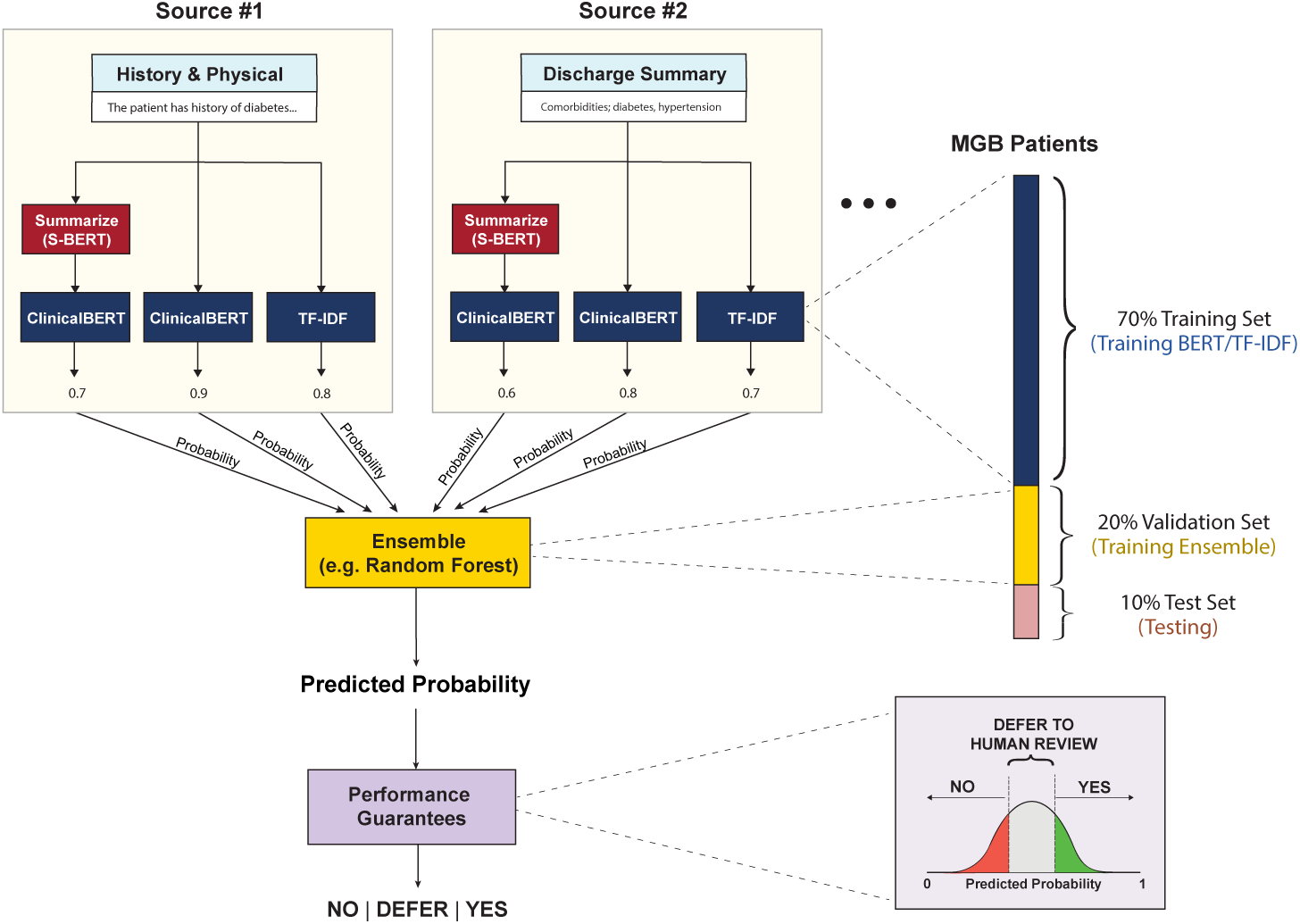
AI-driven registry curation pipeline overview for the target variable “history of diabetes“.

## 2 Results

We trained and evaluated the proposed AI-driven abstraction pipeline across two independent health systems to determine its accuracy, scalability, and potential to improve upon manual data collection.

### Patient Population

The study cohort consisted of 8,515 patients from Mass General Brigham (MGB) and 3,116 patients from Hartford Healthcare (HHC). The distribution of surgical procedures varied between sites, with CABG being the most common surgery at both institutions, accounting for 45% of cases at MGB and 50% at HHC. Aortic valve surgeries comprised 34% and 33% of procedures, respectively, while mitral valve operations accounted for 23% of cases at MGB and 27% at HHC. Patient demographics revealed a mean age of 64.2 years at MGB and 66.3 years at HHC, with a predominance of male patients in both cohorts. The proportion of patients with diabetes was higher at HHC (32%) compared to MGB (27%). Elective procedures comprised 79% of cases at both sites. Table 1 summarizes the patient characteristics across the two study sites. Table S3 in the Supplementary Information provides a detailed overview of the individual target variables distribution statistics.

**Table 1:**
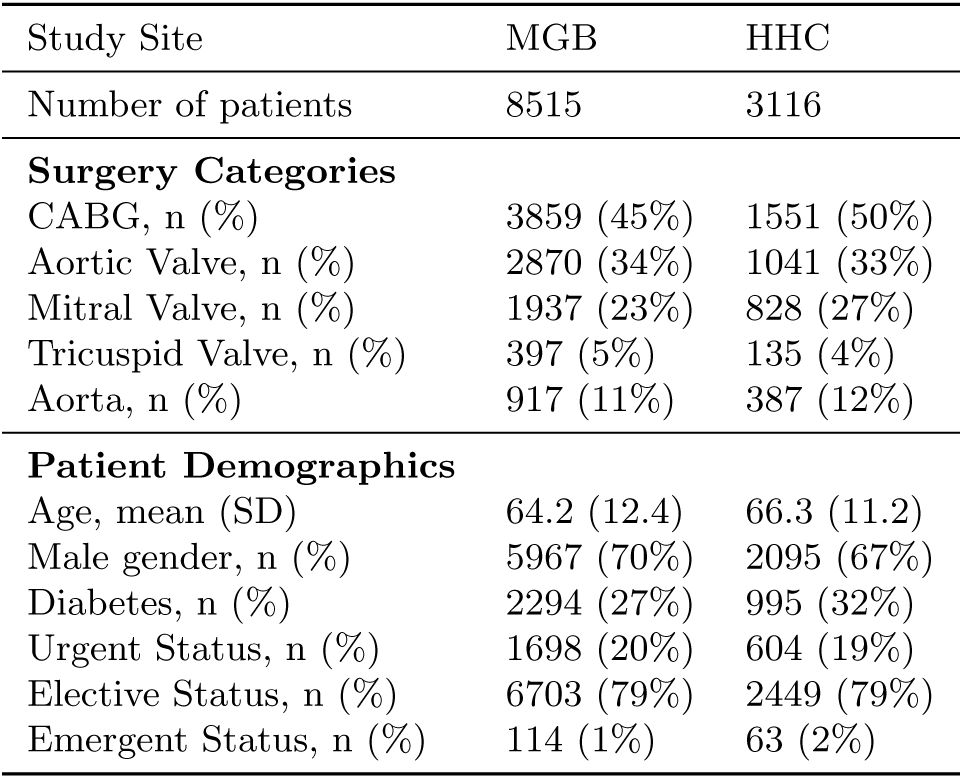
Patient characteristics and surgery types by study site.

The pipeline leveraged a diverse set of unstructured clinical documentation sources. These included history and physical reports, progress notes, visit notes, cardiology and anesthesia notes, operative notes, and discharge summaries. These records reflect the entirety of available patient documentation before and up to the index cardiac surgery encounter, ensuring that the pipeline captured all relevant preoperative and perioperative information. As shown in Table 2, the volume and length of documentation varied markedly between sites, highlighting the heterogeneity of EHR data across institutions. Cardiology notes were more frequent at MGB but substantially longer at HHC (1,369 vs. 241 words). MGB also had visit notes (mean 28 per patient), which were absent at HHC. The average word count of anesthesia notes was nearly double at MGB compared with HHC. Besides unstructured clinical reports, the pipeline also leveraged structured information sources, such as ICD-9/ICD-10 diagnoses codes, Lab Results and Medication information, as described in Section 4.

**Table 2:**
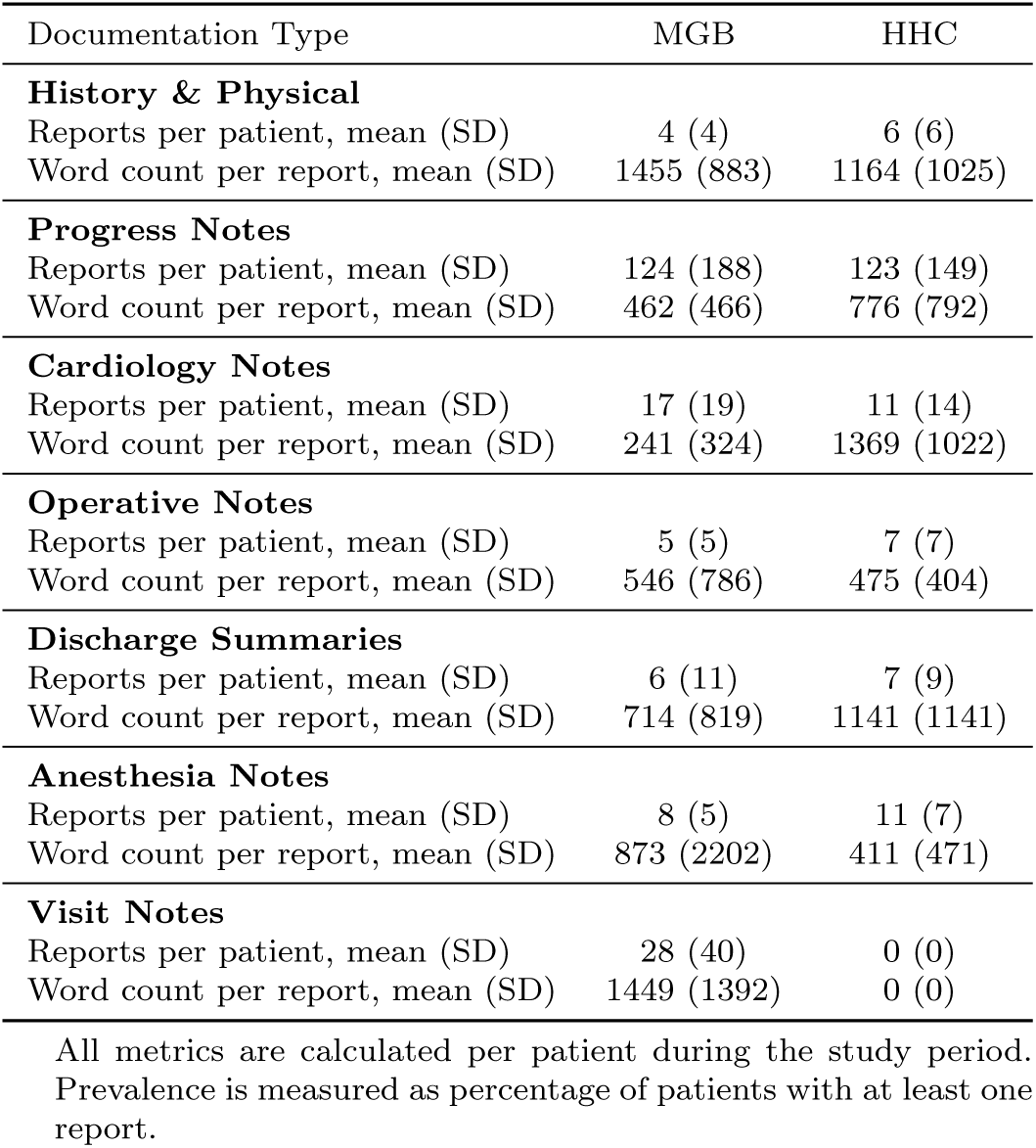
Summary statistics of unstructured sources of clinical documentation.

### Target Variables

Figure 2 provides a quantitative overview of the variable selection and categorization process. Specifically, starting from the total of 1,069 variables defined in the STS ACSD v4.20.2 specification sheet (All STS variables), 311 variables were initially excluded from the study because of insufficient data and extreme sparsity. In particular, these variables are populated in less than 0.15% of patients, making them unsuitable as targets for data abstraction (see Section 4.5 in the Supplementary Information for more details about excluded variables). From the remaining pool (758 Candidate variables), a further 111 variables did not meet the quality standards set for the automated extraction pipeline (Not completed by pipeline) and were therefore not populated by the final pipeline for any patient. This resulted in 647 variables successfully populated (i.e., Completed), at least partially, by our pipeline. These completed variables fall into the following categories: 40 variables are deterministic, extracted directly via rules from structured data; 173 variables are AI-derived, requiring direct inference by machine learning models; and 434 variables are parent/child variables, meaning their values were automatically populated because of an AI model assigning the value of its parent field.

**Fig. 2:**
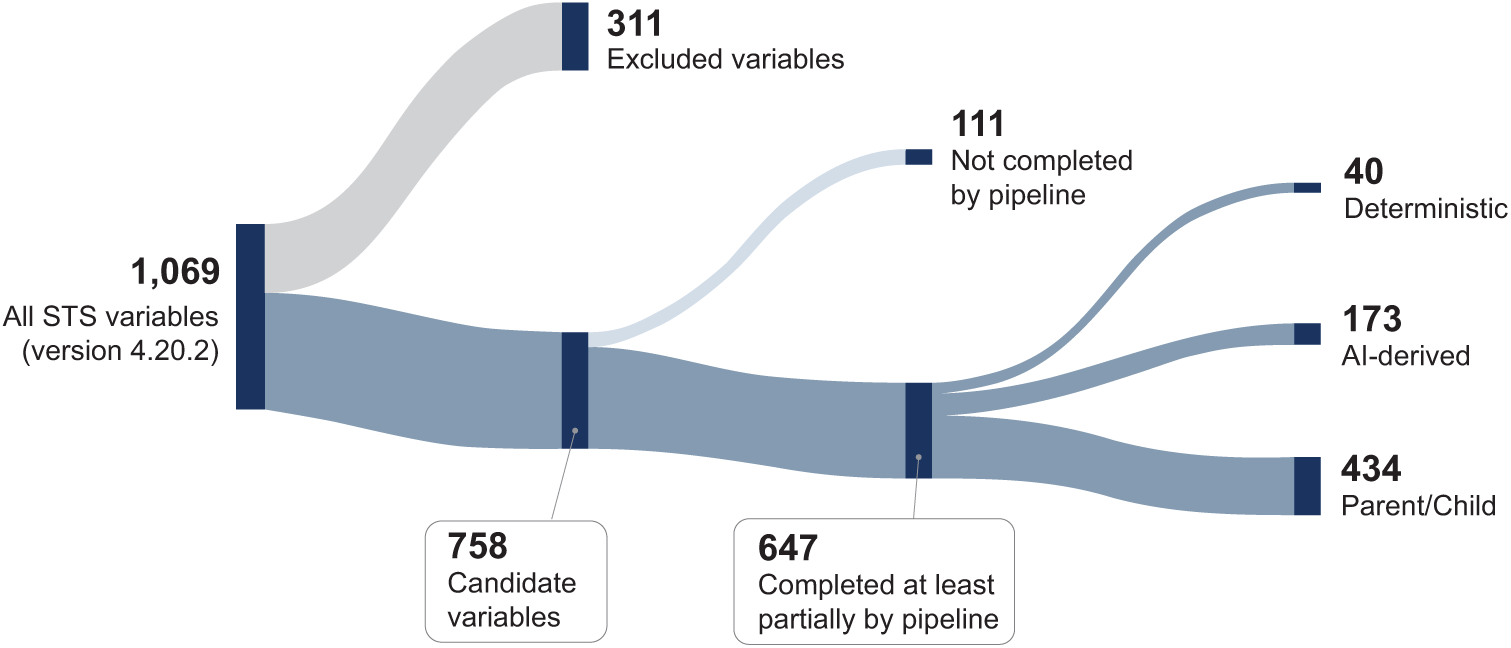
Breakdown of STS ACSD v4.20.2 variables. Starting from all 1,069 variables, 311 were excluded from the study as inapplicable because of insufficient data and high levels of sparsity. Out of the 758 remaining variables, 111 were not completed by the pipeline due to quality constraints. The remaining 647 variables were partially (or fully) populated either deterministically from structured data (Deterministic, 40), directly via AI models (AI-Derived, 173), or following the database hierarchy rules based on the AI-populated parent fields (Paren-t/Child, 434).

### Completion Accuracy and Data Completion Rate

Our automated pipeline was applied to 647 variables from the 758 variables of ACSD version 4.20.2 (see also Figure 2). To evaluate the overall performance of the pipeline, we first assessed the discriminative ability of the AI models across all AI-derived variables, after employing the dual-thresholding approach of Section 4. In the subset of records that the pipeline completes post-thresholding, we calculated AUC, precision, recall and specificity (Table S4 in the Supplementary Information). Although the precision is lower for some variables (see Section 4.2 in the Supplementary Information for a detailed discussion on precision and class imbalance), our pipeline achieves AUC, recall and specificity well above 90% for the majority of variables, indicating very strong discriminative performance.

After populating the AI-derived variables, we inferred the remaining non-deterministic variables through parent–child relationships. We evaluated completion rate and accuracy for both AI-derived and parent–child variables (Table S5 in the Supplementary Information). Accuracy was used because most parent–child variables are multiclass, making it the only quality metric applicable across all variables. We then incorporated the auto-populated deterministic variables and calculated overall performance and completion rate over the entire 758 candidate variables.

As shown in Figure 3 (see also Tables S7-S8 in the Supplementary Information), the output pipeline successfully maintained an overall accuracy that exceeds the threshold of 99% for the extracted outputs at both institutions, ensuring compliance with STS data quality standards. Regarding data collection burden reduction, the models achieved an automated cumulative completion rate of 49.5% across all variables at MGB and 43.2% at HHC, with higher performance in intraoperative data extraction (61.5% and 52.6%, respectively). The completion rate for preoperative variables was lower, at 35.0% for MGB and 31.4% for HHC. Among surgery types, CABG had the highest completion rates, reaching 53.5% at MGB and 46.3% at HHC, followed closely by mitral valve procedures.

**Fig. 3:**
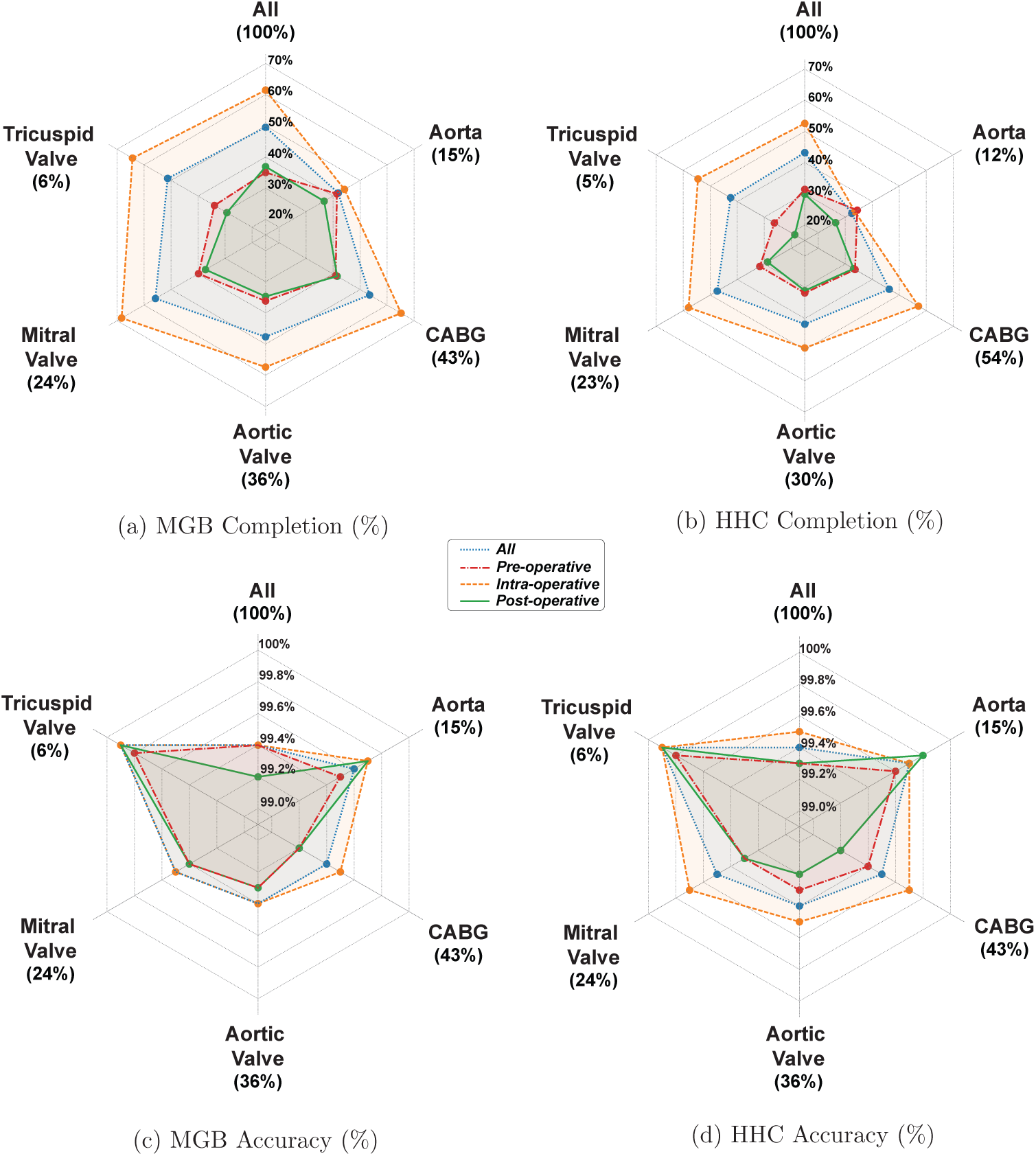
Pipeline performance summary for data completion rate and prediction accuracy across different study sites.

The individual performance metrics across non-deterministic target variables are presented in Table S5 in the Supplementary Information, while the deterministic target variables are summarized in Table S6 in the Supplementary Information. In Section 4.3 in the Supplementary Information, we further analyze the accuracy–completion trade-off, showing that relaxing the accuracy requirement from 99%+ to 94% would increase the completion rate in MGB from 49.5% to approximately 75%. However, in this study we select the operating point based on accuracy, in order to satisfy the stringent accuracy requirements of the STS database.

### Quality Control and Agreement with Manual Review

The manual review for some of the AI-derived target variables of the STS database revealed that some flagged disagreements between the ground truth and model predictions resulted from inconsistencies or errors in human abstraction rather than algorithmic inaccuracies. Specifically, for diabetes, 3.3% of cases required manual review by content-experts due to discrepancies between AI predictions and manually abstracted STS records. Upon blinded expert review of these cases, it was determined that 17.9% of the instances flagged as AI misclassifications were, in fact, correct predictions, indicating that the original label in the manually abstracted STS Database record was likely incorrect. Similarly, for CABG procedure identification, 1.2% of cases required manual review because of discordant results between manual abstraction and the AI algorithm. Among these cases, the AI-driven pipeline predictions were correct in 9.1% of identified cases, and the original, manually abstracted label from the STS Database record was likely inaccurate. Detailed results are presented in Table S9 in the Supplementary Information. Similarly, manual review of deterministic variables identified errors in coding, which can be categorized as: (i) minor data entry mistakes, such as typographical errors in addresses or small discrepancies in laboratory values; (ii) temporally incorrect entries, for example recording an intraoperative laboratory value instead of the pre-operative value for pre-operative A1C; and (iii) missing data, where the manually coded values were entirely absent from the EHR.

## 3 Discussion

Manual abstraction for a single large registry typically requires one to three full-time data managers, along with associated costs for hardware, software, and administrative oversight. Further, there are often substantial delays between patient discharge and data entry completion. This study demonstrates that AI-driven multimodal abstraction can overcome these barriers, delivering registry-grade accuracy, enhancing the efficiency and scalability of clinical registry population. Our pipeline successfully automates 647 of 758 candidate variables of the STS ACSD, achieving an overall accuracy exceeding 99% at both study sites while automating 49.5% of the total data abstraction burden at MGB and 43.2% at HHC. This demonstrates very high accuracy while covering a substantial portion of the abstraction workload.

A dual-threshold confidence control framework enables AI predictions to meet predefined accuracy thresholds, preventing low-confidence outputs from entering downstream applications. Furthermore, the AI system can detect potential human errors, allowing it to function as a quality improvement tool by flagging discrepancies for human review and correction. For example, blinded expert review of cases where AI predictions disagreed with manually abstracted STS records showed that 17.9% of flagged diabetes cases and 9.1% of flagged CABG cases reflected errors in the original human abstraction rather than AI misclassifications (Table S9 in the Supplementary Information).

The pipeline is designed to be agnostic to EHR vendors, data modalities, and the specific class of language models deployed, making it adaptable across healthcare systems with varying infrastructures. This adaptability was validated across two health systems with substantially different EHR configurations and documentation practices: for instance, cardiology notes averaged 241 words per report at MGB versus 1,369 at HHC, visit notes were available only at MGB, and anesthesia note lengths differed nearly twofold between sites (Table 2). Despite this heterogeneity, the pipeline maintained accuracy above 99% at both institutions.

To achieve this robustness across diverse clinical environments, we combined feature extractors such as ClinicalBERT and TF-IDF, balancing predictive performance with computational cost and real-world deployment constraints. These constraints are primarily dictated by strict HIPAA regulations and Protected Health Information (PHI) protections, which lead health system IT governance to prohibit the transmission of raw clinical notes to proprietary cloud-based LLM APIs or third-party cloud providers. While the on-premise deployment of open-source frontier models (e.g., Llama-3 70B) might appear to be a viable alternative, the resulting computational burden—requiring massive, multi-GPU clusters—is often financially and operationally out of reach for most hospital systems.

In contrast, smaller, targeted models like ClinicalBERT and TF-IDF offer highly efficient, low-latency inference that runs more easily on standard institutional hardware. Moreover, recent work has shown that even when frozen state-of-the-art generative LLMs are technically feasible for prediction, they often exhibit similar or worse performance than traditional machine-learning models in clinical settings unless fine-tuned or incorporated into more complex pipelines (e.g. RAG) [46, 47, 48]. Nevertheless, our modular ensemble architecture, in which each variable is served by an independent pipeline of 30 base models, allows individual featurization or classification components to be updated or replaced without retraining the entire system, thereby providing a natural pathway for incorporating future advances in clinical language modeling.

Such frameworks can easily be extrapolated to other healthcare uses in which the data source is the EHR and the end product is a data application requiring standardized, accurate data. AI-driven data abstraction not only reduces inter-institutional variability but can potentially also increase the consistency and fidelity of data collection. By introducing standardized, data-driven extraction, such pipelines can minimize between-institution deviations in the coding of structured variables and enhance the granularity and consistency of recorded information across participating sites. Automated extraction methods enable the extraction of a comprehensive range of clinical features. This increased data richness allows for more precise risk stratification, quality benchmarking, and research applications.

Despite its advantages, the AI-driven approach also presents certain limitations. First, while AI models perform well in structured environments, their accuracy may decline when handling ambiguous or inconsistently documented data. Moreover, external validation in diverse healthcare systems remains crucial to ensuring model generalizability. The exclusion of transferred patients in this study, due to challenges in reconciling multi-institutional data, highlights the need for future research on interoperability solutions. Additionally, the reliance on historical, manually abstracted data as training labels means that the model inherits existing biases and inaccuracies from human abstraction processes. Furthermore, the confidence thresholds optimized on the derivation cohort may require recalibration when the pipeline is deployed at new sites or when underlying documentation practices evolve, as dataset shift can alter the relationship between model confidence and prediction accuracy.

Class imbalance also affects pipeline performance. Variables representing rare clinical conditions tend to achieve lower automated completion rates under stringent confidence thresholds. This severe class imbalance is a primary driver of the broader accuracy-completion tradeoff inherent to our pipeline: Enforcing near-perfect accuracy dictates a higher rate of manual deferral, especially for rare events. Consequently, our > 99% accuracy requirement limits automated completion to roughly half the abstraction workload. However, the system remains highly tuneable. Relaxing this threshold to a 94% accuracy target could elevate the completion rate to approximately 75%, highlighting the pipeline’s adaptability for applications with different risk tolerances. (see Section 4.3 and Figure S5 in the Supplementary Information)

Future improvements should focus on refining models with independent expert-labeled datasets, expanding generalizability across sites with different EHR configurations, and optimizing hybrid frameworks that combine AI-driven automation with targeted human oversight. Also future iterations may incorporate next-generation open-weight models capable of integrating text, imaging, and tabular data.

Given these limitations, a hybrid system that integrates AI-driven automation with selective human oversight of certain data elements offers a practical and effective solution. AI can automate the bulk of data abstraction while flagging uncertain or ambiguous cases for human review. This approach ensures high efficiency while maintaining quality standards, allowing human abstractors to focus on complex cases where AI models may lack contextual understanding. Such human-in-the-loop strategies are likely to accelerate trust and adoption in clinical environments.

## 4 Methods

### Study Design

We conducted a retrospective, proof-of-concept study with external validation to assess the feasibility and accuracy of an AI-assisted approach for automated data extraction from the EHR into the STS ACSD. The study took place across two major healthcare systems: Mass General Brigham (MGB), serving as the derivation and internal validation site of the data-driven models; and Hartford HealthCare (HHC), serving as the external validation site, to ensure consistency and reliability. The study developed an AI-driven data curation pipeline and compared it with manual abstraction to assess efficiency, accuracy, and completeness. The experiments on MGB and HHC were performed using on-premise computational resources.

The study included adult patients undergoing cardiac surgery at the participating institutions. The MGB cohort included all adult cardiac surgery patients recorded in the STS ACSD between 1 January 2015 and 30 September 2021. For the external validation cohort (HHC), we used STS records of all cardiac surgery patients in the HHC cardiac surgery program recorded between 1 July 2020 and 31 December 2023. Patients were excluded from both the MGB and HHC cohorts if they were transferred from another acute or non-acute care facility, or if their STS records were missing, or if key timestamps such as admission, operating room entry and exit, or discharge were absent or inconsistent.

The study protocol was separately reviewed and approved by the Institutional Review Boards of Mass General Brigham (protocol ID: 2020P004072) and Hartford HealthCare (protocol ID: M-HHC-2023-0209). The requirement for informed consent was waived by the institutional review boards of MGB and HHC due to the retrospective nature of the study.

### Target Variables

The data extraction targets correspond to the systematically recorded variables within the STS ACSD database. To determine their nominally “correct,” gold standard values, we used the records retrospectively submitted to the STS ACSD by the collaborating healthcare systems. These values have been abstracted by trained, expert human nurses, typically employed by each hospital institution, who strictly adhere to standardized data specifications [9]. Furthermore, to ensure high data accuracy and inter-annotator reliability across institutions, the STS conducts rigorous annual external audits of these submitted records [9].

In our analysis, we categorize the variables recorded in the STS ACSD (version 4.20.2) into two distinct types: *deterministic* and *non-deterministic*. *Deterministic target variables* (e.g., demographic information or discrete lab results) are directly extracted from the EHR, without resorting to the use of AI. These variables are well-structured, straightforward to locate within the database, and do not require complex inference. They are abstracted using local, rule-based extraction methods tailored to the data format of each institution. For variables recorded in units that differ from those specified by the STS standard (e.g., laboratory values, height, and weight), institution-specific pipelines also perform unit harmonization to ensure compliance with the STS specification.

*Non-determinstic target variables* (e.g. comorbidities such as diabetes or hypertension) exhibit substantial clinical and documentation variability, rendering rule-based extraction unreliable. A subset of these variables (AI-derived variables) are directly estimated using ML models that are trained on multimodal EHR data. These variables are either binary in nature (e.g. history of cancer) or categorical variables that can be meaningfully binarized (e.g. chronic lung disease classified as none/mild/severe). A second subset of the Non-deterministic variables are the Parent/Child variables. These are populated using inferred values of the AI-derived variables, by enforcing hierarchical rules of the STS ACSD database (e.g., child fields are set to “no” when the parent variable is coded as “no”).The breakdown of the STS ACSD variables included in our analysis can be found in Figure 2.

### Data Sources

Structured and unstructured EHR data were used as the AI pipeline input. *Unstructured* sources refer to clinical free text reports including: anesthesia notes; cardiology notes; history and physical examination reports; operative notes; visit notes; progress notes; and discharge summaries.. *Structured* data consist of laboratory results; medication administration records; and diagnosis codes. Detailed definitions of each note category are presented in Section 2.2 in the Supplementary Information. Statistics and prevalence of each note type are shown in Table 2.

### Data Homogenization Across Study Sites

To ensure consistency and comparability across heterogeneous EHR systems, a standardized data homogenization process was implemented across both structured and unstructured sources. For unstructured clinical reports, site-specific pipelines (such as parseRPDR [49]) were applied to remove artifacts, normalize formatting, and harmonize documentation retrieved from the MGB data registry and HHC EPIC databases. To address cross-hospital discrepancies in structured data, structured fields were also converted to text using TabText [50], while also retaining the original structured format for pipeline integration. International Classification of Diseases (ICD-9/ICD-10) codes were mapped to clinically interpretable categories, represented both as structured variables and text descriptors. Laboratory values were preserved in numeric form for structured analyses and additionally converted into natural language representations, and patient medication lists were similarly transformed into text. Together, these procedures ensured that all sources, regardless of format or origin, were processed in a uniform manner suitable for predictive modeling and analysis.

### AI-Driven Pipeline

Our automated data extraction system consists of independent predictive modeling pipelines, one per target variable. Each pipeline was trained independently, while using identical patient-level training, validation, and testing splits across variables. For each target variable, unstructured clinical text was represented using three distinct featurization methods, improving robustness to heterogeneous data formats and variability in clinical note structure. Predictions from each data source and feature representation were then aggregated by a second-stage ensemble model to generate the final variable estimate (Figure 1).

As our first featurization method we used ClinicalBERT, a transformer model pre-trained on over two million clinical notes from MIMIC-III [51, 52]. For each target variable, Clinical-BERT was fine-tuned separately for each text source (e.g., discharge summaries, carrdiology notes) using transfer learning, with a classification head applied to the [CLS] token. The resulting source-level prediction probabilities were used as input features for the downstream ensemble model.

Because perioperative clinical notes are often lengthy and exceed ClinicalBERT’s token limits when concatenated, we implemented a complementary summarization-based featurization approach. Sentence-BERT (S-BERT) [53] was used to compute semantic similarity between individual sentences and the target variable, guided by the STS data abstraction manual. Sentences were ranked by similarity, and the most relevant subset was selected to fit model capacity constraints. ClinicalBERT was subsequently fine-tuned on these summarized documents to produce an alternative estimate of the target variable. Note that all models were trained using on-premise GPU resources, which precluded the use of much larger frontier models.

To complement transformer-based models, we incorporated TF-IDF vectorization [54]. Despite its simplicity, TF-IDF is computationally efficient, interpretable, and competitive with transformer models in clinical NLP tasks [55, 56]. TF-IDF embeddings were fed into standard machine-learning classifiers, and the resulting prediction probabilities were provided as inputs to the ensemble model.

In the final stage, predictions from all source-level models were aggregated using a supervised ensemble learning framework. A meta-learner was trained on a held-out validation set using probability outputs from 30 base models (10 data sources × 3 featurization methods). This approach adaptively weighted sources for each target variable based on data quality and task relevance (see Figure S2 in the Supplementary Information), yielding robust predictions even when individual sources were noisy or incomplete.

All models were developed using the MGB derivation cohort with patient-stratified splits: 70% of patients for training source-specific and language models, 20% for validation and ensemble training, and 10% for final testing. All reported performance metrics are based on the held-out test set. Stratification was performed at the patient level to ensure that all records from the same patient remained in a single partition, preventing data leakage and preserving independence across training, validation, and testing sets, which was essential given multiple visits per patient in the STS database.

Pre-trained language models were fine-tuned and downstream predictors trained using standard hyperparameter optimization procedures [57]. ClinicalBERT models were fine-tuned for only three epochs to avoid catastrophic forgetting [58], with learning rate, batch size, and weight decay optimized on the validation set. For TF-IDF–based predictors and the ensemble model, multiple model types were evaluated and the best-performing configuration was selected per variable using the validation set. All pipeline components—including feature extraction, structured data integration, and ensemble learning—were trained independently for each target variable.

### Quality Assurance via Double Thresholding

To calibrate the pipeline outputs against the stringent accuracy requirements of the STS ACSD, we incorporated a two-tiered quality control framework. At the prediction level, we implemented a dual-threshold confidence system, inspired by [59, 60], to classify outputs into three zones illustrated in Figure 4: predictions below a lower bound (*t*_1_) were labeled negative, those above an upper bound (*t*_2_) were labeled positive, and values falling between the two thresholds were flagged for manual review. Thresholds were optimized on the validation set such that both sensitivity and specificity exceed the 96% level, with the objective of maximizing automated completion rates while meeting the accuracy standards mandated by STS.

**Fig. 4:**
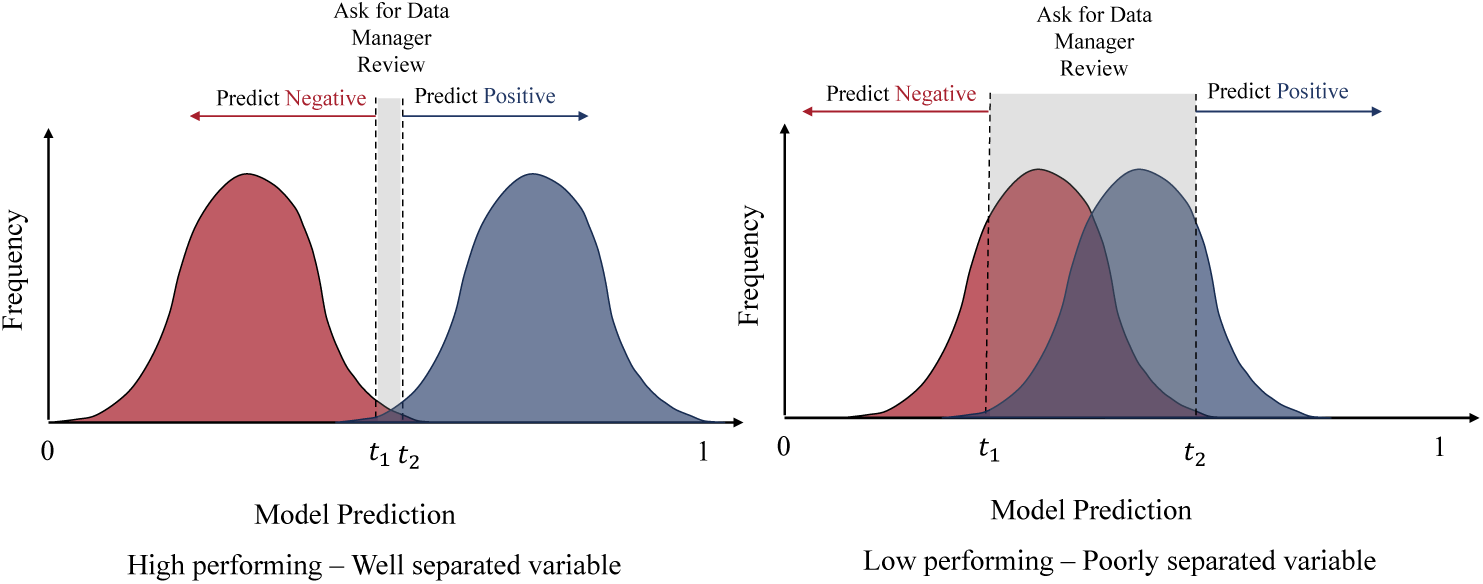
Illustration of dual-threshold approach for prediction quality assurance. Predictions below the lower threshold (*t*_1_) are classified as negative, while those above the upper threshold (*t*_2_) are classified as positive. Predictions between the thresholds are flagged for manual review. Better models will have a lower proportion of predictions requiring manual review.

Each non-deterministic target variable was further required to achieve ≥96% accuracy on automated predictions within the testing set to remain in the automated pipeline. Variables failing to meet this benchmark reverted entirely to manual review, regardless of prediction-level confidence. This dual approach ensured that high-confidence predictions were automated while ambiguous or lower-performing variables were subject to manual oversight, thereby balancing efficiency with accountability. Empirically, these safeguards provided consistent, high-quality outputs and reduced the risk of error propagation.

### Model Evaluation

For comprehensive evaluation of the pipeline, a critical component is the hierarchical nature of the STS database. For instance, when evaluating post-operative complications, the presence of a parent variable (e.g., “any post-operative complication”) governs the logical assessment of child complications (e.g., “stroke” or “infection”). Our system enforces this via deterministic rules: if a parent is negative, all child variables are set to negative; if a parent is positive or flagged for review, child variables retain their independent predictions. To provide a holistic view of system performance, we harmonize model predictions across these hierarchical relationships, ensuring logical consistency while preserving the granularity of the underlying data.

The pipeline is evaluated in two stages. First, we compute metrics such as AUC, precision, recall, and specificity on AI-derived variables to provide detailed performance insights into the individual ensemble models. In the second stage, we assess overall pipeline performance using two measures: (1) prediction accuracy, defined as the proportion of correctly inferred variables compared to manually abstracted registry values, and (2) data completion rate, which represents the percentage of target variables automatically populated using the pipeline at a minimum accuracy threshold. These metrics inherently trade off against each other; higher accuracy thresholds may reduce the automated completion rate, while lower thresholds may risk compromising reliability. The threshold levels of the pipeline were selected by investigating this tradeoff, choosing an operating point that meets the rigorous 99% accuracy standard mandated by STS (Figure S5 in the Supplementary Information).

### Quality Control and Agreement with Manual Review

Discrepancies between AI and manual coding may reflect errors in human abstraction rather than AI inaccuracies. To assess this hypothesis, we conducted a comprehensive quality control experiment to evaluate both deterministic and AI-derived target variables. For deterministic laboratory variables (including the last preoperative A1C, creatinine, and hematocrit values) and demographic variables (such as patient address), we manually reviewed cases in which the automated extraction disagreed with the STS record. For AI-derived variables, a validation protocol was conducted at the testing set of the MGB cohort to assess accuracy and potential biases for the targets of “history of diabetes” and “coronary artery bypass grafting (CABG) procedure.” A blinded manual review was performed on the cases where AI predictions disagreed with STS records that had been manually populated. An expert clinician reassessed flagged cases without knowledge of the AI output or original registry values, documenting justifications for any changes.

## Declarations

### Data availability

The data used in our study come from two academic medical centers in the United States subject to the Health Insurance Portability and Accountability Act. Due to the data use agreement that we have signed with Hartford HealthCare and Massachusetts General Brigham, the datasets cannot be made publicly accessible, as they contain protected health information and other sensitive information about the patients. Any user that wishes to gain access to the dataset needs to complete HIPAA training, obtain Institutional Review Board approval, and sign a data use agreement with the collaborating institutions.

### Code Availability

The code used to generate the results of this study is not publicly available because it is subject to contractual agreements and intellectual property constraints.

## Acknowledgements

We acknowledge Dr Thor Sundt (Mass General Brigham) and Dr Barry Stein (Hartford HealthCare) for facilitating access to data. We also acknowledge Carole Krohn, Director of the STS National Databases, and Shari Sarnevitz (Mass General Brigham) for guidance on data recording and extraction practices. Georgios Margaritis and Periklis Petridis were supported for this work by a research grant from STS. The grant was directly provided from STS to MIT and thus does not involve a specific grant number. The remaining authors did not receive any financial support for their involvement in this study.

## Author contributions

GM: data curation, formal analysis, investigation, methodology, software, validation, visualization, writing - review & editing. PP: conceptualization, data curation, formal analysis, investigation, methodology, project administration, software, validation, visualization, writing - original draft, writing - review & editing. DB: funding acquisition, supervision, writing - review & editing. JB: resources, writing - review & editing. RHag: resources, writing - review & editing. RHab: funding acquisition, supervision, writing - review & editing. DMS: conceptualization, funding acquisition, investigation, supervision, validation, writing - review & editing. AO: conceptualization, funding acquisition, methodology, project administration, supervision, writing - original draft, writing - review & editing.

## Competing interests

The authors declare no competing financial or non-financial interests.

## Notes

### Competing Interest Statement

The authors have declared no competing interest.

### Author Declarations

- IRB of Mass General Brigham approved the study protocol (protocol ID: 2020P004072) - IRB of Hartford HealthCare approved the study protocol (protocol ID: M-HHC-2023-0209)

