## Supplementary Information for "Artificial Intelligence for Automated, Highly Accurate, and Scalable Multimodal EHR Data Abstraction"

The Supplementary Appendix represents an expanded version of the Results of the research described in the main body of the article.

The Appendix is organized in the following sections:

1. **Glossary** provides definitions for the key technical terms used throughout the document.
2. **Data Description** presents an overview of the study cohorts, analyzes the available data sources and presents data distribution and class balance of the target variables.
3. **Pipeline Performance Results** evaluates the accuracy, completion rates, and predictive performance of the AI-driven extraction pipeline.
4. **Additional Result Analysis** provides further analysis of the results and the pipeline performance, focusing on aspects such as feature importance, accuracy-completion tradeoff and error analysis.

#### 1 Glossary

This section provides definitions for key terms and concepts that are frequently referenced throughout the document. It is intended to ensure clarity and consistency in understanding the methodologies, techniques, and terminologies used in the study.

**ClinicalBERT** A transformer-based natural language processing model adapted from BERT (Bidirectional Encoder Representations from Transformers) and fine-tuned on large-scale clinical data. ClinicalBERT is specifically designed to process and analyze unstructured clinical narratives, such as progress notes and discharge summaries, capturing domain-specific semantics for healthcare applications.

**CABG (Coronary Artery Bypass Grafting)** A surgical procedure used to treat coronary artery disease by creating a bypass around blocked or narrowed coronary arteries using blood vessels from other parts of the body, thereby improving blood flow to the heart muscle.

**Confidence Threshold** A predefined value used to determine whether a machine learning prediction is considered reliable. Predictions below the lower threshold are classified as negative, those above the upper threshold as positive, and intermediate cases are flagged for manual review.

**Cross-validation** A robust statistical technique for evaluating machine learning model performance by dividing data into multiple subsets or folds. Models are trained on a combination of subsets and validated on the remaining unseen data, ensuring comprehensive performance assessment and aiding in the optimization of hyperparameters while minimizing the risk of overfitting.

**Data Harmonization** The process of standardizing data from different institutions to ensure consistency, including diagnostic coding and formatting of unstructured clinical text.

**Embedding** A numerical vector representation of data (text, images, etc.) that captures semantic meaning in a format suitable for machine learning models.

**Ensemble Model** A machine learning approach that combines predictions from multiple individual models (in our case, predictions from different data sources and feature extraction methods) to produce more robust and accurate final predictions.

**External Validation** Assessment of model performance using data from a different institution (HHC in our case) to evaluate generalizability.

**Feature** An individual measurable property or characteristic used as input for machine learning models. Features can be derived from both structured data (e.g., lab values) and unstructured data (e.g., clinical notes).

**Feature Extraction** The process of systematically transforming raw structured and unstructured data into a set of measurable and meaningful variables (features) that serve as inputs for machine learning models.

**Generalizability** The ability of a model to maintain performance on external data, such as the HHC cohort, ensuring its applicability across diverse patient populations and institutions.

**HHC Cohort** The external validation dataset from Hartford Healthcare, consisting of 4,173 cardiac surgery patients from August 2020 through December 2023, used solely for evaluating model generalizability.

**Hierarchical Data** Data organized in a structured parent-child relationship, where the values of parent variables define or constrain the possible values or interpretations of child variables.

**Hyperparameter** Configuration variables that control the learning process, such as learning rate, batch size, or model architecture choices, which are set before training begins.

**Internal Validation** Assessment of model performance using held-out data (testing set) from the same institution where the model was developed (MGB in our case).

**MGB Cohort** The derivation dataset from Massachusetts General Brigham hospital system used for model development, comprising 6,539 cardiac surgery patients from January 2015 through September 2021.

**Multimodal Data** Data that combines structured and unstructured information from clinical records, including lab values, medication histories, and free-text clinical narratives, to capture a comprehensive view of patient characteristics.

**Natural Language Processing (NLP)** A subfield of artificial intelligence and machine learning focused on the interaction between computers and human language, enabling the processing, analysis, and generation of unstructured text data for various tasks such as classification, summarization, and semantic understanding.

**Overfitting** A common machine learning issue in which a model performs exceptionally well on training data but fails to generalize to new, unseen data due to excessive complexity or reliance on noise and irrelevant patterns in the training set.

**S-BERT (Sentence-BERT)** A variant of BERT designed for sentence-level tasks, such as semantic similarity and text summarization. S-BERT uses a Siamese network architecture to generate high-quality embeddings for individual sentences, enabling efficient comparison and summarization of clinical texts in this study.

**Structured Data** Data organized in a predefined format, such as database tables containing laboratory values, medications, diagnostic codes, or demographic information.

**Structured Variable** Data stored in a predefined format, such as laboratory values, medications, or diagnostic codes, typically organized in database tables.

**Target Variable** The outcome variable that our models aim to predict, corresponding to specific fields in the STS database.

**Testing Set** The portion of data (10% of MGB cohort) reserved for internal validation.

**TF-IDF (Term Frequency-Inverse Document Frequency)** A natural language processing technique used to represent textual data by calculating the importance of words within a document relative to a corpus.

**Training Set** The portion of data (70% of MGB cohort) used to train the initial models.

**Transformer Model** A state-of-the-art deep learning architecture based on self-attention mechanisms, designed to model sequential data dependencies effectively. Transformer models, such as BERT and ClinicalBERT, have become foundational for tasks involving natural language processing.

**Unstructured Data** Data without a predefined structure, primarily consisting of free-text clinical narratives such as progress notes, discharge summaries, and operative reports.

**Validation Set** The portion of data (20% of MGB cohort) used to tune the ensemble model parameters and prevent overfitting.

### 2 Data Description

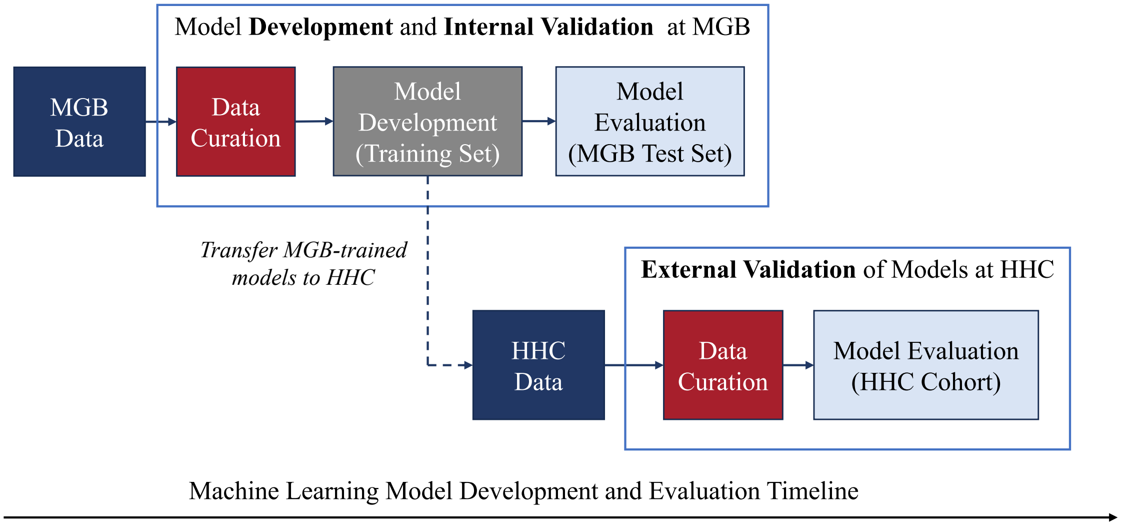


Figure S1: Model development and validation pipeline overview.

#### 2.1 Patient Population and Hospital Characteristics

Table S1: Patient characteristics and surgery types stratified by study site

| Characteristic | MGB | HHC |
|---|---|---|
| Number of patients | 8515 | 3116 |
| **Surgery Categories** | | |
| CABG, n (%) | 3859 (45%) | 1551 (50%) |
| Aortic Valve, n (%) | 2870 (34%) | 1041 (33%) |
| Mitral Valve, n (%) | 1937 (23%) | 828 (27%) |
| Tricuspid Valve, n (%) | 397 (5%) | 135 (4%) |
| Aorta, n (%) | 917 (11%) | 387 (12%) |
| **Patient Demographics** | | |
| Age, mean (SD) | 64.2 (12.4) | 66.3 (11.2) |
| Male gender, n (%) | 5967 (70%) | 2095 (67%) |
| Diabetes, n (%) | 2294 (27%) | 995 (32%) |
| Urgent Status, n (%) | 1698 (20%) | 604 (19%) |
| Elective Status, n (%) | 6703 (79%) | 2449 (79%) |
| Emergent Status, n (%) | 114 (1%) | 63 (2%) |

#### 2.2 Clinical Documentation Characteristics

Below we present the types of notes that each unstructured of source encompasses, in accordance with the Research Patient Data Registry (RPDR) specificiation, which is the centralized clinical data warehouse aggregating structured and unstructured electronic health record data from Partners HealthCare institutions:

- **History & Physical:** Often created as part of a hospital admission. Typical components include the Chief complaint, the main reason the patient is seeking medical care; History of Present Illness (HPI), describing the evolution of the current illness, including any events occurring in the Emergency Department; Past medical history; Medication list; Social history; Family history; Vital signs and Physical exam; overview of Laboratory, Radiology, and Microbiology results; Assessment; and Plan.

- **Progress Note:** Progress reports can be classified by different types, such as Perioperative; Labor & Delivery; Emergency, Trauma, or Observations; Nursing, etc. These are typically inpatient notes created during a patient's hospitalization.
- **Cardiology Reports:** Diagnosis, new findings and interpretation; routine and procedure-specific measurements; tests results such as echocardiogram, EKG, cardiac catheterization, electrophysiology, exercise/stress test, Holter and event monitoring, nuclear perfusion imaging, vascular studies.
- **Operative Note:** Diagnosis, Indications for surgery, Procedure performed; specimens; drains; blood loss; condition at the end of the procedure; disposition
- **Discharge Summary:** Admission and discharge dates; Principal and secondary diagnoses; Summary of patient hospitalization, which may reproduce some information from the Admission HPI; Test results; Procedures performed; Patient status at discharge; and discharge disposition.
- **Anesthesia Notes:** Pre-anesthetic evaluation (American Society of Anesthesiologists [ASA] physical status classification, airway assessment); anesthesia plan; intraoperative record (continuous vital signs, fluid intake/output, medications administered, anesthetic agents used); airway management details; type of anesthesia (e.g., general, regional, MAC); and Post-Anestheisa Care Unit [PACU] handoff/emergence status.
- **Visit Note:** Reason for visit; diagnosis; patient symptoms, chief complaint; vital signs and physical exam; labs and imaging results; medications; assessment; plan

Table S2: Summary Statistics of Clinical Documentation Sources

| Documentation Type | MGB | HHC |
|---|---|---|
| **History & Physical** | | |
| Reports per patient, mean (SD) | 4 (4) | 6 (6) |
| Word count per report, mean (SD) | 1455 (883) | 1164 (1025) |
| **Progress Notes** | | |
| Reports per patient, mean (SD) | 124 (188) | 123 (149) |
| Word count per report, mean (SD) | 462 (466) | 776 (792) |
| **Cardiology Notes** | | |
| Reports per patient, mean (SD) | 17 (19) | 11 (14) |
| Word count per report, mean (SD) | 241 (324) | 1369 (1022) |
| **Operative Notes** | | |
| Reports per patient, mean (SD) | 5 (5) | 7 (7) |
| Word count per report, mean (SD) | 546 (786) | 475 (404) |
| **Discharge Summaries** | | |
| Reports per patient, mean (SD) | 6 (11) | 7 (9) |
| Word count per report, mean (SD) | 714 (819) | 1141 (1141) |
| **Anesthesia Notes** | | |
| Reports per patient, mean (SD) | 8 (5) | 11 (7) |
| Word count per report, mean (SD) | 873 (2202) | 411 (471) |
| **Visit Notes** | | |
| Reports per patient, mean (SD) | 28 (40) | 0 (0) |
| Word count per report, mean (SD) | 1449 (1392) | 0 (0) |

All metrics are calculated per patient during the study period Prevalence is measured as percentage of patients with at least one report.

##### 2.3 Variable Distribution Statistics

Table S3: Class Distribution for AI-derived Variables

| Variable | Value | MGB | | HHC |
|---|---|---|---|---|
| | | Pat. Train | Pat. Test | Pat. Test |
| **Pre-operative** | | | | |
| RF-Family History of Premature CAD | Yes | 949 | 123 | 263 |
| | No | 6705 | 738 | 2853 |
| RF-Diabetes | Yes | 2061 | 233 | 995 |
| | No | 5593 | 628 | 2121 |
| RF-Renal Fail-Dialysis | Yes | 107 | 9 | 66 |
| | No | 7547 | 852 | 3050 |
| RF-Hypertension | Yes | 5823 | 651 | 2669 |
| | No | 1831 | 210 | 447 |
| RF- Endocarditis | Yes | 320 | 40 | 122 |
| | No | 7334 | 821 | 2994 |
| RF-Pulmonary Function Test | Yes | 746 | 73 | 436 |
| | No | 6908 | 788 | 2678 |
| RF-Home Oxygen | Yes | 103 | 10 | 52 |
| | No | 7551 | 851 | 3064 |
| RF-Inhaled Medication or Oral Bronchodilator Therapy | Yes | 866 | 90 | 468 |
| | No | 6788 | 771 | 2648 |
| RF-Sleep Apnea | Yes | 1291 | 144 | 691 |
| | No | 6363 | 717 | 2425 |
| RF-Pneumonia | Recent | 830 | 85 | 376 |
| | No | 6824 | 776 | 2740 |
| RF-Liver Disease | Yes | 288 | 30 | 116 |
| | No | 7366 | 831 | 3000 |
| RF-Liver Cirrhosis | Yes | 16 | 3 | 33 |
| | No | 7638 | 858 | 3083 |
| Immunocompromised Present | Yes | 388 | 41 | 199 |
| | No | 7266 | 820 | 2917 |
| Mediastinal Radiation | Yes | 221 | 16 | 77 |
| | No | 7433 | 845 | 3039 |
| Cancer Within 5 Years | Yes | 703 | 88 | 241 |
| | No | 6951 | 773 | 2875 |
| Peripheral Artery Disease | Yes | 730 | 87 | 428 |
| | No | 6924 | 774 | 2688 |
| Syncope | Yes | 350 | 37 | 138 |
| | No | 7304 | 824 | 2978 |
| Cerebrovascular Disease | Yes | 1205 | 123 | 525 |
| | No | 6449 | 738 | 2591 |
| Prior CVA | Yes | 597 | 58 | 295 |
| | No | 7056 | 803 | 2819 |
| CVD Prior Carotid Surgery | Yes | 175 | 15 | 64 |
| | No | 7478 | 846 | 3049 |
| Five Meter Walk Test Done | Yes | 21 | 4 | 86 |

Table S3: Class Distribution for AI-derived Variables

| Variable | Value | MGB | | HHC |
|---|---|---|---|---|
| | | Pat. Train | Pat. Test | Pat. Test |
| | No | 7633 | 857 | 3030 |
| Prev Cardiac Interventions | Yes | 2537 | 307 | 976 |
| | No | 5117 | 554 | 2140 |
| Prev CAB | Yes | 188 | 23 | 79 |
| | No | 7465 | 838 | 3037 |
| Prev Valve | Yes | 641 | 74 | 213 |
| | No | 7012 | 787 | 2902 |
| Previous PCI | Yes | 1143 | 136 | 445 |
| | No | 6510 | 725 | 2671 |
| Previous PCI-Within This Episode of Care | Yes | 43 | 7 | 21 |
| | No | 7611 | 854 | 3094 |
| Previous PCI-Interval | > 6 Hours | 1131 | 135 | 435 |
| | <= 6 Hours | 10 | 3 | 8 |
| Previous Other Cardiac | Yes | 1314 | 152 | 501 |
| | No | 6340 | 709 | 2615 |
| Prior MI | Yes | 1526 | 172 | 714 |
| | No | 6127 | 689 | 2397 |
| Heart Failure | Yes | 1625 | 179 | 1042 |
| | No | 6029 | 682 | 2074 |
| Cardiogenic Shock | Yes | 78 | 9 | 83 |
| | No | 7576 | 852 | 3033 |
| Cardiac Arrhythmia | Yes | 1938 | 213 | 912 |
| | No | 5716 | 648 | 2204 |
| Cardiac Arrhythmia - Permanently Paced Rhythm | Yes | 229 | 27 | 90 |
| | No | 1530 | 166 | 822 |
| Cardiac Arrhythmia - VTach / VFib | Yes | 7317 | 832 | 3002 |
| | No | 337 | 29 | 114 |
| Cardiac Arrhythmia - Sick Sinus Syndrome | Yes | 88 | 11 | 85 |
| | No | 1850 | 202 | 827 |
| Cardiac Arrhythmia - AFlutter | Yes | 7448 | 839 | 3037 |
| | No | 206 | 22 | 79 |
| Cardiac Arrhythmia - Atrial Fibrillation | Yes | 5966 | 672 | 2346 |
| | No | 1030 | 126 | 770 |
| Cardiac Arrhythmia - Second Degree Heart Block | Yes | 37 | 8 | 19 |
| | No | 1722 | 185 | 891 |
| Cardiac Arrhythmia - Third Degree Heart Block | Yes | 114 | 11 | 55 |
| | No | 1824 | 202 | 855 |
| Meds-ACE Inhibitors or ARB Within 48 Hours | Yes | 711 | 98 | 523 |
| | No | 6849 | 757 | 2452 |
| Meds- Amiodarone Prior To Surgery | Yes | 248 | 26 | 101 |
| | No | 7399 | 835 | 3012 |
| Meds-Beta Blockers Within 24 Hours | Yes | 4767 | 532 | 2310 |
| | No | 2555 | 289 | 729 |
| Meds-Beta Blocker Therapy For More Than 2 Weeks Prior To Surgery | Yes | 4311 | 487 | 1853 |

Table S3: Class Distribution for AI-derived Variables

| Variable | Value | MGB | | HHC |
|---|---|---|---|---|
| | | Pat. Train | Pat. Test | Pat. Test |
| | No | 3061 | 337 | 1243 |
| Meds-Calcium Channel Blocker Therapy For More Than 2 Weeks Prior To Surgery | Yes | 1439 | 158 | 739 |
| | No | 6209 | 702 | 2374 |
| Meds-Long-Acting Nitrate Therapy For More Than 2 Weeks Prior To Surgery | Yes | 643 | 65 | 222 |
| | No | 6148 | 696 | 2891 |
| Meds-Nitrates-I.V. Within 24 Hours | Yes | 320 | 40 | 48 |
| | No | 7330 | 821 | 3065 |
| Meds-Other Antianginal Medication Therapy For More Than 2 Weeks Prior To Surgery | Yes | 804 | 98 | 30 |
| | No | 6846 | 760 | 3082 |
| Meds-ADP Inhibitors Within Five Days | Yes | 214 | 26 | 109 |
| | No | 7437 | 834 | 2986 |
| Meds-Aspirin Within Five Days | Yes | 4325 | 475 | 1620 |
| | No | 3140 | 357 | 1445 |
| Meds-Aspirin One-Time Dose | Yes | 45 | 5 | 20 |
| | No | 3091 | 344 | 1600 |
| Meds-Anticoagulants Within 48 Hours | Yes | 1234 | 139 | 631 |
| | No | 6420 | 721 | 2482 |
| Meds-Warfarin (Coumadin) Within 5 Days | Yes | 75 | 9 | 41 |
| | No | 4827 | 563 | 3069 |
| Meds-DOAC Within 5 Days | Yes | 18 | 3 | 137 |
| | No | 7636 | 858 | 2979 |
| Meds-Inotropes Within 48 Hours | Yes | 134 | 11 | 69 |
| | No | 7518 | 850 | 3044 |
| Meds-Lipid Lowering Within 24 Hours | Yes | 4066 | 475 | 1630 |
| | No | 3454 | 374 | 1126 |
| Meds-Steroids Within 24 Hours | Yes | 145 | 14 | 83 |
| | No | 6536 | 736 | 3014 |
| Cardiac Catheterization Performed | Yes | 6845 | 775 | 2825 |
| | No | 809 | 86 | 289 |
| Left Main Stenosis >= 50% Known | Yes | 169 | 19 | 440 |
| | No | 613 | 75 | 1346 |
| Circumflex distribution stenosis >= 50% known | Yes | 542 | 68 | 1319 |
| | No | 239 | 26 | 493 |
| Circumflex Distribution Stenosis Percentage | >=70% | 457 | 55 | 1120 |
| | 50 - 69% | 85 | 13 | 199 |
| RCA distribution stenosis >= 50% known | Yes | 618 | 77 | 1358 |
| | No | 1265 | 142 | 456 |
| Hemo Data-Dimensions Available | Yes | 6438 | 717 | 2353 |
| | No | 1215 | 144 | 702 |
| Hemo-PA Systolic Pressure Measured | Yes | 4046 | 426 | 1979 |
| | No | 3605 | 435 | 1125 |
| Aortic Valve Regurgitation | Yes | 837 | 107 | 1590 |
| | No | 6817 | 754 | 1526 |
| VD-Stenosis-Aortic | Yes | 2175 | 245 | 882 |

Table S3: Class Distribution for AI-derived Variables

| Variable | Value | MGB | | HHC |
|---|---|---|---|---|
| | | Pat. Train | Pat. Test | Pat. Test |
| | No | 5479 | 616 | 2234 |
| VD-Aortic Hemodynamic Data Available | Yes | 2126 | 237 | 871 |
| | No | 5528 | 624 | 2244 |
| VD-Aortic | Yes | 2623 | 278 | 1446 |
| | No | 5031 | 583 | 1670 |
| Mitral Valve Regurgitation | Yes | 1412 | 174 | 2606 |
| | No | 6242 | 687 | 510 |
| VD-Stenosis-Mitral | Yes | 363 | 30 | 184 |
| | No | 7291 | 831 | 2932 |
| VD-Mitral Hemodynamic Data Available | Yes | 314 | 27 | 165 |
| | No | 7340 | 834 | 2949 |
| VD-Mitral | Yes | 3047 | 326 | 1848 |
| | No | 4607 | 535 | 1268 |
| Tricuspid Valve Regurgitation | Yes | 1417 | 169 | 2594 |
| | No | 6237 | 692 | 522 |
| VD-Tricuspid Valve Disease | Yes | 2092 | 241 | 1240 |
| | No | 5562 | 620 | 1876 |
| VD-Tricuspid Annular Measurement Available | Yes | 31 | 4 | 72 |
| | No | 3555 | 418 | 1140 |
| Pulmonic Valve Regurgitation | Yes | 1074 | 124 | 1760 |
| | No | 6580 | 737 | 1356 |
| VD-Pulmonic Valve Disease | Yes | 539 | 45 | 306 |
| | No | 7115 | 816 | 2810 |
| **Intra-operative** | | | | |
| CAB | Yes | 3462 | 397 | 1551 |
| | No | 4192 | 464 | 1565 |
| Aorta Procedure Performed | Yes | 815 | 102 | 387 |
| | No | 6839 | 759 | 2729 |
| Surgeon Input for Aortic Surgery Data Abstraction | Yes | 487 | 67 | 385 |
| | No | 6872 | 764 | 2731 |
| Valve | Yes | 4253 | 455 | 1769 |
| | No | 3401 | 406 | 1347 |
| Valve Prosthesis Explant | Yes | 1284 | 143 | 147 |
| | No | 5723 | 639 | 2969 |
| AV-Aorta Procedure Performed | Yes | 118 | 13 | 258 |
| | No | 5280 | 603 | 2858 |
| VS-Mitral Valve | Yes | 1757 | 180 | 828 |
| | No | 5895 | 681 | 2288 |
| VS-Tricuspid Valve | Yes | 362 | 35 | 135 |
| | No | 7288 | 826 | 2981 |
| Surgeon Input for Valve Surgery Data Abstraction | Yes | 2771 | 309 | 1642 |
| | No | 3194 | 380 | 1472 |
| Mechanical Assist Device / Ventricular Assist Device | Yes | 59 | 14 | 155 |

Table S3: Class Distribution for AI-derived Variables

| Variable | Value | MGB | | HHC |
|---|---|---|---|---|
| | | Pat. Train | Pat. Test | Pat. Test |
| | No | 7595 | 847 | 2961 |
| Other Cardiac Procedure, except Afib | Yes | 1109 | 123 | 282 |
| | No | 6545 | 738 | 2834 |
| Atrial Fibrillation Procedure Performed | Yes | 788 | 89 | 524 |
| | No | 6866 | 772 | 2592 |
| Other Cardiac Congenital Except Unicuspid, Bicuspid, or Quadricuspid Valve | Yes | 136 | 16 | 29 |
| | No | 7518 | 845 | 3087 |
| Other Non Card | Yes | 104 | 16 | 67 |
| | No | 7550 | 845 | 3049 |
| General Anesthesia | Yes | 4909 | 575 | 3109 |
| | No | 2745 | 286 | 7 |
| Intubation | Yes | 4909 | 575 | 3104 |
| | No | 2745 | 286 | 12 |
| Appropriate Antibiotic Selection | Yes | 7538 | 851 | 3055 |
| | No | 116 | 10 | 61 |
| Appropriate Antibiotic Administration Timing | Yes | 7503 | 837 | 2869 |
| | No | 151 | 24 | 247 |
| Appropriate Antibiotic Discontinuation | Yes | 7418 | 828 | 3057 |
| | No | 236 | 33 | 59 |
| Temperature Measured | Yes | 4906 | 575 | 2434 |
| | No | 2748 | 286 | 682 |
| Circulatory Arrest | Yes | 451 | 55 | 201 |
| | No | 7203 | 806 | 2915 |
| Circulatory Arrest With Cerebral Perfusion | Yes | 157 | 22 | 78 |
| | No | 7497 | 839 | 3034 |
| Cerebral Oximetry Used | Yes | 723 | 89 | 202 |
| | No | 6931 | 772 | 2914 |
| Intraop Blood Products | Yes | 1781 | 201 | 1118 |
| | No | 5873 | 660 | 1998 |
| Intraop TEE post procedure | Yes | 6611 | 725 | 3012 |
| | No | 1043 | 136 | 104 |
| Ejection Fraction Measured Post Procedure | Yes | 1778 | 220 | 2875 |
| | No | 3579 | 419 | 235 |
| Internal Mammary Artery Used | Yes | 2061 | 239 | 1372 |
| | No | 4397 | 492 | 1744 |
| Left IMA Used | Yes | 1521 | 176 | 1366 |
| | No | 5612 | 624 | 1750 |
| Right IMA Used | Yes | 282 | 27 | 56 |
| | No | 6851 | 773 | 3059 |
| Distal Anastomoses with Arterial Conduit(s) | Yes | 1300 | 149 | 1382 |
| | No | 4463 | 492 | 1734 |
| Distal Anastomoses with Radial Artery Conduit(s) Used | Yes | 129 | 15 | 471 |
| | No | 7387 | 829 | 2645 |
| Distal Anastomoses with Venous Conduit(s) Used | Yes | 2013 | 237 | 1355 |

Table S3: Class Distribution for AI-derived Variables

| Variable | Value | MGB | | HHC |
|---|---|---|---|---|
| | | Pat. Train | Pat. Test | Pat. Test |
| | No | 4444 | 494 | 1761 |
| CAB Distal Position 01 | Side to Side | 70 | 11 | 39 |
| | End to Side | 3389 | 385 | 1512 |
| CAB Endarterectomy 01 | Yes | 61 | 9 | 5 |
| | No | 7593 | 852 | 3111 |
| CAB 02 | Yes | 2971 | 340 | 1340 |
| | No | 4683 | 521 | 1776 |
| CAB Distal Position 02 | Yes | 2681 | 309 | 1253 |
| | No | 4971 | 552 | 1863 |
| CAB 03 | Yes | 2226 | 247 | 982 |
| | No | 5428 | 614 | 2134 |
| CAB Distal Position 03 | Yes | 1955 | 209 | 917 |
| | No | 5698 | 652 | 2199 |
| CAB Endarterectomy 03 | Yes | 28 | 3 | 3 |
| | No | 2196 | 246 | 979 |
| CAB 04 | Yes | 947 | 125 | 369 |
| | No | 6707 | 736 | 2747 |
| CAB Distal Position 04 | Side to Side | 121 | 12 | 28 |
| | End to Side | 826 | 113 | 341 |
| CAB 05 | Yes | 201 | 24 | 61 |
| | No | 7453 | 837 | 3055 |
| First Valve Explant Device Known | Yes | 243 | 27 | 96 |
| | No | 6480 | 731 | 3018 |
| Second Valve Prosthesis Explant | Yes | 21 | 2 | 9 |
| | No | 6702 | 756 | 3104 |
| IABP | Yes | 172 | 23 | 113 |
| | No | 7482 | 838 | 3001 |
| Temporary Assist Device Used | Yes | 25 | 2 | 9 |
| | No | 7629 | 859 | 3105 |
| VAD-Patient Admitted With VAD | Yes | 78 | 12 | 2 |
| | No | 7576 | 849 | 3112 |
| Previous VAD Explanted During This Admission | Yes | 78 | 12 | 2 |
| | No | 7576 | 849 | 3114 |
| Ventricular Assist Device Implanted During This Hospitalization | Yes | 43 | 7 | 24 |
| | No | 7611 | 854 | 3090 |
| VAD-Explant | Yes | 16 | 7 | 9 |
| | No | 7638 | 854 | 3073 |
| Other Card-Pulmonary Thromboembolectomy | Yes | 90 | 4 | 3 |
| | No | 1667 | 176 | 279 |
| Other Card-Lead Insertion | Yes | 45 | 8 | 39 |
| | No | 7608 | 853 | 3077 |
| Other Card-Card Tx | Yes | 133 | 17 | 34 |
| | No | 7521 | 844 | 3082 |
| Other Card-Other | Yes | 201 | 22 | 60 |

Table S3: Class Distribution for AI-derived Variables

| Variable | Value | MGB | | HHC |
|---|---|---|---|---|
| | | Pat. Train | Pat. Test | Pat. Test |
| | No | 7453 | 839 | 3056 |
| Lesions Documented | Yes | 648 | 74 | 397 |
| | No | 6953 | 781 | 2719 |
| Prior Aortic Intervention | Yes | 79 | 11 | 57 |
| | No | 7279 | 820 | 3059 |
| Prior Aortic Intervention - Previous Repair - Ascending (Zone 0 - B&C) | Yes | 58 | 4 | 38 |
| | No | 7595 | 857 | 3078 |
| Prior Aortic Intervention - Previous Repair - Arch (Zones 1,2,3) | Yes | 25 | 3 | 22 |
| | No | 7629 | 858 | 3094 |
| Prior Aortic Intervention - Previous Repair - Descending (Zones 4,5) | Yes | 17 | 3 | 16 |
| | No | 7634 | 860 | 3100 |
| Prior Aortic Intervention - Previous Repair - Infrarenal Abdominal (Zones 8,9,10,11) | Yes | 4 | 3 | 11 |
| | No | 7651 | 860 | 3105 |
| Root - Asymmetric Root Dilation | Yes | 12 | 3 | 2 |
| | No | 7236 | 813 | 2880 |
| Root - Sinus Of Valsalva Aneurysm | Yes | 95 | 6 | 10 |
| | No | 7187 | 810 | 2898 |
| Ascending Proximal Coronary Bypass Grafts | Yes | 17 | 3 | 12 |
| | No | 7316 | 824 | 3052 |
| Proximal to Treated Zone(s) (Largest Diameter) Available | Yes | 51 | 8 | 135 |
| | No | 6957 | 775 | 2923 |
| Distal to Treated Zone(s) (Largest Diameter) Available | Yes | 64 | 5 | 171 |
| | No | 6944 | 778 | 2886 |
| Surgical Ascending/Arch Procedure | Yes | 367 | 49 | 293 |
| | No | 6973 | 780 | 2823 |
| Open Arch Procedure - Arch Branch Reimplantation | Yes | 35 | 5 | 28 |
| | No | 7595 | 850 | 3088 |
| Surgical Descending Thoracic Aorta or Thoracoabdominal Procedure | Yes | 971 | 112 | 12 |
| | No | 6369 | 718 | 3097 |
| Planned Staged Hybrid | Yes | 5 | 3 | 2 |
| | No | 7647 | 860 | 3114 |
| Spinal Drain | Yes | 22 | 4 | 15 |
| | No | 7336 | 827 | 3100 |
| Aorta Device Inserted | Yes | 477 | 69 | 382 |
| | No | 6880 | 762 | 2734 |
| Aortic Valve or Aortic Valve Composite Graft Implanted | Yes | 85 | 12 | 205 |
| | No | 7256 | 804 | 2911 |
| **Post-operative** | | | | |
| Blood Prod | Yes | 1639 | 176 | 1061 |
| | No | 5048 | 574 | 2055 |
| Extubated In OR | Yes | 121 | 17 | 89 |
| | No | 7533 | 844 | 3027 |
| Postop Intubation/Reintubation During Hospital Stay | Yes | 126 | 12 | 64 |

Table S3: Class Distribution for AI-derived Variables

| Variable | Value | MGB | | HHC |
|---|---|---|---|---|
| | | Pat. Train | Pat. Test | Pat. Test |
| | No | 4784 | 563 | 3052 |
| ICU Visit | Yes | 6673 | 748 | 3043 |
| | No | 981 | 113 | 73 |
| Readmission to ICU | Yes | 181 | 18 | 59 |
| | No | 7473 | 843 | 3057 |
| Postop Echo | Yes | 1284 | 137 | 624 |
| | No | 6370 | 724 | 2492 |
| Postop EF Done | Yes | 1395 | 149 | 632 |
| | No | 6259 | 712 | 2484 |
| Post-Op-Surgical Site Infection | Yes | 26 | 2 | 54 |
| | No | 7628 | 859 | 3062 |
| Post-Op-Deep Sternal | Yes | 20 | 2 | 4 |
| | No | 7628 | 857 | 3099 |
| In Hospital Post-Op Events | Yes | 2854 | 320 | 1223 |
| | No | 4800 | 541 | 1893 |
| Post-Op-ReOp Bleeding/Tampanade | Yes | 181 | 18 | 79 |
| | No | 7473 | 843 | 3037 |
| Post-Op-Unplanned Coronary Artery Intervention | Yes | 12 | 2 | 6 |
| | No | 6848 | 784 | 3110 |
| Post-Op-ReOp Other Cardiac Reasons | Yes | 42 | 6 | 17 |
| | No | 7612 | 855 | 3099 |
| Post-Op-Return To OR For Other Non-cardiac Reason | Yes | 135 | 8 | 58 |
| | No | 7519 | 853 | 3058 |
| Post-Op-Open Chest With Planned Delayed Sternal Closure | Yes | 87 | 7 | 25 |
| | No | 7567 | 854 | 3091 |
| Post-Op-Sepsis | Yes | 45 | 7 | 17 |
| | No | 7609 | 854 | 3099 |
| Post-Op-Neuro-Stroke Perm | Yes | 24 | 2 | 47 |
| | No | 7549 | 847 | 3069 |
| Post-Op-Recurrent Laryngeal Nerve Injury | Yes | 33 | 8 | 7 |
| | No | 7621 | 853 | 3109 |
| Post-Op-Pulm-Vent Prolonged | Yes | 513 | 52 | 189 |
| | No | 7140 | 809 | 2927 |
| Post-Op-Pulm-Pneumonia | Yes | 204 | 16 | 67 |
| | No | 7450 | 845 | 3049 |
| Post-Op-Pleural Effusion Requiring Drainage | Yes | 318 | 43 | 94 |
| | No | 7336 | 818 | 3022 |
| Post-Op-Renal-Renal Failure | Yes | 136 | 9 | 55 |
| | No | 7518 | 852 | 3060 |
| Post-Op-Vasc-Acute Limb Ischemia | Yes | 17 | 3 | 8 |
| | No | 7635 | 860 | 3108 |
| Post-Op-Deep Venous Thrombosis | Yes | 93 | 7 | 39 |
| | No | 5416 | 626 | 3077 |
| Post-Op-New Dysrhythmia Requiring Insertion of a Permanent Device | Yes | 75 | 7 | 137 |

Table S3: Class Distribution for AI-derived Variables

| Variable | Value | MGB | | HHC |
|---|---|---|---|---|
| | | Pat. Train | Pat. Test | Pat. Test |
| | No | 5361 | 622 | 2978 |
| Post-Op-Other-Cardiac Arrest | Yes | 63 | 5 | 37 |
| | No | 7591 | 856 | 3078 |
| Post-Op-Other-Anticoagulant Bleeding Event | Yes | 56 | 3 | 3 |
| | No | 7598 | 858 | 3112 |
| Post-Op-Other-Gastro-Intestinal Event | Yes | 124 | 13 | 87 |
| | No | 7184 | 811 | 3028 |
| Post-Op-Other-A Fib | Yes | 2088 | 227 | 851 |
| | No | 5566 | 634 | 2264 |

Value: The post-processed representation of each target variable, assigned after binarization. As described in the Supplementary Methods, categorical variables with multiple levels are collapsed into binary form to enhance model interpretability and performance.
MGB Pat. Train/Test.: The number of patients in the Massachusetts General Brigham (MGB) cohort (out of 8,515) that were used for training/testing of the models and for whom the given variable was present and assigned the corresponding binarized value.
HHC Pat. Test: The number of patients in the Hartford Healthcare (HHC) cohort (out of 3,116) that were used for external validation and for whom the given variable was present and assigned the corresponding binarized value.

### 3 Pipeline Performance Results

#### 3.1 Performance after Confidence Intervals

Table S4: Performance of AI-derived variables after Confidence Intervals

| | MGB | | | | HHC | | | |
|---|---|---|---|---|---|---|---|---|
| Variable | AUC | Pr. | Rec. | Sp. | AUC | Pr. | Rec. | Sp. |
| **Pre-operative** | | | | | | | | |
| RF-Family History of Premature CAD | 0.694 | 0.24 | 0.667 | 0.661 | 0.816 | 0.218 | 0.75 | 0.823 |
| RF-Diabetes | 0.98 | 0.897 | 0.939 | 0.96 | 0.984 | 0.904 | 0.96 | 0.951 |
| RF-Renal Fail-Dialysis | 1.0 | 1.0 | 1.0 | 1.0 | 0.999 | 0.362 | 1.0 | 0.946 |
| RF-Hypertension | 0.957 | 0.792 | 0.952 | 0.924 | 0.95 | 0.691 | 0.926 | 0.916 |
| RF- Endocarditis | 0.985 | 0.448 | 0.963 | 0.938 | 0.987 | 0.586 | 0.944 | 0.974 |
| RF-Pulmonary Function Test | 0.751 | 0.526 | 1.0 | 0.0 | 0.802 | 0.507 | 0.878 | 0.715 |
| RF-Arterial Blood Gas | 1.0 | 1.0 | 1.0 | 1.0 | 1.0 | 0.0 | 1.0 | 0.929 |
| RF-Home Oxygen | 0.988 | 0.22 | 1.0 | 0.928 | 0.859 | 0.109 | 1.0 | 0.75 |
| RF-Inhaled Medication or Oral Bronchodilator Therapy | 0.957 | 0.6 | 0.913 | 0.93 | 0.962 | 0.719 | 0.962 | 0.924 |
| RF-Sleep Apnea | 0.971 | 0.808 | 0.933 | 0.947 | 0.969 | 0.841 | 0.956 | 0.943 |
| RF-Pneumonia | 0.916 | 0.519 | 0.871 | 0.86 | 0.734 | 0.439 | 0.829 | 0.664 |
| RF-Illicit Drug Use within One Year | 1.0 | 1.0 | 1.0 | 1.0 | 0.927 | 0.318 | 0.875 | 0.908 |
| RF-Liver Disease | 0.952 | 0.432 | 0.95 | 0.927 | 0.971 | 0.423 | 0.938 | 0.954 |
| RF-Liver Cirrhosis | 0.1 | 0.167 | 1.0 | 0.0 | 0.99 | 0.2 | 1.0 | 0.972 |

Table S4: Performance of AI-derived variables after Confidence Intervals

| | MGB | | | | HHC | | | |
|---|---|---|---|---|---|---|---|---|
| Variable | AUC | Pr. | Rec. | Sp. | AUC | Pr. | Rec. | Sp. |
| Immunocompromised Present | 0.969 | 0.486 | 0.947 | 0.952 | 0.92 | 0.391 | 0.862 | 0.914 |
| Mediastinal Radiation | 0.992 | 0.364 | 1.0 | 0.982 | 0.991 | 0.357 | 0.952 | 0.964 |
| Cancer Within 5 Years | 0.919 | 0.99 | 0.947 | 0.84 | 0.918 | 0.992 | 0.892 | 0.919 |
| Peripheral Artery Disease | 0.957 | 0.583 | 0.913 | 0.895 | 0.936 | 0.645 | 0.973 | 0.856 |
| Syncope | 0.932 | 0.375 | 1.0 | 0.824 | 0.784 | 0.206 | 0.812 | 0.719 |
| Cerebrovascular Disease | 0.972 | 0.753 | 0.961 | 0.923 | 0.965 | 0.776 | 0.95 | 0.943 |
| Prior CVA | 0.982 | 0.65 | 0.975 | 0.949 | 0.973 | 0.761 | 0.922 | 0.967 |
| CVD Prior Carotid Surgery | 0.982 | 0.194 | 1.0 | 0.918 | 0.976 | 0.25 | 0.933 | 0.949 |
| HIT Antibodies | 0.951 | 0.067 | 1.0 | 0.949 | 1.0 | 1.0 | 1.0 | 1.0 |
| Five Meter Walk Test Done | 1.0 | 0.0 | 1.0 | 0.934 | 0.822 | 0.128 | 0.714 | 0.864 |
| Prev Cardiac Interventions | 0.978 | 0.906 | 0.938 | 0.934 | 0.977 | 0.887 | 0.961 | 0.942 |
| Prev CAB | 0.96 | 0.421 | 0.941 | 0.953 | 0.95 | 0.34 | 0.944 | 0.961 |
| Prev Valve | 0.991 | 0.72 | 0.982 | 0.956 | 0.985 | 0.685 | 0.926 | 0.967 |
| Previous PCI | 0.977 | 0.798 | 0.926 | 0.96 | 0.986 | 0.787 | 0.921 | 0.96 |
| Previous PCI-Within This Episode of Care | 0.985 | 0.4 | 0.8 | 0.989 | 0.972 | 0.0 | 0.0 | 0.983 |
| Previous PCI-Interval | 1.0 | 0.333 | 1.0 | 0.996 | 1.0 | 0.0 | 0.0 | 0.999 |
| Previous Other Cardiac | 0.968 | 0.806 | 0.929 | 0.935 | 0.95 | 0.773 | 0.908 | 0.956 |
| Prior MI | 0.971 | 0.81 | 0.964 | 0.935 | 0.96 | 0.877 | 0.934 | 0.958 |
| Heart Failure | 0.952 | 0.726 | 0.92 | 0.896 | 0.925 | 0.844 | 0.894 | 0.915 |
| Cardiogenic Shock | 0.973 | 0.194 | 1.0 | 0.943 | 0.976 | 0.471 | 0.941 | 0.962 |
| Cardiac Arrhythmia | 0.957 | 0.882 | 0.944 | 0.934 | 0.976 | 0.881 | 0.955 | 0.928 |
| Cardiac Arrhythmia - Permanently Paced Rhythm | 1.0 | 0.684 | 1.0 | 0.983 | 1.0 | 0.35 | 0.778 | 0.977 |
| Cardiac Arrhythmia - VTach / VFib | 0.92 | 0.378 | 0.773 | 0.929 | 0.929 | 0.5 | 0.842 | 0.942 |
| Cardiac Arrhythmia - Sick Sinus Syndrome | 1.0 | 0.4 | 1.0 | 0.99 | 1.0 | 0.478 | 0.846 | 0.976 |
| Cardiac Arrhythmia - AFlutter | 0.981 | 0.467 | 0.875 | 0.967 | 0.972 | 0.245 | 1.0 | 0.951 |
| Cardiac Arrhythmia - Atrial Fibrillation | 0.976 | 0.814 | 0.954 | 0.952 | 0.974 | 0.875 | 0.939 | 0.951 |
| Cardiac Arrhythmia - Second Degree Heart Block | 1.0 | 1.0 | 1.0 | 1.0 | 1.0 | 0.0 | 0.0 | 0.998 |
| Cardiac Arrhythmia - Third Degree Heart Block | 1.0 | 0.25 | 1.0 | 0.982 | 1.0 | 0.333 | 0.8 | 0.973 |
| Meds-ACE Inhibitors or ARB Within 48 Hours | 0.362 | 0.0 | 0.0 | 0.93 | 0.667 | 0.255 | 0.6 | 0.679 |
| Meds- Amiodarone Prior To Surgery | 0.698 | 0.31 | 1.0 | 0.0 | 0.896 | 0.212 | 0.846 | 0.903 |
| Meds-Beta Blockers Within 24 Hours | 0.952 | 0.953 | 0.933 | 0.929 | 1.0 | 1.0 | 1.0 | 1.0 |
| Meds-Beta Blocker Therapy For More Than 2 Weeks Prior To Surgery | 0.924 | 0.902 | 0.895 | 0.898 | 1.0 | 1.0 | 1.0 | 1.0 |
| Meds-Calcium Channel Blocker Therapy For More Than 2 Weeks Prior To Surgery | 0.953 | 0.744 | 0.955 | 0.924 | 0.842 | 0.661 | 0.891 | 0.807 |
| Meds-Long-Acting Nitrate Therapy For More Than 2 Weeks Prior To Surgery | 0.983 | 0.635 | 0.943 | 0.959 | 0.969 | 0.317 | 0.905 | 0.932 |
| Meds-Nitrates-I.V. Within 24 Hours | 0.972 | 0.375 | 0.857 | 0.953 | 0.958 | 0.108 | 1.0 | 0.914 |
| Meds-Other Antianginal Medication Therapy For More Than 2 Weeks Prior To Surgery | 0.942 | 0.0 | 0.0 | 0.978 | 1.0 | 0.0 | 1.0 | 0.997 |

Table S4: Performance of AI-derived variables after Confidence Intervals

| | MGB | | | | HHC | | | |
|---|---|---|---|---|---|---|---|---|
| Variable | AUC | Pr. | Rec. | Sp. | AUC | Pr. | Rec. | Sp. |
| Meds-ADP Inhibitors Within Five Days | 0.961 | 0.571 | 0.571 | 0.993 | 0.669 | 0.22 | 1.0 | 0.0 |
| Meds-Aspirin Within Five Days | 0.952 | 0.936 | 0.936 | 0.916 | 0.931 | 0.902 | 0.909 | 0.909 |
| Meds-Aspirin One-Time Dose | 1.0 | 0.5 | 1.0 | 0.996 | 1.0 | 0.125 | 0.667 | 0.957 |
| Meds-Anticoagulants Within 48 Hours | 0.952 | 0.703 | 0.951 | 0.909 | 0.928 | 0.713 | 0.838 | 0.94 |
| Meds-Warfarin (Coumadin) Within 5 Days | 0.95 | 0.161 | 1.0 | 0.904 | 0.872 | 0.053 | 1.0 | 0.745 |
| Meds-DOAC Within 5 Days | 1.0 | 1.0 | 1.0 | 1.0 | 0.639 | 0.13 | 0.778 | 0.489 |
| Meds-Thrombolytics Within 24 Hours | 1.0 | 1.0 | 1.0 | 1.0 | 1.0 | 1.0 | 1.0 | 1.0 |
| Meds-Inotropes Within 48 Hours | 0.986 | 0.333 | 1.0 | 0.953 | 0.983 | 0.426 | 0.909 | 0.966 |
| Meds-Lipid Lowering Within 24 Hours | 0.943 | 0.921 | 0.888 | 0.918 | 0.889 | 0.872 | 0.767 | 0.917 |
| Meds-Steroids Within 24 Hours | 0.994 | 0.31 | 1.0 | 0.957 | 0.755 | 0.188 | 1.0 | 0.0 |
| Cardiac Catheterization Performed | 0.945 | 1.0 | 0.865 | 1.0 | 0.95 | 0.988 | 0.927 | 0.88 |
| Left Main Stenosis >= 50Circumflex distribution stenosis >= 50Circumflex Distribution Stenosis Percentage | 1.0 | 0.0 | 0.0 | 0.993 | 1.0 | 0.333 | 0.111 | 0.995 |
| RCA distribution stenosis >= 50Hemo Data-Dimensions Available | 0.798 | 0.964 | 0.829 | 0.667 | 0.524 | 1.0 | 0.0 | 1.0 |
| Hemo-PA Systolic Pressure Measured | 0.837 | 0.79 | 0.731 | 0.865 | 1.0 | 1.0 | 1.0 | 1.0 |
| Aortic Valve Regurgitation | 0.977 | 0.654 | 0.919 | 0.959 | 0.631 | 0.731 | 0.782 | 0.574 |
| VD-Stenosis-Aortic | 0.979 | 0.879 | 0.929 | 0.954 | 0.954 | 0.848 | 0.951 | 0.912 |
| VD-Aortic Hemodynamic Data Available | 0.97 | 0.889 | 0.934 | 0.96 | 0.963 | 0.856 | 0.954 | 0.936 |
| VD-Aortic | 0.959 | 0.893 | 0.954 | 0.874 | 0.923 | 0.924 | 0.939 | 0.869 |
| Mitral Valve Regurgitation | 0.996 | 0.962 | 0.962 | 0.991 | 0.627 | 1.0 | 0.0 | 1.0 |
| VD-Stenosis-Mitral | 0.989 | 0.472 | 1.0 | 0.941 | 0.975 | 0.5 | 0.964 | 0.927 |
| VD-Mitral Hemodynamic Data Available | 0.993 | 0.421 | 1.0 | 0.952 | 0.969 | 0.436 | 0.919 | 0.948 |
| VD-Mitral | 0.94 | 0.862 | 0.951 | 0.74 | 0.907 | 0.94 | 0.912 | 0.798 |
| Tricuspid Valve Regurgitation | 0.992 | 0.952 | 0.971 | 0.989 | 1.0 | 1.0 | 1.0 | 1.0 |
| VD-Tricuspid Valve Disease | 0.735 | 0.685 | 0.924 | 0.491 | 0.834 | 0.76 | 0.821 | 0.758 |
| VD-Tricuspid Annular Measurement Available | 1.0 | 0.0 | 1.0 | 0.831 | 1.0 | 0.15 | 0.6 | 0.898 |
| Pulmonic Valve Regurgitation | 0.969 | 0.738 | 0.923 | 0.962 | 1.0 | 1.0 | 1.0 | 1.0 |
| VD-Pulmonic Valve Stenosis | 0.985 | 0.182 | 1.0 | 0.98 | 1.0 | 1.0 | 1.0 | 1.0 |
| VD-Pulmonic Valve Disease | 0.808 | 0.294 | 0.938 | 0.561 | 0.5 | 0.118 | 0.545 | 0.43 |
| **Intra-operative** | | | | | | | | |
| Robot Used | 1.0 | 1.0 | 1.0 | 1.0 | 1.0 | 1.0 | 1.0 | 1.0 |
| CAB | 0.999 | 0.991 | 0.94 | 0.994 | 0.994 | 0.969 | 0.959 | 0.969 |
| Aorta Procedure Performed | 0.982 | 0.832 | 0.952 | 0.955 | 0.975 | 0.714 | 0.96 | 0.94 |
| Surgeon Input for Aortic Surgery Data Abstraction | 0.983 | 0.821 | 0.928 | 0.969 | 0.975 | 0.764 | 0.958 | 0.959 |
| Valve | 0.976 | 0.967 | 0.951 | 0.959 | 1.0 | 1.0 | 1.0 | 1.0 |
| Valve Prosthesis Explant | 0.988 | 0.8 | 0.875 | 0.987 | 0.966 | 0.5 | 0.977 | 0.941 |

Table S4: Performance of AI-derived variables after Confidence Intervals

| | MGB | | | | HHC | | | |
|---|---|---|---|---|---|---|---|---|
| Variable | AUC | Pr. | Rec. | Sp. | AUC | Pr. | Rec. | Sp. |
| AV-Aorta Procedure Performed | 1.0 | 0.545 | 0.923 | 0.974 | 0.96 | 0.533 | 0.934 | 0.93 |
| VS-Mitral Valve | 0.997 | 0.966 | 0.942 | 0.991 | 0.994 | 0.938 | 0.959 | 0.978 |
| VS-Tricuspid Valve | 0.996 | 0.781 | 0.893 | 0.987 | 0.991 | 0.468 | 0.974 | 0.956 |
| VS-Pulmonic Valve | 0.999 | 0.786 | 1.0 | 0.994 | 1.0 | 1.0 | 1.0 | 1.0 |
| Surgeon Input for Valve Surgery Data Abstraction | 1.0 | 0.92 | 0.905 | 0.945 | 0.969 | 0.917 | 0.951 | 0.929 |
| Mechanical Assist Device / Ventricular Assist Device | 0.997 | 0.4 | 1.0 | 0.974 | 0.958 | 0.463 | 0.962 | 0.922 |
| Other Cardiac Procedure, except Afib | 0.962 | 0.702 | 0.952 | 0.908 | 0.941 | 0.516 | 0.904 | 0.921 |
| Atrial Fibrillation Procedure Performed | 0.989 | 0.797 | 0.937 | 0.969 | 0.99 | 0.771 | 0.969 | 0.941 |
| Other Cardiac Congenital Except Unicuspid, Bicuspid, or Quadricuspid Valve | 0.978 | 0.25 | 1.0 | 0.915 | 0.963 | 0.123 | 1.0 | 0.889 |
| Other Non Card | 0.973 | 0.2 | 1.0 | 0.901 | 0.774 | 0.048 | 0.667 | 0.788 |
| General Anesthesia | 0.999 | 1.0 | 0.984 | 1.0 | 1.0 | 1.0 | 0.841 | 1.0 |
| Intubation | 0.998 | 1.0 | 0.941 | 1.0 | 0.944 | 1.0 | 0.859 | 1.0 |
| Appropriate Antibiotic Selection | 0.529 | 1.0 | 0.412 | 1.0 | 0.533 | 1.0 | 0.0 | 1.0 |
| Appropriate Antibiotic Administration Timing | 1.0 | 1.0 | 0.975 | 1.0 | 1.0 | 1.0 | 1.0 | 1.0 |
| Appropriate Antibiotic Discontinuation | 0.934 | 0.997 | 0.935 | 0.889 | 0.514 | 1.0 | 0.0 | 1.0 |
| Temperature Measured | 0.999 | 1.0 | 0.963 | 1.0 | 1.0 | 1.0 | 1.0 | 1.0 |
| Circulatory Arrest | 0.98 | 0.614 | 0.962 | 0.932 | 0.969 | 0.486 | 0.944 | 0.941 |
| Circulatory Arrest With Cerebral Perfusion | 0.987 | 0.435 | 0.952 | 0.95 | 0.944 | 0.209 | 0.824 | 0.945 |
| Cerebral Oximetry Used | 0.957 | 0.627 | 0.981 | 0.875 | 0.883 | 0.361 | 0.897 | 0.882 |
| Intraop Blood Products | 0.888 | 0.691 | 0.944 | 0.79 | 0.842 | 0.75 | 0.765 | 0.867 |
| Intraop TEE post procedure | 0.5 | 1.0 | 0.0 | 1.0 | 0.399 | 1.0 | 0.0 | 1.0 |
| Ejection Fraction Measured Post Procedure | 1.0 | 0.722 | 0.703 | 0.965 | 0.657 | 1.0 | 0.0 | 1.0 |
| Internal Mammary Artery Used | 1.0 | 0.985 | 0.941 | 0.994 | 0.991 | 0.955 | 0.957 | 0.966 |
| Left IMA Used | 0.992 | 0.925 | 0.925 | 0.976 | 0.98 | 0.962 | 0.925 | 0.974 |
| Right IMA Used | 0.982 | 0.421 | 0.96 | 0.937 | 0.992 | 0.31 | 1.0 | 0.957 |
| Distal Anastomoses with Arterial Conduit(s) | 1.0 | 0.964 | 0.931 | 0.997 | 0.98 | 0.94 | 0.935 | 0.957 |
| Distal Anastomoses with Radial Artery Conduit(s) Used | 0.994 | 0.462 | 1.0 | 0.987 | 0.971 | 0.706 | 0.932 | 0.941 |
| Distal Anastomoses with Venous Conduit(s) Used | 1.0 | 0.976 | 0.917 | 0.991 | 0.987 | 0.937 | 0.937 | 0.952 |
| CAB Distal Position 01 | 1.0 | 1.0 | 0.729 | 1.0 | 1.0 | 1.0 | 0.644 | 1.0 |
| CAB Endarterectomy 01 | 0.97 | 0.091 | 1.0 | 0.975 | 1.0 | 0.0 | 1.0 | 0.971 |
| CAB 02 | 0.997 | 0.978 | 0.923 | 0.989 | 0.991 | 0.941 | 0.948 | 0.959 |
| CAB Distal Position 02 | 0.986 | 0.939 | 0.895 | 0.973 | 0.978 | 0.922 | 0.917 | 0.958 |
| CAB 03 | 0.985 | 0.88 | 0.926 | 0.958 | 0.988 | 0.901 | 0.926 | 0.959 |
| CAB Distal Position 03 | 0.982 | 0.871 | 0.886 | 0.964 | 0.975 | 0.876 | 0.898 | 0.956 |
| CAB Endarterectomy 03 | 1.0 | 0.333 | 1.0 | 0.996 | 1.0 | 0.0 | 1.0 | 0.999 |
| CAB 04 | 0.989 | 0.901 | 0.948 | 0.983 | 0.983 | 0.793 | 0.881 | 0.97 |

Table S4: Performance of AI-derived variables after Confidence Intervals

| | MGB | | | | HHC | | | |
|---|---|---|---|---|---|---|---|---|
| Variable | AUC | Pr. | Rec. | Sp. | AUC | Pr. | Rec. | Sp. |
| CAB Distal Position 04 | 1.0 | 1.0 | 0.75 | 1.0 | 1.0 | 1.0 | 0.0 | 1.0 |
| CAB 05 | 0.969 | 0.214 | 0.857 | 0.955 | 0.964 | 0.238 | 0.833 | 0.964 |
| First Valve Explant Device Known | 0.982 | 0.613 | 0.905 | 0.977 | 0.957 | 0.444 | 0.933 | 0.962 |
| Second Valve Prosthesis Explant | 1.0 | 1.0 | 1.0 | 1.0 | 0.968 | 0.045 | 1.0 | 0.979 |
| IABP | 0.968 | 0.25 | 0.333 | 0.977 | 1.0 | 0.375 | 0.9 | 0.957 |
| Temporary Assist Device Used | 0.973 | 0.1 | 1.0 | 0.967 | 1.0 | 0.056 | 1.0 | 0.964 |
| VAD-Patient Admitted With VAD | 0.994 | 0.571 | 1.0 | 0.983 | 1.0 | 1.0 | 1.0 | 1.0 |
| Previous VAD Explanted During This Admission | 0.995 | 0.632 | 1.0 | 0.987 | 1.0 | 1.0 | 1.0 | 1.0 |
| Ventricular Assist Device Implanted During This Hospitalization | 0.992 | 0.444 | 0.889 | 0.982 | 1.0 | 0.222 | 0.889 | 0.973 |
| VAD-Explant | 0.998 | 0.636 | 1.0 | 0.993 | 1.0 | 0.095 | 1.0 | 0.981 |
| Other Card-Pulmonary Thromboembolectomy | 1.0 | 0.25 | 0.667 | 0.987 | 1.0 | 1.0 | 1.0 | 1.0 |
| Other Card-Lead Insertion | 0.996 | 0.5 | 1.0 | 0.989 | 0.988 | 0.27 | 1.0 | 0.971 |
| Other Card-Card Tx | 0.998 | 0.81 | 1.0 | 0.993 | 0.997 | 0.278 | 0.833 | 0.974 |
| Other Card-Other | 0.865 | 0.282 | 0.688 | 0.933 | 0.765 | 0.086 | 0.5 | 0.949 |
| Atrial Fibrillation - Lesion Creation Radiofrequency - Bipolar | 1.0 | 1.0 | 1.0 | 1.0 | 1.0 | 1.0 | 1.0 | 1.0 |
| Lesions Documented | 0.983 | 0.836 | 0.962 | 0.98 | 0.989 | 0.772 | 0.953 | 0.96 |
| Prior Aortic Intervention | 0.976 | 0.279 | 1.0 | 0.938 | 0.966 | 0.333 | 0.941 | 0.965 |
| Prior Aortic Intervention - Previous Repair - Ascending (Zone 0 - B&C) | 0.973 | 0.192 | 1.0 | 0.962 | 0.977 | 0.295 | 0.929 | 0.968 |
| Prior Aortic Intervention - Previous Repair - Arch (Zones 1,2,3) | 0.983 | 0.062 | 1.0 | 0.94 | 0.986 | 0.148 | 1.0 | 0.955 |
| Prior Aortic Intervention - Previous Repair - Descending (Zones 4,5) | 0.996 | 0.333 | 1.0 | 0.993 | 0.994 | 0.119 | 1.0 | 0.962 |
| Prior Aortic Intervention - Previous Repair - Infrarenal Abdominal (Zones 8,9,10,11) | 1.0 | 1.0 | 1.0 | 1.0 | 0.996 | 0.1 | 1.0 | 0.981 |
| Current Procedure with Endoleak involvement | 1.0 | 1.0 | 1.0 | 1.0 | 1.0 | 0.0 | 1.0 | 0.987 |
| Current Procedure with Aorta Infection | 1.0 | 1.0 | 1.0 | 1.0 | 1.0 | 0.143 | 1.0 | 0.993 |
| Root - Aortic-Annular Ectasia | 1.0 | 0.211 | 1.0 | 0.97 | 1.0 | 1.0 | 1.0 | 1.0 |
| Root - Asymmetric Root Dilation | 1.0 | 1.0 | 1.0 | 1.0 | 1.0 | 1.0 | 1.0 | 1.0 |
| Root - Sinus Of Valsalva Aneurysm | 1.0 | 0.194 | 1.0 | 0.942 | 1.0 | 0.0 | 1.0 | 0.965 |
| Ascending Proximal Coronary Bypass Grafts | 0.969 | 0.2 | 0.5 | 0.991 | 1.0 | 0.0 | 0.0 | 0.99 |
| Proximal to Treated Zone(s) (Largest Diameter) Available | 1.0 | 0.421 | 1.0 | 0.976 | 1.0 | 0.447 | 0.971 | 0.955 |
| Distal to Treated Zone(s) (Largest Diameter) Available | 1.0 | 0.179 | 1.0 | 0.951 | 1.0 | 0.516 | 0.942 | 0.95 |
| Surgical Ascending/Arch Procedure | 0.972 | 0.606 | 0.915 | 0.94 | 0.979 | 0.628 | 0.942 | 0.947 |
| Open Arch Procedure - Arch Branch Reimplantation | 1.0 | 0.462 | 1.0 | 0.987 | 0.986 | 0.28 | 0.875 | 0.981 |
| Surgical Descending Thoracic Aorta or Thoracoabdominal Procedure | 0.938 | 1.0 | 0.0 | 1.0 | 0.977 | 0.15 | 1.0 | 0.981 |
| Planned Staged Hybrid | 0.998 | 0.5 | 1.0 | 0.998 | 0.983 | 0.105 | 1.0 | 0.983 |

Table S4: Performance of AI-derived variables after Confidence Intervals

| | MGB | | | | HHC | | | |
|---|---|---|---|---|---|---|---|---|
| Variable | AUC | Pr. | Rec. | Sp. | AUC | Pr. | Rec. | Sp. |
| Spinal Drain | 0.963 | 0.172 | 1.0 | 0.954 | 0.984 | 0.132 | 1.0 | 0.968 |
| IntraOp Somatosensory Evoked Potential | 0.994 | 0.4 | 1.0 | 0.994 | 1.0 | 1.0 | 1.0 | 1.0 |
| Aorta Device Inserted | 0.977 | 0.812 | 0.915 | 0.967 | 0.979 | 0.772 | 0.95 | 0.961 |
| Aortic Valve or Aortic Valve Composite Graft Implanted | 1.0 | 0.379 | 0.917 | 0.964 | 0.947 | 0.571 | 0.897 | 0.957 |
| **Post-operative** | | | | | | | | |
| Blood Prod | 0.92 | 0.745 | 0.973 | 0.839 | 0.86 | 0.727 | 0.816 | 0.856 |
| Extubated In OR | 0.9 | 0.267 | 1.0 | 0.753 | 0.83 | 0.234 | 1.0 | 0.0 |
| Postop Intubation/Reintubation During Hospital Stay | 0.992 | 0.643 | 1.0 | 0.979 | 0.828 | 0.231 | 0.818 | 0.942 |
| ICU Visit | 0.916 | 0.998 | 0.973 | 0.5 | 0.538 | 1.0 | 0.0 | 1.0 |
| Readmission to ICU | 0.948 | 0.231 | 1.0 | 0.918 | 0.842 | 0.075 | 0.75 | 0.829 |
| Postop Echo | 0.872 | 0.639 | 0.885 | 0.78 | 0.876 | 0.545 | 0.783 | 0.849 |
| Postop EF Done | 0.909 | 0.587 | 0.86 | 0.841 | 0.764 | 0.64 | 0.8 | 0.78 |
| Post-Op-Surgical Site Infection | 0.982 | 0.091 | 1.0 | 0.982 | 1.0 | 0.0 | 1.0 | 0.882 |
| Post-Op-Deep Sternal | 0.993 | 1.0 | 0.0 | 1.0 | 0.977 | 0.041 | 1.0 | 0.953 |
| In Hospital Post-Op Events | 0.878 | 0.83 | 0.938 | 0.802 | 0.869 | 0.707 | 0.813 | 0.851 |
| Post-Op-ReOp Bleeding/Tampanade | 0.991 | 0.268 | 1.0 | 0.942 | 0.977 | 0.368 | 1.0 | 0.863 |
| Post-Op-Unplanned Coronary Artery Intervention | 0.97 | 0.125 | 1.0 | 0.94 | 0.886 | 0.0 | 0.0 | 0.97 |
| Post-Op-Aortic Intervention | 1.0 | 1.0 | 1.0 | 1.0 | 1.0 | 0.0 | 1.0 | 0.977 |
| Post-Op-ReOp Other Cardiac Reasons | 0.948 | 0.04 | 1.0 | 0.946 | 0.982 | 0.107 | 1.0 | 0.971 |
| Post-Op-Return To OR For Other Non-cardiac Reason | 0.973 | 0.146 | 0.857 | 0.919 | 0.955 | 0.229 | 0.889 | 0.962 |
| Post-Op-Open Chest With Planned Delayed Sternal Closure | 0.99 | 0.122 | 1.0 | 0.907 | 0.988 | 0.256 | 0.917 | 0.969 |
| Post-Op-Sepsis | 0.98 | 0.152 | 1.0 | 0.945 | 0.99 | 0.097 | 1.0 | 0.97 |
| Post-Op-Neuro-Stroke Perm | 0.959 | 0.095 | 1.0 | 0.919 | 0.974 | 0.227 | 1.0 | 0.941 |
| Post-Op-Recurrent Laryngeal Nerve Injury | 0.837 | 0.028 | 0.5 | 0.755 | 0.997 | 0.027 | 1.0 | 0.954 |
| Post-Op-Pulm-Vent Prolonged | 0.965 | 0.541 | 0.971 | 0.898 | 0.943 | 0.514 | 0.902 | 0.953 |
| Post-Op-Pulm-Pneumonia | 0.985 | 0.379 | 1.0 | 0.953 | 0.979 | 0.289 | 0.929 | 0.961 |
| Post-Op-Pulmonary Thromboembolism | 1.0 | 0.0 | 1.0 | 0.997 | 1.0 | 1.0 | 1.0 | 1.0 |
| Post-Op-Pleural Effusion Requiring Drainage | 0.907 | 0.295 | 0.867 | 0.913 | 0.88 | 0.214 | 0.692 | 0.941 |
| Post-Op-Renal-Renal Failure | 0.991 | 0.333 | 1.0 | 0.977 | 0.938 | 0.234 | 0.846 | 0.956 |
| Post-Op-Vasc-Acute Limb Ischemia | 1.0 | 0.0 | 1.0 | 0.998 | 0.992 | 0.062 | 1.0 | 0.959 |
| Post-Op-Deep Venous Thrombosis | 1.0 | 0.24 | 1.0 | 0.946 | 0.81 | 0.023 | 0.333 | 0.931 |
| Post-Op-New Dysrhythmia Requiring Insertion of a Permanent Device | 1.0 | 0.556 | 1.0 | 0.989 | 0.926 | 0.275 | 0.731 | 0.913 |
| Post-Op-Other-Cardiac Arrest | 0.991 | 0.065 | 1.0 | 0.938 | 0.984 | 0.219 | 1.0 | 0.971 |
| Post-Op-Other-Anticoagulant Bleeding Event | 0.94 | 0.071 | 1.0 | 0.957 | 1.0 | 0.0 | 1.0 | 0.957 |
| Post-Op-Other-Pericardiocentesis | 0.979 | 0.091 | 1.0 | 0.97 | 1.0 | 1.0 | 1.0 | 1.0 |

Table S4: Performance of AI-derived variables after Confidence Intervals

| | MGB | | | | HHC | | | |
|---|---|---|---|---|---|---|---|---|
| Variable | AUC | Pr. | Rec. | Sp. | AUC | Pr. | Rec. | Sp. |
| Post-Op-Other-Gastro-Intestinal Event | 0.964 | 0.206 | 1.0 | 0.92 | 0.918 | 0.263 | 0.769 | 0.962 |
| Post-Op-Other-A Fib | 0.939 | 0.785 | 0.903 | 0.895 | 0.917 | 0.718 | 0.839 | 0.901 |

AUC, Pr., Rec. and Sp.: ROC Area Under Curve, Precision, Recall and Specificity of AI-derived variables after applying confidence intervals. Metrics for each variable are calculated only on the subset of the test set for which the pipeline chooses to complete, excluding records deferred to human review.

##### 3.2 Pipeline Performance with Performance Guarantees

Table S5: Completion results on AI-derived and Parent-Child variables.

| | MGB | | HHC | |
|---|---|---|---|---|
| Variable | ACC | Compl. (%) | ACC | Compl. (%) |
| **Pre-operative** | | | | |
| RF-Diabetes | 0.964 | 98.6% | 0.964 | 97.1% |
| RF-Diabetes-Control | 0.99 | 70.5% | 0.983 | 63.7% |
| RF-Renal Fail-Dialysis | 1.0 | 100.0% | | |
| RF- Endocarditis | | | 0.973 | 94.4% |
| RF-Infect Endocard Type | 1.0 | 89.0% | 0.999 | 88.7% |
| RF-Infect Endocard Culture | 0.998 | 87.9% | 0.998 | 88.7% |
| RF-Arterial Blood Gas | 1.0 | 100.0% | | |
| RF-Illicit Drug Use within One Year | 1.0 | 100.0% | | |
| RF-Intravenous Drug Use within One Year | 1.0 | 100.0% | 1.0 | 29.0% |
| RF-Drug use within 30 days of procedure | 1.0 | 100.0% | 0.997 | 29.0% |
| RF-Liver Cirrhosis | | | 0.972 | 84.2% |
| RF-Liver Disease - Child Pugh Class | | | 1.0 | 81.3% |
| Mediastinal Radiation | 0.983 | 72.7% | 0.964 | 100.0% |
| Unresponsive State | 1.0 | 100.0% | | |
| Prior CVA | | | 0.963 | 85.7% |
| Prior CVA-When | 0.997 | 70.7% | 0.991 | 75.2% |
| CVD TIA | | | 0.998 | 53.2% |
| CVD Carotid Stenosis | 0.99 | 52.4% | 0.989 | 53.2% |
| CVD Carotid Stenosis - Right | 0.997 | 52.4% | 0.998 | 53.2% |
| CVD Carotid Stenosis - Left | 0.99 | 52.4% | 0.995 | 53.2% |
| HIT Antibodies | | | 1.0 | 100.0% |
| Five Meter Walk Time 1 | 1.0 | 77.2% | 0.991 | 21.1% |
| Five Meter Walk Time 2 | 1.0 | 77.2% | 0.991 | 21.1% |
| Five Meter Walk Time 3 | 1.0 | 77.2% | 0.991 | 21.1% |
| Prev CAB | | | 0.961 | 84.0% |
| Prev Valve | | | 0.964 | 92.2% |
| Prev Valve Procedure 1 | 0.998 | 83.4% | 0.994 | 83.3% |
| Prev Valve Procedure 2 | 0.998 | 83.4% | 0.994 | 83.3% |
| Prev Valve Procedure 3 | 1.0 | 83.4% | 1.0 | 83.3% |
| Prev Valve Procedure 4 | 1.0 | 83.4% | 1.0 | 83.3% |
| Prev Valve Procedure 5 | 1.0 | 83.4% | 1.0 | 83.3% |
| Previous PCI-Within This Episode of Care | 0.987 | 98.0% | 0.981 | 87.6% |
| Previous PCI-Indication For Surgery | 1.0 | 96.2% | 0.998 | 86.1% |
| Previous PCI-Stent | 0.989 | 83.9% | 0.988 | 83.1% |
| Previous PCI-Interval | 0.996 | 96.4% | 0.998 | 92.9% |
| Previous Other Cardiac Intervention 1 | 0.978 | 66.5% | 0.984 | 61.7% |
| Previous Other Cardiac Intervention 2 | 0.978 | 66.5% | 0.984 | 61.7% |
| Previous Other Cardiac Intervention 3 | 1.0 | 66.5% | 1.0 | 61.7% |
| Previous Other Cardiac Intervention 4 | 1.0 | 66.5% | 1.0 | 61.7% |
| Previous Other Cardiac Intervention 5 | 1.0 | 66.5% | 1.0 | 61.7% |
| Previous Other Cardiac Intervention 6 | 1.0 | 66.5% | 1.0 | 61.7% |
| MI-When | 0.989 | 50.3% | 0.978 | 62.9% |
| Heart Failure Timing | 0.996 | 41.4% | 0.989 | 36.6% |
| Heart Failure Type | 0.991 | 41.4% | | |
| Cardiogenic Shock | | | 0.961 | 47.8% |
| Resuscitation | 1.0 | 100.0% | | |
| Cardiac Arrhythmia - Permanently Paced Rhythm | 0.984 | 68.0% | 0.974 | 55.8% |
| Cardiac Arrhythmia - Sick Sinus Syndrome | 0.99 | 56.8% | 0.973 | 49.5% |
| Cardiac Arrhythmia - AFlutter | 0.964 | 91.3% | | |
| Cardiac Arrhythmia - Second Degree Heart Block | 1.0 | 47.4% | 0.996 | 52.9% |

Table S5: Completion results on AI-derived and Parent-Child variables.

| | MGB | | HHC | |
|---|---|---|---|---|
| Variable | ACC | Compl. (%) | ACC | Compl. (%) |
| Cardiac Arrhythmia - Third Degree Heart Block | 0.982 | 61.5% | 0.97 | 57.9% |
| Atrial Fibrillation - Type | 1.0 | 69.4% | 0.998 | 59.5% |
| Patient in A-fib at OR Entry | 0.997 | 69.4% | 0.998 | 59.5% |
| Meds-Other Antianginal Medication Therapy For More Than 2 Weeks Prior To Surgery | 0.971 | 57.0% | 0.997 | 28.3% |
| Meds-ADP Inhibitors Within Five Days | 0.987 | 83.7% | | |
| Meds-ADP Inhibitors Discontinuation | 0.996 | 82.5% | | |
| Meds-Aspirin One-Time Dose | 0.978 | 48.8% | | |
| Meds-Glycoprotein IIb/IIIa Inhibitor Within 24 Hours | 0.993 | 100.0% | | |
| Meds-Anticoagulants-Medication Name | 0.988 | 60.0% | 0.983 | 39.4% |
| Meds-Warfarin (Coumadin) Discontinuation | 1.0 | 44.5% | 1.0 | 10.2% |
| Meds-DOAC Within 5 Days | 1.0 | 15.6% | | |
| Meds-DOAC Discontinuation | 1.0 | 15.6% | | |
| Meds-Thrombolytics Within 24 Hours | 1.0 | 100.0% | | |
| Meds-Inotropes Within 48 Hours | | | 0.965 | 79.3% |
| LAD Distribution Stenosis Percentage | 0.996 | 49.7% | | |
| LAD distribution stenosis - current revascularization - Location Known | 0.989 | 49.7% | | |
| LAD distribution stenosis - current revascularization | 0.989 | 49.7% | | |
| Ramus Stenosis Percentage | 0.996 | 49.7% | 1.0 | 29.6% |
| Ramus stenosis - current revascularization - Location Known | 0.996 | 49.7% | 0.993 | 29.6% |
| Ramus stenosis - current revascularization | 0.996 | 49.7% | 0.993 | 29.6% |
| Circumflex Distribution Stenosis Percentage | | | 0.973 | 38.8% |
| Circumflex distribution stenosis - current revascularization - Location Known | 0.996 | 50.6% | | |
| Circumflex distribution stenosis - current revascularization | 0.996 | 50.6% | | |
| RCA Distribution Stenosis Percentage | | | 0.965 | 33.7% |
| RCA distribution stenosis - current revascularization - Location Known | 0.996 | 49.9% | | |
| RCA distribution stenosis - current revascularization | 0.996 | 49.9% | | |
| Aortic Valve Stenosis Degree | 0.997 | 69.4% | 0.975 | 46.5% |
| VD-Aortic Valve Area | 0.995 | 72.0% | 0.981 | 60.6% |
| VD-Aortic Gradient-Mean | 0.977 | 72.0% | 0.982 | 60.6% |
| VD - Aortic Jet Velocity (Vmax) | 0.997 | 72.0% | 0.984 | 60.6% |
| VD-Aortic Valve Disease Primary Etiology | 0.967 | 33.1% | | |
| Mitral Valve Regurgitation | 0.986 | 100.0% | | |
| Mitral Valve Stenosis Degree | 1.0 | 54.8% | 0.999 | 65.4% |
| Mitral - Valve Area | 1.0 | 78.5% | 0.996 | 78.6% |
| Mitral - Mean Gradient | 1.0 | 78.5% | 0.996 | 78.6% |
| VD-Mitral Valve Disease | 0.975 | 14.3% | | |
| VD-Mitral Valve Disease - Carpentier Classification - Class I - Type | 1.0 | 14.3% | 0.98 | 9.5% |
| VD-Mitral Valve Disease - Carpentier Classification - Class II - Type | 1.0 | 14.3% | 1.0 | 9.5% |
| VD-Mitral Valve Disease - Carpentier Classification - Class II - Myomatous | 1.0 | 14.3% | 1.0 | 9.5% |
| VD-Mitral Valve Disease - Carpentier Classification - Class III A - Type | 1.0 | 14.3% | 0.99 | 9.5% |
| VD-Mitral Valve Disease - Carpentier Classification - Class III B - Type | 1.0 | 14.3% | 1.0 | 9.5% |
| VD-Mitral Valve Disease - Mixed Lesion - Type | 1.0 | 14.3% | 1.0 | 9.5% |
| Tricuspid Valve Regurgitation | 0.986 | 100.0% | | |
| VD-Tricuspid Annulus Size (Diameter) | 1.0 | 11.6% | 1.0 | 14.7% |
| VD-Pulmonic Valve Stenosis | 0.98 | 82.6% | 1.0 | 100.0% |
| Pulmonic Valve Stenosis Degree | 1.0 | 80.7% | 1.0 | 100.0% |
| VD-Pulmonic Hemodynamic Data Available | 1.0 | 80.7% | 1.0 | 100.0% |
| VD-Pulmonic Gradient-Highest Mean | 1.0 | 80.7% | 1.0 | 100.0% |
| VD-Pulmonic Valve Disease Etiology | 0.979 | 8.5% | | |
| **Intra-operative** | | | | |
| Robot Used | 1.0 | 100.0% | | |

Table S5: Completion results on AI-derived and Parent-Child variables.

| | MGB | | HHC | |
|---|---|---|---|---|
| Variable | ACC | Compl. (%) | ACC | Compl. (%) |
| CAB | 0.971 | 100.0% | 0.964 | 100.0% |
| Surgeon Input for Aortic Surgery Data Abstraction | 0.963 | 94.0% | | |
| Valve Prosthesis Explant | 0.98 | 100.0% | | |
| VS-Aortic Valve | 0.96 | 45.2% | | |
| VS-Mitral Valve | 0.98 | 100.0% | 0.973 | 100.0% |
| VS-Tricuspid Valve | 0.982 | 100.0% | | |
| VS-Pulmonic Valve | 0.995 | 100.0% | 1.0 | 100.0% |
| Mechanical Assist Device / Ventricular Assist Device | 0.975 | 85.7% | | |
| Atrial Fibrillation Procedure Performed | 0.966 | 100.0% | | |
| Surgeon Input for Other Cardiac Afib Data Abstraction | 0.992 | 86.6% | 0.993 | 72.1% |
| General Anesthesia | 0.989 | 100.0% | | |
| Intubation | 0.96 | 100.0% | | |
| Appropriate Antibiotic Administration Timing | 0.975 | 57.0% | | |
| Temperature Measured | 0.975 | 100.0% | | |
| Lowest Hematocrit during CPB | 1.0 | 79.9% | 0.997 | 84.1% |
| Circulatory Arrest Time Without Cerebral Perfusion | 0.995 | 79.9% | 0.997 | 84.1% |
| Cerebral Perfusion Time | 0.998 | 90.2% | 0.997 | 88.0% |
| Cerebral Perfusion Type | 0.998 | 90.2% | 0.997 | 88.0% |
| Total Circulatory Arrest Time | 0.995 | 79.9% | 0.997 | 84.1% |
| Cooling Time prior to Circ Arrest | 1.0 | 79.9% | 0.998 | 84.1% |
| Intraop Blood Products - RBC Units | 0.966 | 21.2% | | |
| Intraop Blood Products - Platelet Dose Pack | 0.983 | 21.2% | | |
| Intraop Blood Products - FFP/Plasma Units | 0.966 | 21.2% | | |
| Intraop Blood Products - Cryo Units | 0.966 | 21.2% | | |
| Aortic Gradient - Post Repair Mean | 1.0 | 9.0% | | |
| Mitral Gradient - Post Repair Mean | 1.0 | 9.0% | | |
| Tricuspid Gradient - Post Repair Mean | 1.0 | 9.0% | 0.978 | 4.4% |
| Ejection Fraction Post Procedure | 0.967 | 54.1% | | |
| Internal Mammary Artery Used | 0.967 | 86.8% | 0.962 | 99.3% |
| Left IMA Used | 0.964 | 100.0% | | |
| Distal Anastomoses with Arterial Conduit(s) | 0.964 | 66.0% | | |
| Total Number of Distal Anastomoses with Arterial Conduits | 0.994 | 60.9% | | |
| Distal Anastomoses with Radial Artery Conduit(s) Used | 0.987 | 100.0% | | |
| Radial Dist Anast # | 1.0 | 97.6% | 0.989 | 63.0% |
| Total Number of Distal Anastomoses with Venous Conduits | 0.969 | 64.2% | 0.963 | 55.1% |
| Proximal Technique | 0.961 | 59.9% | 0.964 | 50.6% |
| CAB Proximal Site 01 | 0.97 | 59.9% | | |
| CAB Conduit 01 | 0.976 | 59.9% | | |
| CAB Distal Position 01 | 0.964 | 66.2% | 0.963 | 54.3% |
| CAB Endarterectomy 01 | 0.975 | 71.4% | 0.971 | 84.6% |
| CAB 02 | 0.966 | 100.0% | | |
| CAB Proximal Site 02 | 0.984 | 66.9% | 0.964 | 58.5% |
| CAB Distal Site 02 | | | 0.964 | 58.5% |
| CAB Conduit 02 | 0.965 | 66.9% | 0.964 | 58.5% |
| CAB Endarterectomy 02 | 1.0 | 66.9% | 1.0 | 58.5% |
| CAB Proximal Site 03 | 0.99 | 72.2% | 0.97 | 67.1% |
| CAB Distal Site 03 | 0.977 | 72.2% | 0.97 | 67.1% |
| CAB Conduit 03 | 0.98 | 72.2% | 0.97 | 67.1% |
| CAB Endarterectomy 03 | 0.996 | 96.2% | 0.999 | 88.0% |
| CAB 04 | 0.978 | 96.9% | | |
| CAB Proximal Site 04 | 0.998 | 82.3% | 0.984 | 79.0% |
| CAB Distal Site 04 | 0.991 | 82.3% | 0.984 | 79.0% |
| CAB Conduit 04 | 0.993 | 82.3% | 0.984 | 79.0% |
| CAB Distal Position 04 | 0.989 | 85.5% | 0.985 | 79.0% |

Table S5: Completion results on AI-derived and Parent-Child variables.

| | MGB | | HHC | |
|---|---|---|---|---|
| Variable | ACC | Compl. (%) | ACC | Compl. (%) |
| CAB Endarterectomy 04 | 1.0 | 82.3% | 1.0 | 79.0% |
| CAB 05 | | | 0.962 | 86.7% |
| CAB Proximal Site 05 | 1.0 | 85.4% | 0.998 | 82.6% |
| CAB Distal Site 05 | 0.998 | 85.4% | 0.998 | 82.6% |
| CAB Conduit 05 | 0.998 | 85.4% | 0.998 | 82.6% |
| CAB Distal Position 05 | 0.998 | 85.4% | 0.999 | 82.6% |
| CAB Endarterectomy 05 | 1.0 | 85.4% | 1.0 | 82.6% |
| CAB 06 | 0.998 | 85.4% | 1.0 | 82.6% |
| CAB Proximal Site 06 | 1.0 | 85.4% | 1.0 | 82.6% |
| CAB Distal Site 06 | 0.998 | 85.4% | 1.0 | 82.6% |
| CAB Conduit 06 | 0.998 | 85.4% | 1.0 | 82.6% |
| CAB Distal Position 06 | 0.998 | 85.4% | 1.0 | 82.6% |
| CAB 07 | 0.998 | 85.4% | 1.0 | 82.6% |
| CAB Proximal Site 07 | 1.0 | 85.4% | 1.0 | 82.6% |
| CAB Distal Site 07 | 0.998 | 85.4% | 1.0 | 82.6% |
| CAB Conduit 07 | 0.998 | 85.4% | 1.0 | 82.6% |
| CAB 08 | 1.0 | 85.4% | 1.0 | 82.6% |
| CAB Proximal Site 08 | 1.0 | 85.4% | 1.0 | 82.6% |
| CAB Distal Site 08 | 1.0 | 85.4% | 1.0 | 82.6% |
| CAB Conduit 08 | 1.0 | 85.4% | 1.0 | 82.6% |
| CAB Distal Position 08 | 1.0 | 85.4% | 1.0 | 82.6% |
| First Valve Prosthesis Explant Position | 1.0 | 93.7% | 0.999 | 66.3% |
| First Valve Explant Type | 0.992 | 93.7% | 0.999 | 66.3% |
| First Valve Explant Etiology | 0.994 | 93.7% | 0.999 | 66.3% |
| First Valve Explant Device Known | 0.975 | 99.8% | 0.961 | 92.4% |
| First Valve Explant Device | 0.998 | 94.2% | 0.998 | 86.3% |
| First Valve Explant Unique Device Identifier (UDI) | 1.0 | 94.2% | 1.0 | 86.3% |
| First Valve Explant Device Year Known | 1.0 | 94.2% | 0.998 | 86.3% |
| First Valve Explant Implant Year | 1.0 | 94.2% | 0.998 | 86.3% |
| Second Valve Prosthesis Explant | 1.0 | 100.0% | 0.979 | 98.6% |
| Second Valve Prosthesis Explant Position | 1.0 | 99.6% | 1.0 | 96.5% |
| Second Valve Explant Type | 1.0 | 99.6% | 1.0 | 96.5% |
| Second Valve Explant Etiology | 1.0 | 99.6% | 1.0 | 96.5% |
| Second Valve Explant Device Known | 1.0 | 99.6% | 1.0 | 96.5% |
| Second Valve Explant Device | 1.0 | 99.6% | 1.0 | 96.5% |
| Third Valve Prosthesis Explant | 1.0 | 99.6% | 1.0 | 96.5% |
| IABP | 0.962 | 94.9% | | |
| IABP-When Inserted | 0.988 | 92.0% | 0.999 | 64.7% |
| MCAD - ECMO | 1.0 | 82.1% | 1.0 | 33.4% |
| ECMO Mode | 1.0 | 82.1% | 1.0 | 33.4% |
| ECMO Initiated | 1.0 | 82.1% | 1.0 | 33.4% |
| Temporary Assist Device Used | 0.967 | 100.0% | 0.963 | 90.9% |
| Temporary Assist Device Used - Position | 1.0 | 96.4% | 1.0 | 87.4% |
| Temporary Assist Type | 1.0 | 96.4% | 1.0 | 87.4% |
| Temporary Assist Device When Inserted | 1.0 | 96.4% | 1.0 | 87.4% |
| VAD-Patient Admitted With VAD | 0.984 | 100.0% | 0.999 | 100.0% |
| Previous VAD Insertion Date | 1.0 | 96.2% | 1.0 | 100.0% |
| Previous VAD Device Model Number | 1.0 | 96.2% | 1.0 | 100.0% |
| Previous VAD Explanted During This Admission | 0.987 | 100.0% | 1.0 | 100.0% |
| Ventricular Assist Device Implanted During This Hospitalization | 0.98 | 100.0% | 0.971 | 100.0% |
| VAD-Implant Timing | 1.0 | 96.7% | 0.999 | 96.5% |
| VAD Implant Indication | 0.998 | 96.7% | 0.999 | 96.5% |
| VAD-Implant Type | 1.0 | 96.7% | 1.0 | 96.5% |
| VAD-Device | 0.998 | 96.7% | 0.999 | 96.5% |

Table S5: Completion results on AI-derived and Parent-Child variables.

| | MGB | | HHC | |
|---|---|---|---|---|
| Variable | ACC | Compl. (%) | ACC | Compl. (%) |
| VAD-Implant Date | 0.998 | 96.7% | 0.999 | 96.5% |
| VAD-Implant Unique Device Identifier (UDI) | 1.0 | 96.7% | 1.0 | 96.5% |
| VAD-Explant | 0.993 | 100.0% | 0.98 | 99.1% |
| VAD-Explant Reason | 1.0 | 98.0% | 1.0 | 97.1% |
| VAD-Explant Date | 1.0 | 98.0% | 1.0 | 97.1% |
| VAD-Implant #2 | 1.0 | 96.7% | 1.0 | 96.5% |
| VAD-Implant Timing #2 | 1.0 | 96.7% | 1.0 | 96.5% |
| VAD Implant Indication #2 | 1.0 | 96.7% | 1.0 | 96.5% |
| VAD-Implant Type #2 | 1.0 | 96.7% | 1.0 | 96.5% |
| VAD-Device #2 | 1.0 | 96.7% | 1.0 | 96.5% |
| VAD-Implant Date #2 | 1.0 | 96.7% | 1.0 | 96.5% |
| VAD-Explant #2 | 1.0 | 82.1% | 1.0 | 33.4% |
| VAD-Explant Reason #2 | 1.0 | 82.1% | 1.0 | 33.4% |
| VAD-Implant #3 | 1.0 | 96.7% | 1.0 | 96.5% |
| Other Card-Subaortic Stenosis Resection Type | 0.991 | 61.7% | 0.99 | 50.4% |
| Other Card-Pulmonary Thromboembolectomy | 0.984 | 81.6% | 1.0 | 59.0% |
| Other Card-LVA | 1.0 | 61.7% | 1.0 | 50.4% |
| Other Card-Arrhythmia Device Surgery | | | 0.99 | 50.4% |
| Other Card-Lead Insertion | 0.989 | 100.0% | 0.971 | 91.0% |
| Other Card-Arrhythmia Correction Surgery-Lead Extraction | 0.997 | 61.7% | 1.0 | 50.4% |
| Other Card-Tumor | | | 0.99 | 50.4% |
| Other Card-Card Tx | 0.993 | 100.0% | 0.973 | 99.5% |
| Other Card-Acquired VSD Repair | 1.0 | 61.7% | 1.0 | 50.4% |
| Other Card - ASD Repair | 1.0 | 61.7% | 1.0 | 50.4% |
| Other Card - ASD Repair Type | 1.0 | 61.7% | 1.0 | 50.4% |
| Other Card - PFO Repair | 1.0 | 61.7% | 0.998 | 50.4% |
| Other Card-Left Atrial Appendage Obliteration | 0.99 | 86.6% | 0.993 | 72.1% |
| Other Card- Epicardial Occlusion Device UDI | 1.0 | 86.6% | 1.0 | 72.1% |
| Other Card- Left Atrial Appendage Amputation | 1.0 | 86.6% | 1.0 | 72.1% |
| AFib Lesion Location | 0.992 | 86.6% | 0.993 | 72.1% |
| AFib Lesion - Method | 1.0 | 86.6% | 1.0 | 72.1% |
| Atrial Fibrillation - Lesion Creation Radiofrequency - Bipolar | 0.992 | 88.2% | 0.994 | 78.6% |
| Lesions Documented | 0.978 | 99.8% | | |
| Atrial Afibrillation - Lesion - Left Atrial | 1.0 | 88.8% | 0.993 | 84.7% |
| Atrial Afibrillation - Left Atrial Lesion - Method | 1.0 | 88.8% | 0.993 | 84.7% |
| Atrial Afibrillation - Right Atrial Lesion | 1.0 | 88.8% | 1.0 | 84.7% |
| Atrial Afibrillation - Right Atrial Lesion - Method | 1.0 | 88.8% | 1.0 | 84.7% |
| Family History Of Disease Of The Aorta | 0.988 | 78.5% | 0.993 | 73.8% |
| Patient's Genetic History | 0.988 | 78.5% | 0.993 | 73.8% |
| Prior Aortic Intervention | | | 0.964 | 89.8% |
| Prior Aortic Intervention - Previous Repair - Root (Zone 0 - A) | 1.0 | 84.3% | 1.0 | 85.2% |
| Prior Aortic Intervention - Repair Failure - Root (Zone 0 - A) | 1.0 | 84.3% | 1.0 | 85.2% |
| Prior Aortic Intervention - Disease Progression - Root (Zone 0 - A) | 1.0 | 84.3% | 1.0 | 85.2% |
| Prior Aortic Intervention - Previous Repair - Ascending (Zone 0 - B&C) | 0.962 | 100.0% | 0.968 | 95.9% |
| Prior Aortic Intervention - Previous Repair Type - Ascending (Zone 0 - B&C) | 1.0 | 95.3% | 0.999 | 91.7% |
| Prior Aortic Intervention - Repair Failure - Ascending (Zone 0 - B&C) | 1.0 | 95.3% | 1.0 | 91.7% |
| Prior Aortic Intervention - Disease Progression - Ascending (Zone 0 - B&C) | 1.0 | 95.3% | 0.999 | 91.7% |
| Prior Aortic Intervention - Previous Repair Type - Arch (Zones 1,2,3) | 1.0 | 85.2% | 1.0 | 94.8% |
| Prior Aortic Intervention - Repair Failure - Arch (Zones 1,2,3) | 1.0 | 85.2% | 1.0 | 94.8% |
| Prior Aortic Intervention - Disease Progression - Arch (Zones 1,2,3) | 1.0 | 85.2% | 1.0 | 94.8% |
| Prior Aortic Intervention - Previous Repair - Descending (Zones 4,5) | 0.993 | 100.0% | 0.962 | 93.8% |

Table S5: Completion results on AI-derived and Parent-Child variables.

| | MGB | | HHC | |
|---|---|---|---|---|
| Variable | ACC | Compl. (%) | ACC | Compl. (%) |
| Prior Aortic Intervention - Previous Repair Type - Descending (Zones 4,5) | 1.0 | 98.9% | 1.0 | 89.7% |
| Prior Aortic Intervention - Repair Failure - Descending (Zones 4,5) | 1.0 | 98.9% | 1.0 | 89.7% |
| Prior Aortic Intervention - Disease Progression - Descending (Zones 4,5) | 1.0 | 98.9% | 1.0 | 89.7% |
| Prior Aortic Intervention - Previous Repair - Suprarenal Abdominal (Zones 6,7) | 1.0 | 84.3% | 1.0 | 85.2% |
| Prior Aortic Intervention - Previous Repair Type - Suprarenal Abdominal (Zones 6,7) | 1.0 | 84.3% | 1.0 | 85.2% |
| Prior Aortic Intervention - Repair Failure - Suprarenal Abdominal (Zones 6,7) | 1.0 | 84.3% | 1.0 | 85.2% |
| Prior Aortic Intervention - Disease Progression - Suprarenal Abdominal (Zones 6,7) | 1.0 | 84.3% | 1.0 | 85.2% |
| Prior Aortic Intervention - Previous Repair - Infrarenal Abdominal (Zones 8,9,10,11) | 1.0 | 86.3% | 0.981 | 91.0% |
| Prior Aortic Intervention - Previous Repair Type - Infrarenal Abdominal (Zones 8,9,10,11) | 1.0 | 86.3% | 1.0 | 89.0% |
| Current Procedure with Endoleak involvement | 1.0 | 94.6% | 0.987 | 76.0% |
| Current Procedure with Aorta Infection | 1.0 | 94.6% | 0.993 | 78.2% |
| Aorta Infection Type | 1.0 | 94.6% | 1.0 | 77.5% |
| Current Procedure with Trauma | 1.0 | 78.5% | 1.0 | 73.8% |
| Aorta Presentation | 0.988 | 78.5% | 0.999 | 73.8% |
| Aorta Primary indication | 0.988 | 78.5% | 0.993 | 73.8% |
| Aneurysm - Etiology | 0.993 | 78.5% | 0.999 | 73.8% |
| Aneurysm - Type | 0.993 | 78.5% | 0.999 | 73.8% |
| Aneurysm - Location | 1.0 | 78.5% | 1.0 | 73.8% |
| Dissection - Timing | 1.0 | 78.5% | 1.0 | 73.8% |
| Dissection Onset Date Known | 1.0 | 78.5% | 1.0 | 73.8% |
| Dissection Onset Date | 1.0 | 78.5% | 1.0 | 73.8% |
| Dissection - Primary Tear Location | 1.0 | 78.5% | 1.0 | 73.8% |
| Proximal Dissection Extent Known | 1.0 | 78.5% | 1.0 | 73.8% |
| Most Proximal Dissection Location | 1.0 | 78.5% | 1.0 | 73.8% |
| Distal Dissection Extent Known | 1.0 | 78.5% | 1.0 | 73.8% |
| Distal Dissection Extension Location | 1.0 | 78.5% | 1.0 | 73.8% |
| Stanford Classification Known | 1.0 | 78.5% | 1.0 | 73.8% |
| Dissection - Malperfusion | 1.0 | 78.5% | 1.0 | 73.8% |
| Dissection - Malperfusion Type | 1.0 | 78.5% | 1.0 | 73.8% |
| Dissection - Lower Extremity Motor Function | 1.0 | 78.5% | 1.0 | 73.8% |
| Dissection - Lower Extremity Sensory Deficit | 1.0 | 78.5% | 1.0 | 73.8% |
| Dissection - Rupture | 1.0 | 78.5% | 1.0 | 73.8% |
| Dissection - Rupture Location | 1.0 | 78.5% | 1.0 | 73.8% |
| Aorta Primary Indication - Other | 1.0 | 78.5% | 0.997 | 73.8% |
| Root - Aortic-Annular Ectasia | 0.968 | 91.3% | 0.997 | 93.4% |
| Root - Asymmetric Root Dilation | 0.998 | 91.5% | 0.996 | 78.1% |
| Root - Asymmetric Root Dilation - Location | 1.0 | 91.3% | 1.0 | 78.1% |
| Root - Sinus Of Valsalva Aneurysm | | | 0.962 | 88.7% |
| Root - Sinus Of Valsalva Aneurysm - Multi Location | 1.0 | 85.5% | 1.0 | 85.6% |
| Arch Anomalies | 1.0 | 78.5% | 1.0 | 73.8% |
| Arch Anomalies Type | 1.0 | 78.5% | 1.0 | 73.8% |
| Patent Internal Mammary Artery Bypass Graft | 0.996 | 87.2% | 0.995 | 73.8% |
| Ascending Asymmetric Dilation | 0.99 | 88.8% | 0.996 | 73.8% |
| Ascending Proximal Coronary Bypass Grafts | 0.989 | 85.2% | 0.986 | 83.5% |
| Treated Zone with the Largest Diameter | 1.0 | 78.5% | 0.999 | 73.8% |
| Treated Zone with the Largest Diameter - Measurement | 1.0 | 78.5% | 0.997 | 73.8% |

Table S5: Completion results on AI-derived and Parent-Child variables.

| | MGB | | HHC | |
|---|---|---|---|---|
| Variable | ACC | Compl. (%) | ACC | Compl. (%) |
| Treated Zone with the Largest Diameter - Method Obtained | 1.0 | 78.5% | 1.0 | 73.8% |
| Proximal to Treated Zone(s) (Largest Diameter) Available | 0.967 | 86.8% | | |
| Proximal to Treated Zone(s) (Largest Diameter) Available - Location | 1.0 | 83.4% | 0.999 | 85.8% |
| Proximal to Treated Zone(s) (Largest Diameter) Available - Measurement | 1.0 | 83.4% | 0.999 | 85.8% |
| Proximal to Treated Zone(s) (Largest Diameter) - Method Obtained | 1.0 | 83.4% | 0.999 | 85.8% |
| Distal to Treated Zone(s) (Largest Diameter) Available - Location | 1.0 | 81.7% | 0.997 | 84.9% |
| Distal to Treated Zone(s) (Largest Diameter) Available - Measurement | 1.0 | 81.7% | 0.997 | 84.9% |
| Distal to Treated Zone(s) (Largest Diameter) Available - Method Obtained | 1.0 | 81.7% | 0.999 | 84.9% |
| VS-Aorta - Aortic Valve or Root Procedure Performed | 0.997 | 69.3% | 0.994 | 64.6% |
| VS-Aorta - Aortic Valve Procedure Performed | 0.997 | 69.3% | 1.0 | 64.6% |
| VS-Aorta - Aortic Surgical Valve Replacement | 1.0 | 69.3% | 0.994 | 64.6% |
| VS-Aorta - Aortic Surgical Valve Replacement - Device Type | 1.0 | 69.3% | 1.0 | 64.6% |
| VS-Aorta - Aortic Valve Procedure Repair Type | 0.997 | 69.3% | 1.0 | 64.6% |
| VS-Aorta - Replacement of non-coronary sinus (Modified Wheat/Modified Yacoub) | 1.0 | 69.3% | 1.0 | 64.6% |
| VS-Aortic Root Procedure | 0.995 | 69.3% | 0.995 | 64.6% |
| VS-Aortic Root Replacement With Coronary Ostial Reimplantation | 0.997 | 69.3% | 0.997 | 64.6% |
| VS-Aortic Root Replacement With Coronary Ostial Reimplantation - Type | 1.0 | 69.3% | 0.998 | 64.6% |
| VS-Aortic Root Procedure With Coronary Ostial Reimplantation (Bentall) - Type | 1.0 | 69.3% | 1.0 | 64.6% |
| VS-Aortic Root Procedure With Coronary Ostial Reimplantation - Bioprosthetic Type | 1.0 | 69.3% | 0.998 | 64.6% |
| VS-Aortic Valve Sparing Root Operation | 0.997 | 69.3% | 0.998 | 64.6% |
| Coronary Reimplantation | 1.0 | 69.3% | 0.998 | 64.6% |
| VS-Aortic Valve Major Root Reconstruction/Debridement without coronary ostial reimplantation | 1.0 | 69.3% | 0.998 | 64.6% |
| Surgical Ascending/Arch Procedure - Proximal Location | 1.0 | 79.9% | 0.994 | 84.4% |
| Open Arch Procedure - Distal Technique | 0.996 | 81.9% | 0.994 | 84.4% |
| Open Arch Procedure - Distal Site | 0.993 | 79.9% | 0.995 | 84.4% |
| Open Arch Procedure - Distal Extension | 0.993 | 79.9% | 0.994 | 84.4% |
| Open Arch Procedure - Arch Branch Reimplantation | 0.985 | 99.1% | 0.98 | 90.7% |
| Arch Branch Location | 1.0 | 96.7% | 0.999 | 88.3% |
| Surgical Descending Thoracic Aorta or Thoracoabdominal Procedure | 0.996 | 94.4% | 0.981 | 86.4% |
| Open Surgical Descending - Proximal Location | 0.996 | 94.4% | 1.0 | 84.5% |
| Intercostal Reimplantation | 0.996 | 94.4% | 1.0 | 84.5% |
| Distal Location | 0.996 | 94.4% | 1.0 | 84.5% |
| Visceral Vessel Intervention | 1.0 | 94.4% | 1.0 | 84.5% |
| Visceral Vessel Intervention - Celiac | 1.0 | 94.4% | 1.0 | 84.5% |
| Visceral Vessel Intervention - Superior Mesenteric | 1.0 | 94.4% | 1.0 | 84.5% |
| Visceral Vessel Intervention - Right Renal | 1.0 | 94.4% | 1.0 | 84.5% |
| Visceral Vessel Intervention - Left Renal | 1.0 | 94.4% | 1.0 | 84.5% |
| Endovascular Procedures | 0.998 | 94.6% | 1.0 | 73.8% |
| Endovascular Procedures - Access | 1.0 | 94.0% | 1.0 | 73.8% |
| Endovascular Procedures - Percutaneous Access | 1.0 | 94.0% | 1.0 | 73.8% |
| Endovascular Procedures - Proximal Landing Zone | 1.0 | 94.0% | 1.0 | 73.8% |
| Endovascular Procedures - Distal Landing Zone | 1.0 | 94.0% | 1.0 | 73.8% |
| Arch Vessel Management - Innominate | 1.0 | 94.0% | 1.0 | 73.8% |
| Arch Vessel Management - Left Carotid | 1.0 | 94.0% | 1.0 | 73.8% |
| Arch Vessel Management - Left Subclavian | 1.0 | 94.0% | 1.0 | 73.8% |
| Visceral Vessel Management - Celiac | 1.0 | 94.0% | 1.0 | 73.8% |
| Visceral Vessel Management - Superior Mesenteric | 1.0 | 94.0% | 1.0 | 73.8% |

Table S5: Completion results on AI-derived and Parent-Child variables.

| | MGB | | HHC | |
|---|---|---|---|---|
| Variable | ACC | Compl. (%) | ACC | Compl. (%) |
| Visceral Vessel Management - Right Renal | 1.0 | 94.0% | 1.0 | 73.8% |
| Visceral Vessel Management - Left Renal | 1.0 | 94.0% | 1.0 | 73.8% |
| Visceral Vessel Management - Internal Iliac Preserved | 1.0 | 94.0% | 1.0 | 73.8% |
| Planned Staged Hybrid | 0.998 | 100.0% | 0.984 | 100.0% |
| Dissection Proximal Entry Tear Covered | 1.0 | 94.0% | 1.0 | 73.8% |
| Conversion To Open | 1.0 | 94.0% | 1.0 | 73.8% |
| Intraop Dissection Extension | 1.0 | 94.0% | 1.0 | 73.8% |
| Spinal Drain | | | 0.968 | 99.2% |
| IntraOp Motor Evoked Potential | 0.987 | 94.4% | 1.0 | 73.8% |
| IntraOp Motor Evoked Potential - Documented MEP Abnormality | 1.0 | 92.9% | 1.0 | 73.8% |
| IntraOp Somatosensory Evoked Potential | 0.994 | 94.6% | 1.0 | 73.8% |
| IntraOp Somatosensory Evoked Potential - Documented SEP Abnormality | 1.0 | 93.7% | 1.0 | 73.8% |
| IntraOp EEG | 0.983 | 94.6% | 1.0 | 73.8% |
| IntraOp Intravascular Ultrasound (IVUS) | 1.0 | 78.5% | 1.0 | 73.8% |
| IntraOp Angiogram | 0.986 | 91.3% | 1.0 | 73.8% |
| IntraOp Angiogram - Volume Of Contrast | 1.0 | 90.1% | 1.0 | 73.8% |
| Endovascular Balloon Fenestration of the Dissection Flap | 1.0 | 78.5% | 0.993 | 73.8% |
| Aortic Valve or Aortic Valve Composite Graft Implanted - Model Number | 0.998 | 87.5% | 0.993 | 83.8% |
| Aortic Valve or Aortic Valve Composite Graft Implanted - Size | 0.998 | 87.5% | 0.993 | 83.8% |
| Aorta Device - Location #01 | 0.991 | 80.1% | 0.998 | 82.3% |
| Aorta Device - Implant Method #01 | 0.986 | 80.1% | 0.998 | 82.3% |
| Aorta Device - Outcome #01 | 0.986 | 80.1% | 0.996 | 82.3% |
| Aorta Device - Model Number #01 | 0.991 | 80.1% | 0.998 | 82.3% |
| Aorta Device - Unique Device Identifier #01 | 1.0 | 80.1% | 1.0 | 82.3% |
| Aorta Device - Location #02 | 0.986 | 80.1% | 0.996 | 82.3% |
| Aorta Device - Implant Method #02 | 1.0 | 80.1% | 1.0 | 82.3% |
| Aorta Device - Model Number #02 | 1.0 | 80.1% | 1.0 | 82.3% |
| Aorta Device - Unique Device Identifier #02 | 1.0 | 80.1% | 1.0 | 82.3% |
| Aorta Device - Location #03 | 1.0 | 80.1% | 1.0 | 82.3% |
| Aorta Device - Implant Method #03 | 1.0 | 80.1% | 1.0 | 82.3% |
| Aorta Device - Model Number #03 | 1.0 | 80.1% | 1.0 | 82.3% |
| Aorta Device - Location #04 | 1.0 | 80.1% | 1.0 | 82.3% |
| Aorta Device - Implant Method #04 | 1.0 | 80.1% | 1.0 | 82.3% |
| Aorta Device - Location #05 | 1.0 | 80.1% | 1.0 | 82.3% |
| Other Non Card-Caro Endart | 1.0 | 52.4% | 1.0 | 14.5% |
| Other Non Card-Other Vasc | 1.0 | 52.4% | 0.993 | 14.5% |
| Other Non Card-Other Thor | 1.0 | 52.4% | 1.0 | 14.5% |
| Other Non Card-Other | 1.0 | 52.4% | 1.0 | 14.5% |
| **Post-operative** | | | | |
| Blood Prod - RBC Units | 0.992 | 23.9% | | |
| Blood Prod - Fresh Frozen Plasma/Plasma Units | 0.992 | 23.9% | | |
| Blood Prod - Cryo Units | 0.992 | 23.9% | | |
| Blood Prod - Platelet Dose Pack | 1.0 | 23.9% | | |
| Postop Intubation/Reintubation During Hospital Stay | 0.98 | 44.7% | | |
| Additional Hours Ventilated | 1.0 | 42.1% | 0.996 | 47.6% |
| ICU Visit | 0.971 | 100.0% | | |
| Additional ICU Hours | 1.0 | 20.3% | 0.994 | 17.6% |
| Postop Echo Aortic Paravalvular Leak | 0.969 | 17.7% | | |
| Postop Echo Mitral Insufficiency | 0.98 | 17.7% | 0.966 | 17.4% |
| Postop Echo Mitral Paravalvular leak | 0.969 | 17.7% | | |
| Postop Echo Tricuspid Insufficiency | 0.969 | 17.7% | 0.966 | 17.4% |
| Postop Echo Pulmonic Insufficiency | 0.99 | 17.7% | 0.961 | 17.4% |

Table S5: Completion results on AI-derived and Parent-Child variables.

| | MGB | | HHC | |
|---|---|---|---|---|
| Variable | ACC | Compl. (%) | ACC | Compl. (%) |
| Post-Op-Surgical Site Infection | 0.982 | 100.0% | | |
| Post-Op-Superficial Sternal Wound | 0.996 | 98.0% | 1.0 | 23.2% |
| Post-Op-Deep Sternal | 0.998 | 99.8% | | |
| Post-Op-Deep Sternal - Diagnosis Date | 0.998 | 99.8% | 1.0 | 93.0% |
| Post-Op-Conduit Harvest (within 30 days or initial hospitalization) | 1.0 | 98.0% | 1.0 | 23.2% |
| Post-Op-Cannulation Site (within 30 days or initial hospitalization) | 1.0 | 98.0% | 1.0 | 23.2% |
| Deep Sternal Wound Infection Within 90 Days | 1.0 | 100.0% | | |
| Post-Op-ReOp Bleed Timing | 1.0 | 87.5% | 1.0 | 21.9% |
| Post-Op-ReOp for Valvular Dysfunction | 1.0 | 100.0% | 1.0 | 21.9% |
| Post-Op-Unplanned Coronary Artery Intervention | | | 0.968 | 60.7% |
| Post-Op-Unplanned Coronary Artery Intervention - Vessels | 1.0 | 19.7% | 0.998 | 58.9% |
| Post-Op-Unplanned Coronary Artery Intervention - Intervention Type | 1.0 | 19.7% | 1.0 | 58.9% |
| Post-Op-Aortic Intervention | 1.0 | 86.1% | 0.977 | 98.5% |
| Post-Op-Aortic Reintervention-Type | 1.0 | 86.1% | 1.0 | 96.3% |
| Post-Op-ReOp Other Cardiac Reasons | | | 0.971 | 84.1% |
| Post-Op-Return To OR For Other Non-cardiac Reason | | | 0.961 | 69.0% |
| Post-Op-Open Chest With Planned Delayed Sternal Closure | | | 0.968 | 99.7% |
| Post-Op-Sepsis | | | 0.97 | 90.1% |
| Post-Op-Neuro - Lower Extremity Paralysis >24 Hours | 1.0 | 19.7% | 1.0 | 21.9% |
| Post-Op-Neuro-Paresis >24 Hours | 1.0 | 19.7% | 1.0 | 21.9% |
| Pulm - Prolonged Ventilation - Tracheostomy Required after OR Exit | 1.0 | 44.8% | 1.0 | 68.6% |
| Post-Op-Pulm-Pneumonia | | | 0.96 | 80.0% |
| Post-Op-Pulmonary Thromboembolism | 0.974 | 62.4% | 1.0 | 21.9% |
| Post-Op-Pneumothorax Requiring Intervention | | | 0.987 | 21.9% |
| Post-Op-Renal-Renal Failure | 0.977 | 94.9% | | |
| Post-Op-Renal-Dialysis Req | 1.0 | 91.7% | 1.0 | 76.7% |
| Post-Op-Dialysis Required After Discharge | 1.0 | 91.7% | 1.0 | 76.7% |
| Post-Op-Vasc-Iliac/Fem Dissect | 1.0 | 19.7% | 1.0 | 21.9% |
| Post-Op-Vasc-Acute Limb Ischemia | 0.998 | 94.8% | | |
| Post-Op-Mechanical Assist Device Related Complication | 1.0 | 86.1% | 1.0 | 21.9% |
| Post-Op-New Dysrhythmia Requiring Insertion of a Permanent Device | 0.967 | 65.8% | | |
| Post-Op-Other-Cardiac Arrest | | | 0.972 | 85.4% |
| PostOp - Other - Aortic Complication | 0.978 | 65.8% | 1.0 | 21.9% |
| Post-Op-Other-Aortic Dissection | 1.0 | 65.8% | 1.0 | 21.9% |
| Post-Op-Other-Aortic Side Branch Malperfusion | 1.0 | 65.8% | 1.0 | 21.9% |
| Anticoagulant Bleeding Event - Type | 1.0 | 51.7% | 1.0 | 85.2% |
| Heparin Induced Thrombocytopenia (HIT) | 1.0 | 19.7% | 1.0 | 21.9% |
| Heparin Induced Thrombocytopenia Thrombosis | 1.0 | 19.7% | 1.0 | 21.9% |
| Post-Op-Other-Pericardiocentesis | 0.97 | 59.5% | 1.0 | 100.0% |
| Gastro-Intestinal Event - Type | 1.0 | 56.4% | 0.996 | 69.0% |

ACC: Completion accuracy. Evaluated only over the records that were completed.
Compl.: Percentage of records that were completed.

Accuracy and completion rate are calculated after using confidence intervals, thresholding and parent-child relationships. Only variables where we achieve a non-zero level of completion in MGB or HHC are shown.

##### 3.3 Deterministic Variables

Table S6: Statistics on Deterministic Variables

| Variable | Mode | MGB | HHC |
|---|---|---|---|
| **Continuous** | | | |
| Last A1C Level | | 6.0 (1.1) | 6.3 (1.5) |
| Patient Age | | 64.2 (12.4) | 66.3 (11.2) |
| Calculated BMI | | 28.3 (5.7) | 29.1 (6.2) |
| Last Creatinine Level | | 1.1 (0.7) | 1.2 (1.0) |
| Hematocrit | | 40.8 (5.2) | 39.2 (5.3) |
| Height (cm) | | 171.8 (10.1) | 171.3 (10.3) |
| Highest Intra-op Glucose | | 174.9 (38.4) | 174.6 (48.1) |
| INR | | 1.1 (0.3) | 1.1 (0.4) |
| Lowest Hematocrit | | 27.8 (4.6) | 23.9 (4.3) |
| Lowest Intra-op Hemoglobin | | 9.2 (1.7) | 8.1 (1.6) |
| RF-Carbon Dioxide Level | | 37.0 (4.9) | 34.4 (4.4) |
| Peak Postoperative Creatinine Level within 48 hours of OR Exit | | 1.2 (0.9) | 1.3 (1.0) |
| Platelet Count | | 220974.0 (65477.7) | 227762.7 (71470.1) |
| RF-Oxygen Level | | 96.5 (44.3) | 99.2 (33.8) |
| Peak Postoperative Creatinine Level prior to discharge | | 1.3 (1.0) | 1.5 (1.3) |
| Discharge Hematocrit | | 29.9 (4.3) | 31.6 (4.5) |
| Discharge Hemoglobin | | 9.8 (1.5) | 10.3 (1.7) |
| Postoperative Peak Glucose within 18-24 hours after OR Exit Time | | 150.5 (33.6) | 154.0 (38.7) |
| Hemoglobin | | 13.5 (1.9) | 13.1 (1.9) |
| Sodium | | 139.3 (2.8) | 138.4 (3.8) |
| Total Bilirubin | | 0.6 (0.6) | 0.6 (0.5) |
| WBC Count | | 7.4 (2.5) | 7.5 (3.7) |
| Weight (kg) | | 84.4 (18.9) | 85.7 (20.5) |
| **Categorical** | | | |
| Sex | Male | 5967 (70%) | 2095 (67%) |
| Meds-Beta Blockers Within 24 Hours | Yes | 5299 (62%) | 2310 (74%) |
| Race Documented | Yes | 8342 (98%) | 3046 (98%) |
| Race - Multi-Select | White | 0 (0%) | 2542 (82%) |
| RF-Tobacco Use | Never | 4154 (49%) | 1404 (45%) |
| **Other** | | | |
| Date of Birth | | | |
| Patient's Permanent Street Address | | | |
| Patient's Permanent City | | | |
| Patient First Name | | | |
| Patient Last Name | | | |
| Patient Middle Name | | | |
| Patient's Permanent Region | | | |
| ZIP Code | | | |

Mode: Most frequent value. Only reported for categorical variables.
MGB-HHC: Reporting mean (standard deviation) for continuous variables and count (percentage) of mode for categorical variables.

##### 3.4 Pipeline Performance Summary

Table S7: Completion Rate

| **Surgery** | **MGB** | | | | **HHC** | | | |
|---|---|---|---|---|---|---|---|---|
| | **All** | **Pre** | **Intra** | **Post** | **All** | **Pre** | **Intra** | **Post** |
| All | 49.5 % | 35.0 % | 61.5 % | 36.9 % | 43.2 % | 31.4 % | 52.6 % | 29.9 % |
| Aorta | 41.9 % | 41.4 % | 44.2 % | 36.7 % | 32.4 % | 34.4 % | 33.6 % | 26.4 % |
| CABG | 53.5 % | 41.0 % | 65.2 % | 41.5 % | 46.3 % | 33.6 % | 57.1 % | 33.1 % |
| Aortic V. | 47.7 % | 36.2 % | 57.4 % | 34.7 % | 41.8 % | 31.8 % | 49.5 % | 31.1 % |
| Mitral V. | 55.8 % | 39.8 % | 68.3 % | 37.2 % | 47.4 % | 31.6 % | 58.0 % | 28.8 % |
| Tricuspid V. | 51.3 % | 33.9 % | 64.3 % | 29.3 % | 42.5 % | 26.3 % | 54.5 % | 18.7 % |

Completion percentage for different surgery types, timings and study sites.
All: Variables that correspond to any admission time.
Pre: Variables that correspond to the pre-operative period.
Intra: Variables that correspond to the intra-operative period.
Post: Variables that correspond to the post-operative period.

Table S8: Completion Accuracy

| **Surgery** | **MGB** | | | | **HHC** | | | |
|---|---|---|---|---|---|---|---|---|
| | **All** | **Pre** | **Intra** | **Post** | **All** | **Pre** | **Intra** | **Post** |
| All | 99.4 % | 99.4 % | 99.4 % | 99.2 % | 99.4 % | 99.3 % | 99.5 % | 99.3 % |
| Aorta | 99.6 % | 99.5 % | 99.7 % | 99.7 % | 99.7 % | 99.6 % | 99.7 % | 99.8 % |
| CABG | 99.4 % | 99.2 % | 99.5 % | 99.2 % | 99.5 % | 99.4 % | 99.7 % | 99.2 % |
| Aortic Valve | 99.4 % | 99.3 % | 99.4 % | 99.3 % | 99.4 % | 99.3 % | 99.5 % | 99.2 % |
| Mitral Valve | 99.5 % | 99.4 % | 99.5 % | 99.4 % | 99.5 % | 99.3 % | 99.7 % | 99.3 % |
| Tricuspid Valve | 99.9 % | 99.8 % | 99.9 % | 99.9 % | 99.9 % | 99.8 % | 99.9 % | 99.9 % |

Completion accuracies for different surgery types, timings and study sites.
All: Variables that correspond to any admission time.
Pre: Variables that correspond to the pre-operative period.
Intra: Variables that correspond to the intra-operative period.
Post: Variables that correspond to the post-operative period.

##### 3.5 Quality Control Analysis

We manually reviewed the conflicts between the pipeline's predictions and the ground-truth of the STS tables for a subset of variables. The manual review is performed by a domain expert (data manager or physician) at each of the two hospital systems.

Table S9: Quality Control Results by Variable Type

| Variable Type | AUC | Manual Review (%) | Agreement Rate w/ model |
|---|---|---|---|
| Diabetes (MGH) | 98.5% | 28 (3.3%) | 17.9 % |
| CABG Procedure (MGH) | 93% | 11 (1.2%) | 9.1% |

Manual Review n (%): Number and percentage of conflicting cases reviewed (i.e. cases where model disagrees with the STS record)
Agreement Rate w/ model: Percentage of reviewed conflicts where the expert reviewer agreed with the AI model prediction rather than the original value from the STS record.

##### 3.6 Excluded Variables

Table S10: Variables excluded from our analysis

| Variable | Section | Collected (%) |
|---|---|---|
| ABG Management During Cooling | Adult Cardiac Anesthesiology | 0.0 |
| ABG Management During Rewarming | Adult Cardiac Anesthesiology | 0.0 |
| Algorithm used to Guide Transfusion | Adult Cardiac Anesthesiology | 0.0 |
| Anesthesiology Care Team Model - Anesthesiologist Directing CRNA Ratio | Adult Cardiac Anesthesiology | 0.0 |
| Anesthesiology Care Team Model - Anesthesiology Directing AA - Ratio | Adult Cardiac Anesthesiology | 0.0 |
| Anesthesiology Total 25% Albumin | Adult Cardiac Anesthesiology | 0.0 |
| Anesthesiology Total 5 % Albumin | Adult Cardiac Anesthesiology | 0.0 |
| Aneurysm - Rupture - Contained | Aorta and Aortic Root Procedures | 0.0 |
| Anticoagulant Bleeding Event - Type | Postoperative Events | 0.117 |
| Antithrombin III Total Dose | Adult Cardiac Anesthesiology | 0.0 |
| Antithrombin III prior to CPB | Adult Cardiac Anesthesiology | 0.0 |
| Aorta Device - Implant Method #04 | Aorta and Aortic Root Procedures | 0.033 |
| Aorta Device - Implant Method #05 | Aorta and Aortic Root Procedures | 0.017 |
| Aorta Device - Implant Method #06 | Aorta and Aortic Root Procedures | 0.0 |
| Aorta Device - Implant Method #07 | Aorta and Aortic Root Procedures | 0.0 |
| Aorta Device - Implant Method #08 | Aorta and Aortic Root Procedures | 0.0 |
| Aorta Device - Implant Method #09 | Aorta and Aortic Root Procedures | 0.0 |
| Aorta Device - Implant Method #10 | Aorta and Aortic Root Procedures | 0.0 |
| Aorta Device - Implant Method #11 | Aorta and Aortic Root Procedures | 0.0 |
| Aorta Device - Implant Method #12 | Aorta and Aortic Root Procedures | 0.0 |
| Aorta Device - Implant Method #13 | Aorta and Aortic Root Procedures | 0.0 |
| Aorta Device - Implant Method #14 | Aorta and Aortic Root Procedures | 0.0 |
| Aorta Device - Implant Method #15 | Aorta and Aortic Root Procedures | 0.0 |
| Aorta Device - Location #05 | Aorta and Aortic Root Procedures | 0.033 |
| Aorta Device - Location #06 | Aorta and Aortic Root Procedures | 0.017 |
| Aorta Device - Location #07 | Aorta and Aortic Root Procedures | 0.0 |
| Aorta Device - Location #08 | Aorta and Aortic Root Procedures | 0.0 |
| Aorta Device - Location #09 | Aorta and Aortic Root Procedures | 0.0 |
| Aorta Device - Location #10 | Aorta and Aortic Root Procedures | 0.0 |
| Aorta Device - Location #11 | Aorta and Aortic Root Procedures | 0.0 |
| Aorta Device - Location #12 | Aorta and Aortic Root Procedures | 0.0 |
| Aorta Device - Location #13 | Aorta and Aortic Root Procedures | 0.0 |
| Aorta Device - Location #14 | Aorta and Aortic Root Procedures | 0.0 |
| Aorta Device - Location #15 | Aorta and Aortic Root Procedures | 0.0 |
| Aorta Device - Model Number #03 | Aorta and Aortic Root Procedures | 0.151 |
| Aorta Device - Model Number #04 | Aorta and Aortic Root Procedures | 0.0 |
| Aorta Device - Model Number #05 | Aorta and Aortic Root Procedures | 0.0 |
| Aorta Device - Model Number #06 | Aorta and Aortic Root Procedures | 0.0 |
| Aorta Device - Model Number #07 | Aorta and Aortic Root Procedures | 0.0 |
| Aorta Device - Model Number #08 | Aorta and Aortic Root Procedures | 0.0 |
| Aorta Device - Model Number #09 | Aorta and Aortic Root Procedures | 0.0 |
| Aorta Device - Model Number #10 | Aorta and Aortic Root Procedures | 0.0 |
| Aorta Device - Model Number #11 | Aorta and Aortic Root Procedures | 0.0 |
| Aorta Device - Model Number #12 | Aorta and Aortic Root Procedures | 0.0 |
| Aorta Device - Model Number #13 | Aorta and Aortic Root Procedures | 0.0 |
| Aorta Device - Model Number #14 | Aorta and Aortic Root Procedures | 0.0 |
| Aorta Device - Model Number #15 | Aorta and Aortic Root Procedures | 0.0 |
| Aorta Device - Outcome #04 | Aorta and Aortic Root Procedures | 0.033 |
| Aorta Device - Outcome #05 | Aorta and Aortic Root Procedures | 0.017 |
| Aorta Device - Outcome #06 | Aorta and Aortic Root Procedures | 0.0 |
| Aorta Device - Outcome #07 | Aorta and Aortic Root Procedures | 0.0 |

Table S10: Variables excluded from our analysis

| Variable | Section | Collected (%) |
|---|---|---|
| Aorta Device - Outcome #08 | Aorta and Aortic Root Procedures | 0.0 |
| Aorta Device - Outcome #09 | Aorta and Aortic Root Procedures | 0.0 |
| Aorta Device - Outcome #10 | Aorta and Aortic Root Procedures | 0.0 |
| Aorta Device - Outcome #11 | Aorta and Aortic Root Procedures | 0.0 |
| Aorta Device - Outcome #12 | Aorta and Aortic Root Procedures | 0.0 |
| Aorta Device - Outcome #13 | Aorta and Aortic Root Procedures | 0.0 |
| Aorta Device - Outcome #14 | Aorta and Aortic Root Procedures | 0.0 |
| Aorta Device - Outcome #15 | Aorta and Aortic Root Procedures | 0.0 |
| Aorta Device - Unique Device Identifier #02 | Aorta and Aortic Root Procedures | 0.117 |
| Aorta Device - Unique Device Identifier #03 | Aorta and Aortic Root Procedures | 0.0 |
| Aorta Device - Unique Device Identifier #04 | Aorta and Aortic Root Procedures | 0.0 |
| Aorta Device - Unique Device Identifier #05 | Aorta and Aortic Root Procedures | 0.0 |
| Aorta Device - Unique Device Identifier #06 | Aorta and Aortic Root Procedures | 0.0 |
| Aorta Device - Unique Device Identifier #07 | Aorta and Aortic Root Procedures | 0.0 |
| Aorta Device - Unique Device Identifier #08 | Aorta and Aortic Root Procedures | 0.0 |
| Aorta Device - Unique Device Identifier #09 | Aorta and Aortic Root Procedures | 0.0 |
| Aorta Device - Unique Device Identifier #10 | Aorta and Aortic Root Procedures | 0.0 |
| Aorta Device - Unique Device Identifier #11 | Aorta and Aortic Root Procedures | 0.0 |
| Aorta Device - Unique Device Identifier #12 | Aorta and Aortic Root Procedures | 0.0 |
| Aorta Device - Unique Device Identifier #13 | Aorta and Aortic Root Procedures | 0.0 |
| Aorta Device - Unique Device Identifier #14 | Aorta and Aortic Root Procedures | 0.0 |
| Aorta Device - Unique Device Identifier #15 | Aorta and Aortic Root Procedures | 0.0 |
| Aorta Presentation - Neuro Deficit | Aorta and Aortic Root Procedures | 0.0 |
| Aortic Arch Atheroma Mobility | Adult Cardiac Anesthesiology | 0.0 |
| Aortic Arch Visualized | Adult Cardiac Anesthesiology | 0.0 |
| Aortic Trauma - Location | Aorta and Aortic Root Procedures | 0.0 |
| Aortic Valve or Aortic Valve Composite Graft Implanted - Unique Device Identifier | Aorta and Aortic Root Procedures | 0.0 |
| Arch Vessel Management - Left Carotid - Extra-anatomic Bypass | Aorta and Aortic Root Procedures | 0.0 |
| Arch Vessel Management - Left Subclavian - Extra-anatomic Bypass | Aorta and Aortic Root Procedures | 0.033 |
| Argatroban | Adult Cardiac Anesthesiology | 0.0 |
| Arterial Outflow Temperature Measured | Adult Cardiac Anesthesiology | 0.0 |
| Ascending Aorta Assessed | Adult Cardiac Anesthesiology | 0.0 |
| Ascending Aortic Atheroma Mobility | Adult Cardiac Anesthesiology | 0.0 |
| Autologous Normovolemic Hemodilution (ANH) | Adult Cardiac Anesthesiology | 0.0 |
| Autologous Normovolemic Hemodilution (ANH) Volume | Adult Cardiac Anesthesiology | 0.0 |
| Bivalirudin | Adult Cardiac Anesthesiology | 0.0 |
| Blood Pressure Baseline (Pre-Anesthetic Induction) - Diastolic | Adult Cardiac Anesthesiology | 0.0 |
| Blood Pressure Baseline (Pre-Anesthetic Induction) - Systolic | Adult Cardiac Anesthesiology | 0.0 |
| CAB 08 | Coronary Bypass | 0.1 |
| CAB 09 | Coronary Bypass | 0.033 |
| CAB 10 | Coronary Bypass | 0.0 |
| CAB Conduit 07 | Coronary Bypass | 0.1 |
| CAB Conduit 08 | Coronary Bypass | 0.033 |
| CAB Conduit 09 | Coronary Bypass | 0.0 |
| CAB Conduit 10 | Coronary Bypass | 0.0 |
| CAB Distal Position 07 | Coronary Bypass | 0.1 |
| CAB Distal Position 08 | Coronary Bypass | 0.033 |
| CAB Distal Position 09 | Coronary Bypass | 0.0 |
| CAB Distal Position 10 | Coronary Bypass | 0.0 |
| CAB Distal Site 07 | Coronary Bypass | 0.1 |

Table S10: Variables excluded from our analysis

| Variable | Section | Collected (%) |
|---|---|---|
| CAB Distal Site 08 | Coronary Bypass | 0.033 |
| CAB Distal Site 09 | Coronary Bypass | 0.0 |
| CAB Distal Site 10 | Coronary Bypass | 0.0 |
| CAB Endarterectomy 07 | Coronary Bypass | 0.1 |
| CAB Endarterectomy 08 | Coronary Bypass | 0.033 |
| CAB Endarterectomy 09 | Coronary Bypass | 0.0 |
| CAB Endarterectomy 10 | Coronary Bypass | 0.0 |
| CAB Proximal Site 07 | Coronary Bypass | 0.1 |
| CAB Proximal Site 08 | Coronary Bypass | 0.033 |
| CAB Proximal Site 09 | Coronary Bypass | 0.0 |
| CAB Proximal Site 10 | Coronary Bypass | 0.0 |
| CPB Utilization - Combination Plan | Operative | 0.134 |
| CPB Utilization - Unplanned Combination Reason | Operative | 0.017 |
| CPT-1 Code # 10 | Operative | 0.0 |
| CPT-1 Code # 6 | Operative | 0.033 |
| CPT-1 Code # 7 | Operative | 0.0 |
| CPT-1 Code # 8 | Operative | 0.0 |
| CPT-1 Code # 9 | Operative | 0.0 |
| Cardiopulmonary Bypass Used | Adult Cardiac Anesthesiology | 0.0 |
| Care Team Model | Adult Cardiac Anesthesiology | 0.0 |
| Cell saver volume | Adult Cardiac Anesthesiology | 0.0 |
| Conversion To Open - Reason | Aorta and Aortic Root Procedures | 0.017 |
| Core Temperature Maximum During Rewarming | Adult Cardiac Anesthesiology | 0.0 |
| Core Temperature Source In OR | Adult Cardiac Anesthesiology | 0.0 |
| Core Temperature Upon Entry To ICU/PACU | Adult Cardiac Anesthesiology | 0.0 |
| Core Temperature Upon Entry To ICU/PACU Measured | Adult Cardiac Anesthesiology | 0.0 |
| Crystalloid given by Anesthesia | Adult Cardiac Anesthesiology | 0.0 |
| Crystalloid given by Anesthesia - Type | Adult Cardiac Anesthesiology | 0.0 |
| Crytalloid Administered by Perfusion Team - Type | Adult Cardiac Anesthesiology | 0.0 |
| Dissection - Malperfusion Type | Aorta and Aortic Root Procedures | 0.05 |
| Dissection - Rupture - Contained | Aorta and Aortic Root Procedures | 0.084 |
| Dissection - Rupture Location | Aorta and Aortic Root Procedures | 0.084 |
| Endoleak - Type I - Leak At Graft Attachment Site | Aorta and Aortic Root Procedures | 0.017 |
| Endoleak - Type I - Location | Aorta and Aortic Root Procedures | 0.017 |
| Endoleak - Type II - Aneurysm Sac Filling Via Branch Vessel | Aorta and Aortic Root Procedures | 0.017 |
| Endoleak - Type II - Number Of Vessels | Aorta and Aortic Root Procedures | 0.0 |
| Endoleak - Type III - Graft Defect Type | Aorta and Aortic Root Procedures | 0.0 |
| Endoleak - Type III - Leak Through Defect In Graft | Aorta and Aortic Root Procedures | 0.017 |
| Endoleak - Type IV - Leak Through Graft Fabric - Porosity | Aorta and Aortic Root Procedures | 0.017 |
| Endoleak - Type V - Endotension-Expansion Aneurysm Sac Without Leak | Aorta and Aortic Root Procedures | 0.017 |
| Endoleak At End Of Procedure - Type | Aorta and Aortic Root Procedures | 0.0 |
| Endovascular Procedures - Distal Landing Zone | Aorta and Aortic Root Procedures | 0.134 |
| Endovascular Procedures - Proximal Landing Zone | Aorta and Aortic Root Procedures | 0.151 |
| Fibrinogen Upon Entry To ICU/PACU | Adult Cardiac Anesthesiology | 0.0 |
| Fibrinogen Upon Entry To ICU/PACU Measured | Adult Cardiac Anesthesiology | 0.0 |
| First Postoperative INR | Adult Cardiac Anesthesiology | 0.0 |
| Five Percent Albumin Given by Anesthesia | Adult Cardiac Anesthesiology | 0.0 |
| Fresh Frozen Plasma prior to CPB | Adult Cardiac Anesthesiology | 0.0 |

Table S10: Variables excluded from our analysis

| Variable | Section | Collected (%) |
|---|---|---|
| Fresh Frozen Plasma prior to CPB - Total Dose | Adult Cardiac Anesthesiology | 0.0 |
| HICN / MBI Number - Secondary | Hospitalization | 0.0 |
| HICN / MBI Number Primary | Hospitalization | 0.0 |
| Heart Rate Baseline (Pre-Anesthetic Induction) | Adult Cardiac Anesthesiology | 0.0 |
| Hematocrit Upon Entry To ICU/PACU | Adult Cardiac Anesthesiology | 0.0 |
| Hematocrit Upon Entry To ICU/PACU Measured | Adult Cardiac Anesthesiology | 0.0 |
| Hemofiltration Volume Removed by Perfusion Team | Adult Cardiac Anesthesiology | 0.0 |
| Hemoglobin Measured upon admission to Post Op Care Location (PACU, ICU) | Adult Cardiac Anesthesiology | 0.0 |
| Hemoglobin Upon Entry To ICU/PACU | Adult Cardiac Anesthesiology | 0.0 |
| Heparin Induced Thrombocytopenia Thrombosis | Postoperative Events | 0.067 |
| Heparin Management | Adult Cardiac Anesthesiology | 0.0 |
| Heparin Prior to CPB | Adult Cardiac Anesthesiology | 0.0 |
| Heparin Total Dose | Adult Cardiac Anesthesiology | 0.0 |
| Highest Arterial Outflow Temperature | Adult Cardiac Anesthesiology | 0.0 |
| Inhaled Vasodilator | Adult Cardiac Anesthesiology | 0.0 |
| Innominate - Extra-Anatomic Bypass Location | Aorta and Aortic Root Procedures | 0.0 |
| Inotropes used to wean from CPB | Adult Cardiac Anesthesiology | 0.0 |
| IntraOp Angiogram - Fluoroscopy Time In Minutes | Aorta and Aortic Root Procedures | 0.017 |
| IntraOp Angiogram - Volume Of Contrast | Aorta and Aortic Root Procedures | 0.084 |
| Intraop Fentanyl | Adult Cardiac Anesthesiology | 0.0 |
| Intraop Fentanyl Dose | Adult Cardiac Anesthesiology | 0.0 |
| Intraop Glucose Trough | Adult Cardiac Anesthesiology | 0.0 |
| Intraop IV Vasodilators Used | Adult Cardiac Anesthesiology | 0.0 |
| Intraop Insulin | Adult Cardiac Anesthesiology | 0.0 |
| Intraop Insulin Total Dose (max units) | Adult Cardiac Anesthesiology | 0.0 |
| Intraop Mgs of Midazolam - Dose | Adult Cardiac Anesthesiology | 0.0 |
| Intraop Midazolam | Adult Cardiac Anesthesiology | 0.0 |
| Intraop Remifentanil Dose | Adult Cardiac Anesthesiology | 0.0 |
| Intraop Reminfentinal | Adult Cardiac Anesthesiology | 0.0 |
| Intraop Sufentanil | Adult Cardiac Anesthesiology | 0.0 |
| Intraop Sufentanil Dose | Adult Cardiac Anesthesiology | 0.0 |
| Intraoperative Glucose Trough Value | Adult Cardiac Anesthesiology | 0.0 |
| Intraoperative Post-Induction/Pre-Incision Transesophageal Echo (TEE) | Adult Cardiac Anesthesiology | 0.0 |
| Intraoperative Post-procedure TEE Performed | Adult Cardiac Anesthesiology | 0.0 |
| Intraoperative Processed EEG (BIS) | Adult Cardiac Anesthesiology | 0.0 |
| LVEF Measured or Estimated | Adult Cardiac Anesthesiology | 0.0 |
| LVEF Percentage | Adult Cardiac Anesthesiology | 0.0 |
| Lactate Upon Entry To ICU/PACU | Adult Cardiac Anesthesiology | 0.0 |
| Lactate Upon Entry To ICU/PACU Measured | Adult Cardiac Anesthesiology | 0.0 |
| Left Atrial Medial-Lateral Size | Adult Cardiac Anesthesiology | 0.0 |
| Left Atrial Size | Adult Cardiac Anesthesiology | 0.0 |
| Left Atrial Superior-Inferior Size | Adult Cardiac Anesthesiology | 0.0 |
| Maximal Aortic Arch Atheroma Thickness | Adult Cardiac Anesthesiology | 0.0 |
| Maximal Ascending Aortic Atheroma Thickness | Adult Cardiac Anesthesiology | 0.0 |
| Maximal Ascending Aortic Diameter | Adult Cardiac Anesthesiology | 0.0 |
| Mitral Regurgitation | Adult Cardiac Anesthesiology | 0.0 |
| Multimodal Analgesics (OR Entry to 24h post OR exit) | Adult Cardiac Anesthesiology | 0.0 |
| Multimodal Analgesics Given (OR Entry to 24h post OR exit) | Adult Cardiac Anesthesiology | 0.0 |
| Non-Infective Surgical Wound Dehiscence (includes non-infective sterile wound) | Postoperative Events | 0.033 |

Table S10: Variables excluded from our analysis

| Variable | Section | Collected (%) |
|---|---|---|
| Other Card- Epicardial Occlusion Device UDI | Atrial Fibrillation Procedures | 0.067 |
| Other Hospital Performs Cardiac Surgery | Hospitalization | 0.0 |
| Pain Score Baseline | Adult Cardiac Anesthesiology | 0.0 |
| Pain Score Hospital Discharge | Adult Cardiac Anesthesiology | 0.0 |
| Pain Score POD #3 | Adult Cardiac Anesthesiology | 0.0 |
| Patent Foramen Ovale | Adult Cardiac Anesthesiology | 0.0 |
| Patient Died in the OR | Adult Cardiac Anesthesiology | 0.0 |
| Patient Participating In STS-Related Clinical Trial - Patient ID | Administrative | 0.0 |
| Patient Transfer to Acute Care Hospital - Date | Discharge/Mortality | 0.0 |
| Platelets Upon Entry To ICU/PACU | Adult Cardiac Anesthesiology | 0.0 |
| Platelets Upon Entry To ICU/PACU Measured | Adult Cardiac Anesthesiology | 0.0 |
| Post Op Other Sedation | Adult Cardiac Anesthesiology | 0.0 |
| Post-Op-Aortic Reintervention-Type | Postoperative Events | 0.067 |
| Post-Op-Mechanical Assist Device Related Events | Postoperative Events | 0.017 |
| Post-Op-Other-Aortic Endoleak Type | Postoperative Events | 0.0 |
| Post-Procedure Left Ventricular Ejection Fraction Estimate | Adult Cardiac Anesthesiology | 0.0 |
| Post-Procedure Left Ventricular Ejection Fraction Measured | Adult Cardiac Anesthesiology | 0.0 |
| Post-Procedure RV Function | Adult Cardiac Anesthesiology | 0.0 |
| Postop Infusion: Propofol | Adult Cardiac Anesthesiology | 0.0 |
| Postoperative Delirium | Adult Cardiac Anesthesiology | 0.0 |
| Postoperative INR Measured | Adult Cardiac Anesthesiology | 0.0 |
| Previous Other Cardiac Intervention 7 | Previous Cardiac Interventions | 0.017 |
| Previous VAD Unique Device Identifier (UDI) | Mechanical Cardiac Assist Devices | 0.033 |
| Prior Aortic Intervention - Disease Progression - Infrarenal Abdominal (Zones 8,9,10,11) | Aorta and Aortic Root Procedures | 0.067 |
| Prior Aortic Intervention - Previous Repair Type - Infrarenal Abdominal (Zones 8,9,10,11) | Aorta and Aortic Root Procedures | 0.067 |
| Prior Aortic Intervention - Previous Repair Type - Suprarenal Abdominal (Zones 6,7) | Aorta and Aortic Root Procedures | 0.134 |
| Prior Aortic Intervention - Repair Failure - Infrarenal Abdominal (Zones 8,9,10,11) | Aorta and Aortic Root Procedures | 0.067 |
| Procedural Sedation | Operative | 0.017 |
| Protamine total dose | Adult Cardiac Anesthesiology | 0.0 |
| Pulmonary Artery Catheter Used | Adult Cardiac Anesthesiology | 0.0 |
| Pulmonic Valve Stenosis Degree | Hemodynamics/Cath/Echo | 0.117 |
| RV Function | Adult Cardiac Anesthesiology | 0.0 |
| Radial Artery Harvest and Preparation Time | Coronary Bypass | 0.0 |
| Readmit Reason - Primary Procedure - Aorta Intervention Indication | Readmission | 0.05 |
| Readmit Reason - Primary Procedure - Aorta Intervention Type | Readmission | 0.05 |
| Reason for return to CPB | Adult Cardiac Anesthesiology | 0.0 |
| Retrograde Autologous Priming of CPB Circuit | Adult Cardiac Anesthesiology | 0.0 |
| Return to CPB Reason - Ventricular Failure Type | Adult Cardiac Anesthesiology | 0.0 |
| Return to CPB for Echo-Related Diagnosis | Adult Cardiac Anesthesiology | 0.0 |
| Robot Use Time Frame | Operative | 0.067 |
| Saphenous Vein Harvest And Preparation Time | Coronary Bypass | 0.033 |
| Second Explant Year of Implant Known | Valve Surgery Explant | 0.033 |
| Second Valve Explant Device Unique Device Identifier (UDI) | Valve Surgery Explant | 0.017 |
| Second Valve Explant Implant Year | Valve Surgery Explant | 0.033 |
| Systolic Anterior Motion of Mitral Valve | Adult Cardiac Anesthesiology | 0.0 |

Table S10: Variables excluded from our analysis

| Variable | Section | Collected (%) |
|---|---|---|
| Temporary Assist Device Used - Position | Mechanical Cardiac Assist Devices | 0.117 |
| Temporary Text Field | | 0.017 |
| Third Explant Year of Implant Known | Valve Surgery Explant | 0.0 |
| Third Valve Explant Device | Valve Surgery Explant | 0.0 |
| Third Valve Explant Device Known | Valve Surgery Explant | 0.017 |
| Third Valve Explant Device Unique Device Identifier (UDI) | Valve Surgery Explant | 0.0 |
| Third Valve Explant Etiology | Valve Surgery Explant | 0.017 |
| Third Valve Explant Implant Year | Valve Surgery Explant | 0.0 |
| Third Valve Explant Type | Valve Surgery Explant | 0.017 |
| Third Valve Prosthesis Explant | Valve Surgery Explant | 0.117 |
| Third Valve Prosthesis Explant Position | Valve Surgery Explant | 0.017 |
| Total 25% Albumin Administered by Perfusion Team | Adult Cardiac Anesthesiology | 0.0 |
| Total 5% Albumin Administered by Perfusion Team | Adult Cardiac Anesthesiology | 0.0 |
| Total Crystalloid Administered by Anesthesia Care Team | Adult Cardiac Anesthesiology | 0.0 |
| Total Crystalloid Administered by Perfusion Team | Adult Cardiac Anesthesiology | 0.0 |
| Tricuspid Valve Stenosis Degree | Hemodynamics/Cath/Echo | 0.05 |
| Twenty-five Percent Albumin given by Anesthesia | Adult Cardiac Anesthesiology | 0.0 |
| Unintentional Rupture Of Dissection Septum - Location | Aorta and Aortic Root Procedures | 0.0 |
| VAD Implant Indication #2 | Mechanical Cardiac Assist Devices | 0.067 |
| VAD Implant Indication #3 | Mechanical Cardiac Assist Devices | 0.017 |
| VAD-Device #2 | Mechanical Cardiac Assist Devices | 0.067 |
| VAD-Device #3 | Mechanical Cardiac Assist Devices | 0.017 |
| VAD-Explant #3 | Mechanical Cardiac Assist Devices | 0.033 |
| VAD-Explant Date | Mechanical Cardiac Assist Devices | 0.084 |
| VAD-Explant Date #2 | Mechanical Cardiac Assist Devices | 0.017 |
| VAD-Explant Date #3 | Mechanical Cardiac Assist Devices | 0.0 |
| VAD-Explant Reason #2 | Mechanical Cardiac Assist Devices | 0.05 |
| VAD-Explant Reason #3 | Mechanical Cardiac Assist Devices | 0.0 |
| VAD-Implant #3 | Mechanical Cardiac Assist Devices | 0.067 |
| VAD-Implant Date #2 | Mechanical Cardiac Assist Devices | 0.067 |
| VAD-Implant Date #3 | Mechanical Cardiac Assist Devices | 0.017 |
| VAD-Implant Timing #2 | Mechanical Cardiac Assist Devices | 0.067 |
| VAD-Implant Timing #3 | Mechanical Cardiac Assist Devices | 0.017 |
| VAD-Implant Type #2 | Mechanical Cardiac Assist Devices | 0.067 |
| VAD-Implant Type #3 | Mechanical Cardiac Assist Devices | 0.017 |
| VAD-Implant Unique Device Identifier (UDI) | Mechanical Cardiac Assist Devices | 0.1 |
| VAD-Implant Unique Device Identifier (UDI) #2 | Mechanical Cardiac Assist Devices | 0.0 |
| VAD-Implant Unique Device Identifier (UDI) #3 | Mechanical Cardiac Assist Devices | 0.0 |
| VD-Mitral Valve Disease - Carpentier Classification - Class I - Type | Hemodynamics/Cath/Echo | 0.117 |
| VS - Mitral Leaflet Clip Number Implanted | Mitral Valve Procedure | 0.0 |
| VS-Aorta - Aortic Annular Enlargement - Technique | Aorta and Aortic Root Procedures | 0.0 |
| VS-Aorta - Aortic Valve Procedure Surgical Prosthetic Valve Intervention (Not Explant of Valve) | Aorta and Aortic Root Procedures | 0.017 |
| VS-Aorta - Transcatheter Valve Replacement Approach | Aorta and Aortic Root Procedures | 0.0 |
| VS-Aortic Valve Procedure Surgical Prosthetic Valve Intervention Type | Aortic Valve without concomitant Aorta Procedure | 0.0 |
| VS-Mitral Valve Procedure - Surgical Prosthetic Valve Intervention (Not Explant of Valve): | Mitral Valve Procedure | 0.0 |

Table S10: Variables excluded from our analysis

| Variable | Section | Collected (%) |
|---|---|---|
| VS-Pulmonic - Surgeon Fashioned Implant Material | Pulmonic Valve Procedure | 0.0 |
| VS-Pulmonic Proc-Imp-Unique Device Identifier | Pulmonic Valve Procedure | 0.084 |
| VS-Transcatheter Valve Replacement Approach | Aortic Valve without concomitant Aorta Procedure | 0.017 |
| VS-Tricuspid Valve Procedure - Surgical Prosthetic Valve Intervention (Not Explant of Valve) | Triscupid Valve Procedure | 0.0 |
| Valve Surgery - Mitral Valve Chordal Transfer Location | Mitral Valve Procedure | 0.0 |
| Vasopressors used to wean from CPB | Adult Cardiac Anesthesiology | 0.0 |
| Visceral Vessel Intervention - Celiac | Aorta and Aortic Root Procedures | 0.05 |
| Visceral Vessel Intervention - Left Renal | Aorta and Aortic Root Procedures | 0.05 |
| Visceral Vessel Intervention - Right Renal | Aorta and Aortic Root Procedures | 0.05 |
| Visceral Vessel Intervention - Superior Mesenteric | Aorta and Aortic Root Procedures | 0.05 |
| Visceral Vessel Management - Celiac - Extra-anatomic Bypass | Aorta and Aortic Root Procedures | 0.0 |
| Visceral Vessel Management - Left Iliac - Extra-anatomic Bypass | Aorta and Aortic Root Procedures | 0.0 |
| Visceral Vessel Management - Left Renal - Extra-anatomic Bypass | Aorta and Aortic Root Procedures | 0.0 |
| Visceral Vessel Management - Other Visceral Vessel(s) Extra-anatomic Bypass - Location | Aorta and Aortic Root Procedures | 0.0 |
| Visceral Vessel Management - Right Iliac - Extra-anatomic Bypass | Aorta and Aortic Root Procedures | 0.0 |
| Visceral Vessel Management - Right Renal - Extra-anatomic Bypass | Aorta and Aortic Root Procedures | 0.0 |
| Visceral Vessel Management - Superior mesenteric - Extra-anatomic Bypass | Aorta and Aortic Root Procedures | 0.0 |
| Viscoelastic Testing Used During Operation | Adult Cardiac Anesthesiology | 0.0 |
| Volatile Agent Used | Adult Cardiac Anesthesiology | 0.0 |
| Volatile Agent(s) Timing | Adult Cardiac Anesthesiology | 0.0 |
| Volatile Agent(s) Used - Type | Adult Cardiac Anesthesiology | 0.0 |
| WBC Upon Entry To ICU/PACU | Adult Cardiac Anesthesiology | 0.0 |
| WBC Upon Entry To ICU/PACU Measured | Adult Cardiac Anesthesiology | 0.0 |

Table shows the 311 variables that were excluded from our analysis due being collected for less than 0.15% of patients.
Section: Section in the data collection form that the variable corresponds to.
Collected (%).: Percentage of patients for which the variable was collected (i.e. had a non-empty value) in MGB.

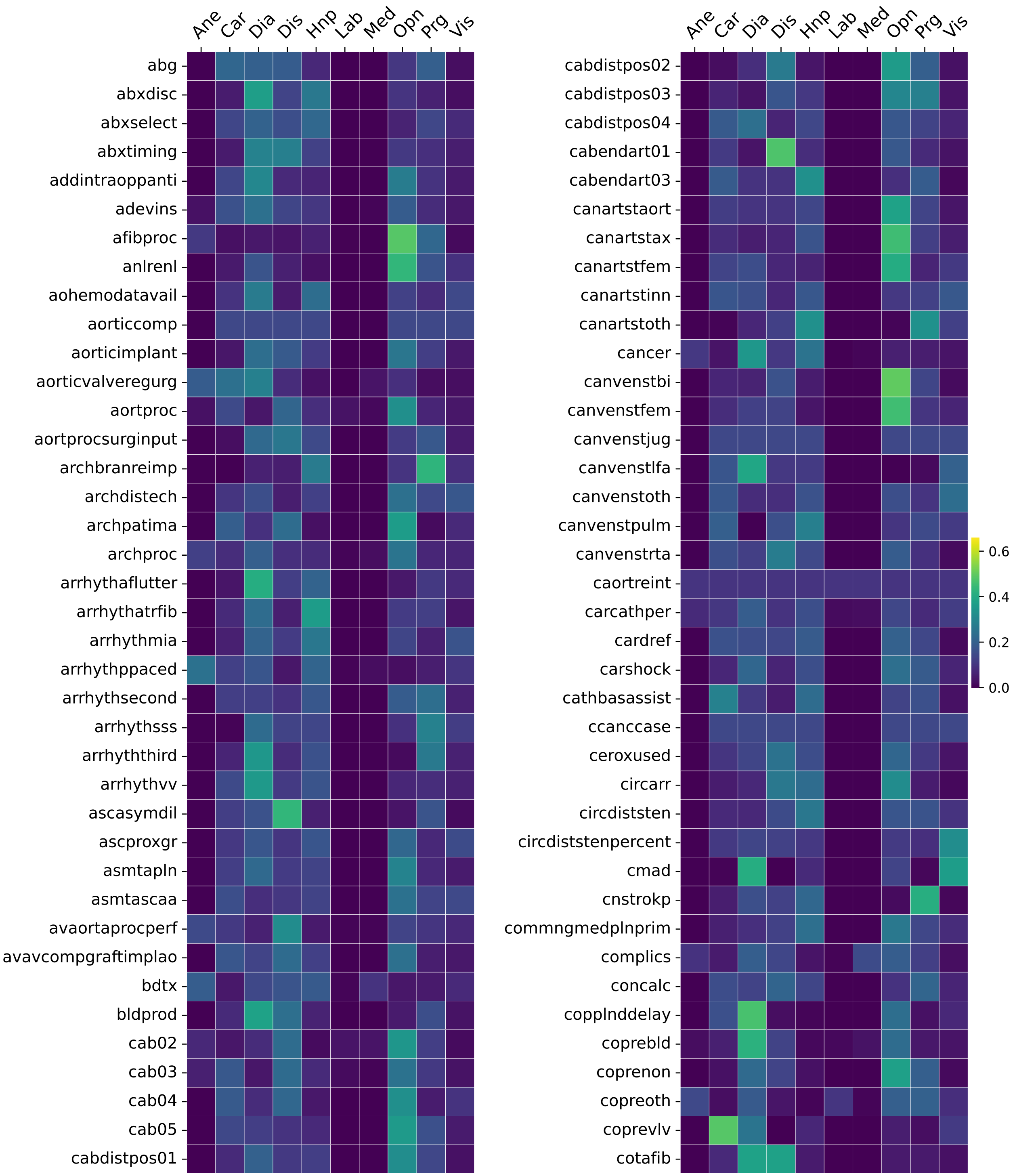


Figure S2: Feature importance of each source on ensemble predictions for AI-derived variables.

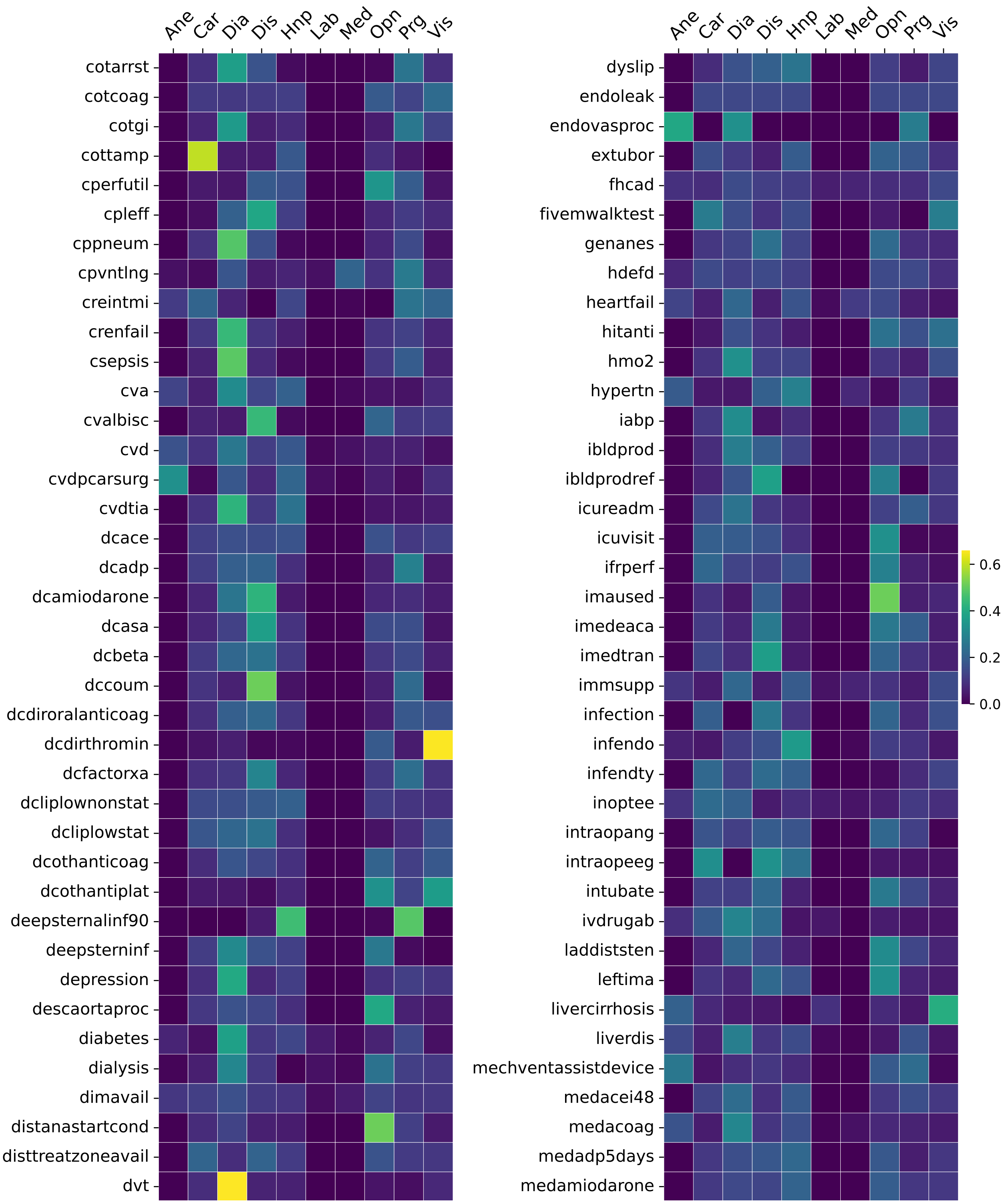


Figure S2: Feature importance of each source on ensemble predictions for AI-derived variables.

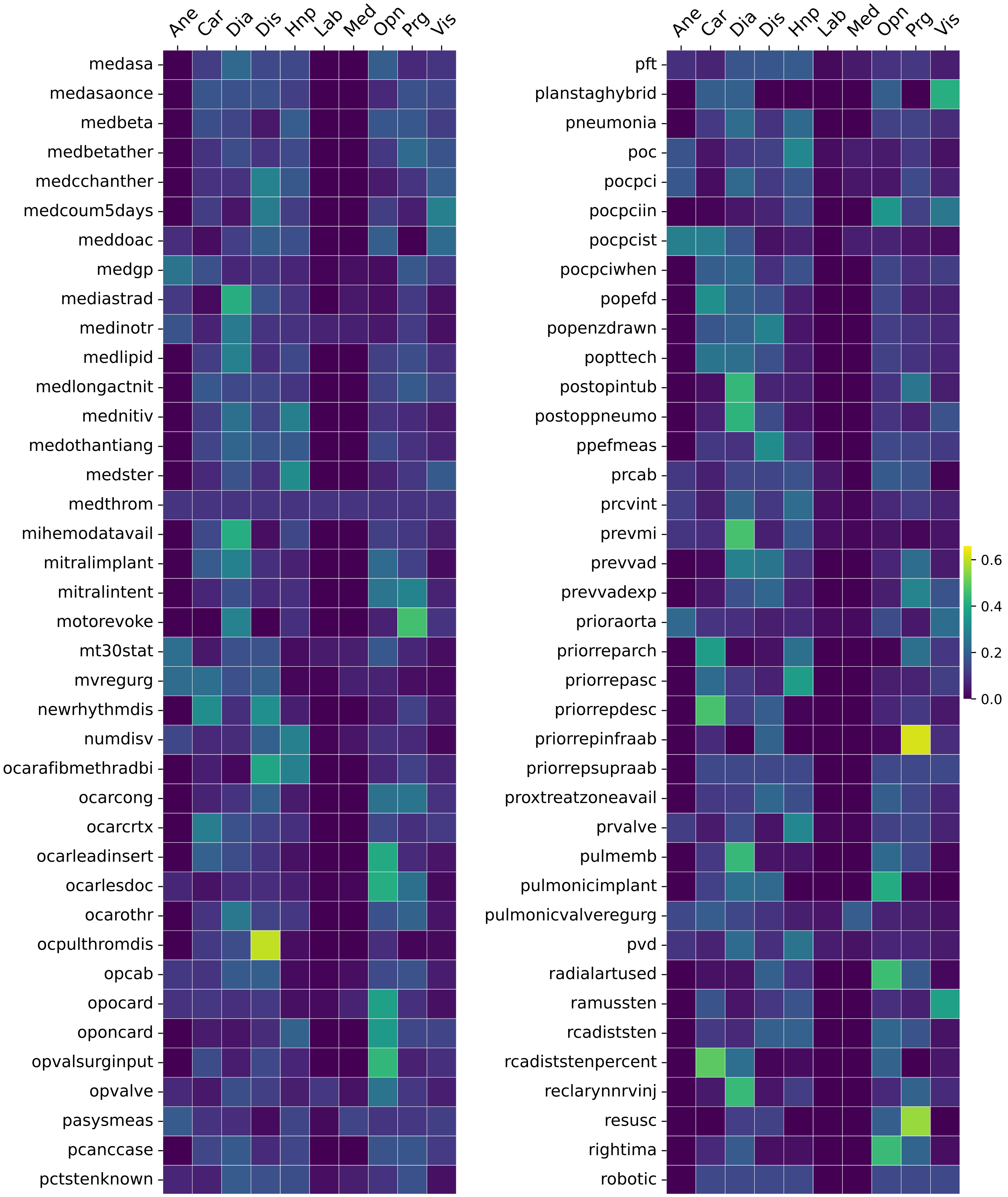


Figure S2: Feature importance of each source on ensemble predictions for AI-derived variables.

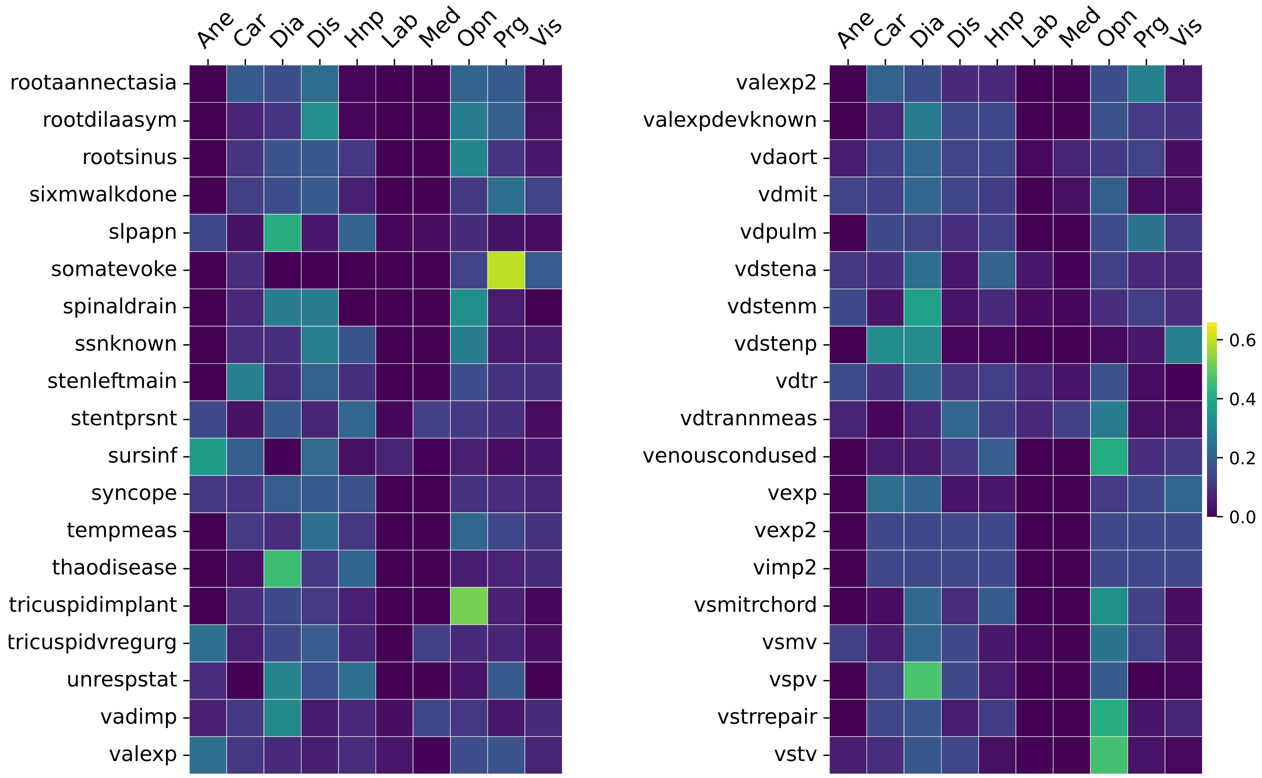


Figure S2: Feature importance of each source on ensemble predictions for AI-derived variables. Heatmap shows normalized absolute values of SHAP feature importance of each source on ensemble predictions. SHAP values from all featurizations of a source were summed to yield a single importance score. Rows represent clinical variables, columns represent prediction sources. Variable abbreviations follow the STS Manual [1]; source abbreviations are: Ane — Anesthesia notes, Car — Cardiology reports, Dia — Diagnoses codes, Dis — Discharge summary, Hnp — History & Physical, Lab — Labs, Med — Medications, Opn — Operative notes, Prg — Progress notes, Vis — Visit notes. Colors indicate feature importance on a common scale. Heatmaps are split across pages for readability. Heatmap was generated based on MGB.

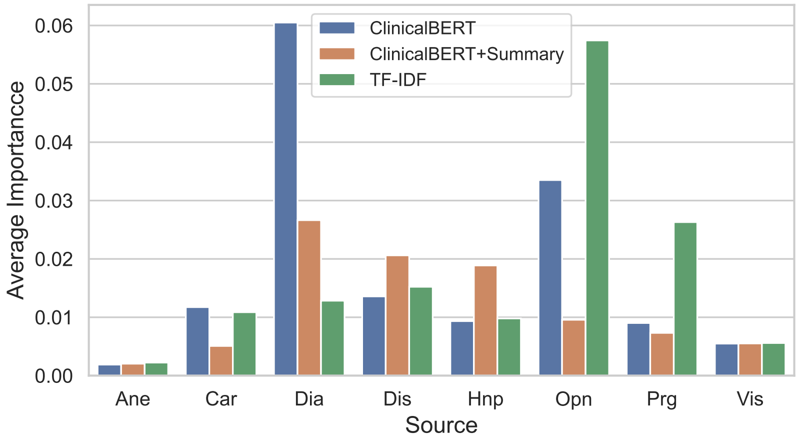


Figure S3: Average importance of unstructured source and featurization method on ensemble prediction. For each variable, importance is calculated by normalizing the absolute SHAP values over sources and featurization methods. Importances are then averaged over all AI-derived variables. Dia represents diagnoses codes that are converted to unstructured text as mentioned in the Methods section. Other source abbreviations are described in Figure S2.

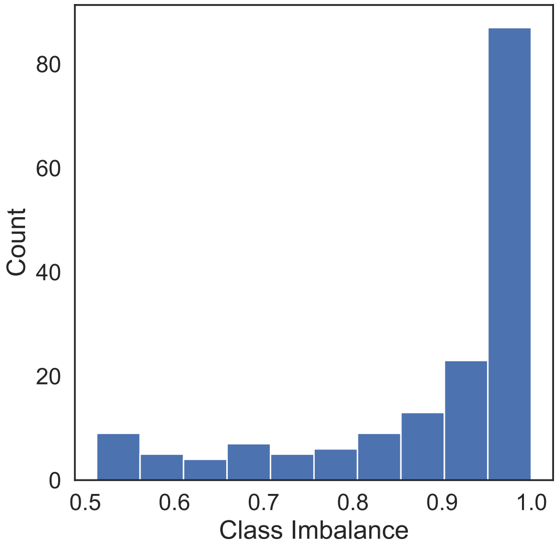


(a) Distribution of class imbalance for AI-derived variables.

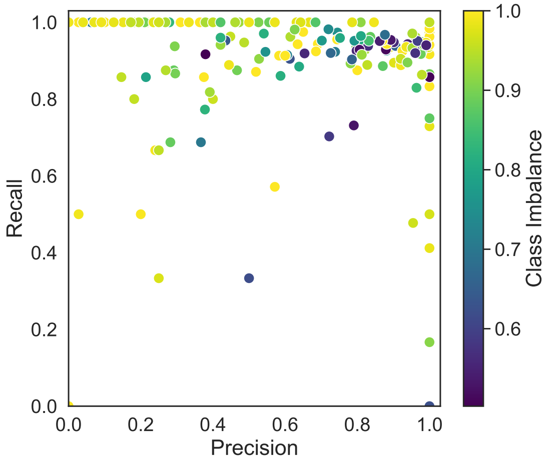


(b) Precision and Recall of AI-derived variables.

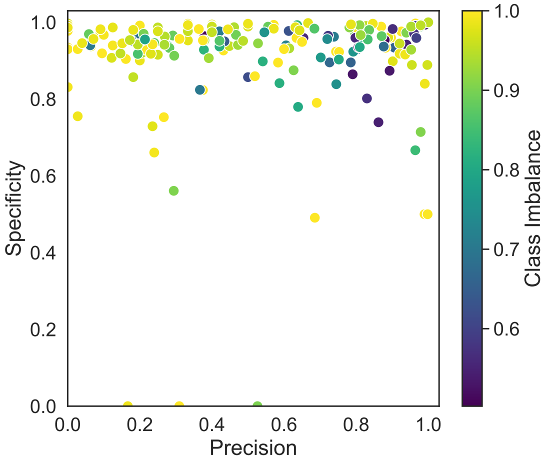


(c) Precision and Specificity of AI-derived variables.

Figure S4: Class imbalance of AI-derived variables and its effect on predictive performance. **Up:** Histogram of class imbalance for the AI-derived variables reported in Table S4. Imbalance is defined as the relative prevalence of the majority class among labeled examples, with higher values indicating greater class imbalance. **Left:** Precision–recall performance of the same AI-derived variables, with points colored by the corresponding class imbalance rate (i.e., majority-class prevalence). **Right:** Precision–specificity of the AI-derived variables, with points colored by the corresponding class imbalance rate. All three plots were generated based on MGB.

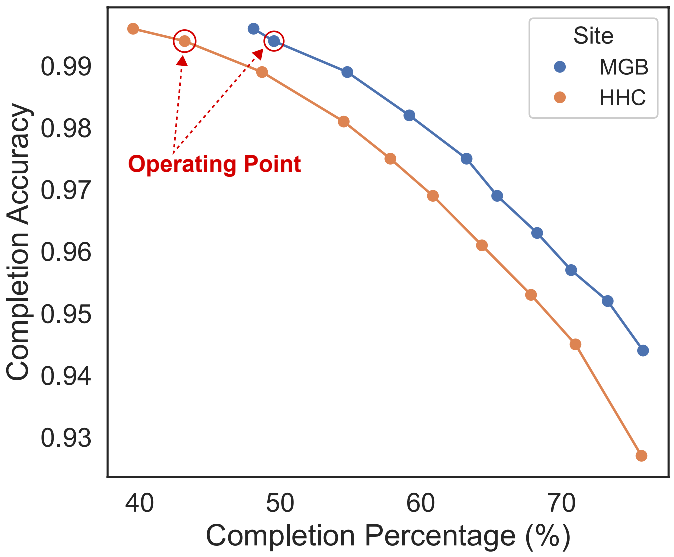


(a) Accuracy–completion tradeoff for the full pipeline.

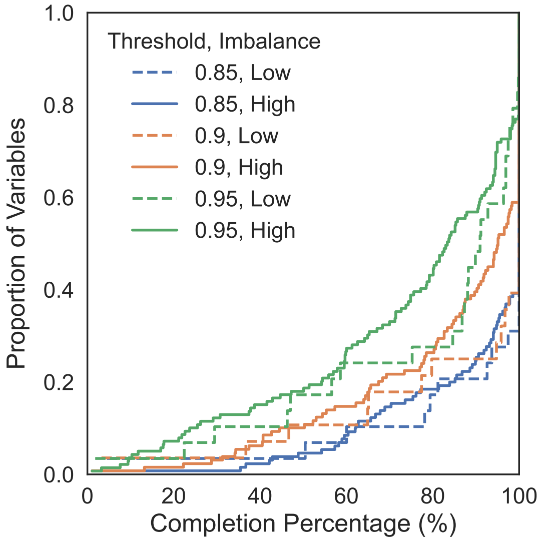


(b) ECDF of AI-derived variable completions stratified by class imbalance and quality control threshold.

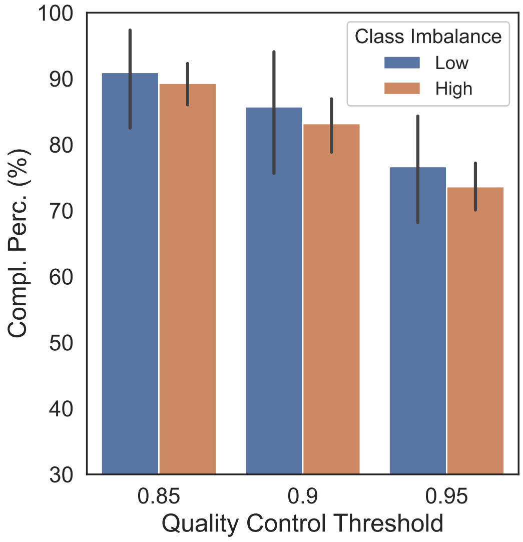


(c) Average completion percentage of AI-derived variables stratified by class imbalance and quality control threshold.

Figure S5: Effect of quality-control thresholds and class imbalance on completion rate. **Up**: Accuracy–completion tradeoff for the full pipeline, obtained by varying quality-control thresholds. The operating point denotes the thresholds used in our analysis, chosen to preserve high accuracy. **Left**: ECDF of completion rates only for the AI-derived variables. For any value of $x$, the ECDF shows the proportion of AI-derived variables with completion rates $\leq x$. Colors indicate different quality-control thresholds. Line styles represent class imbalance: Low imbalance corresponds to minority class prevalence of $25\%$–$50\%$, and High imbalance corresponds to $0\%$–$25\%$. **Right:** Average completion rates of AI-derived variables stratified by class imbalance and quality-control threshold. Both Left and Right plots reflect performance on MGB.

### 4 Additional Result Analysis

#### 4.1 Feature Importance & Ensemble Diversity

To analyze the influence of diverse clinical sources on model predictions, Figure S2 presents the normalized SHAP (SHapley Additive exPlanations) values, detailing the source-specific contributions to the ensemble output for each AI-derived variable [2]. The analysis reveals that the predictive dominance of clinical sources varies considerably across target variables. For instance, the preoperative comorbidity of hypertension ("hypertn") is primarily inferred from the History & Physical reports. Conversely, the abstraction of surgical procedures, such as mitral and tricuspid valve procedures ("vsmv" and "vstv", respectively), depends predominantly on Operative Notes. These findings demonstrate that the ensemble model effectively leverages a diverse set of inputs, dynamically adjusting its reliance based on the clinical context of the target variable rather than overfitting to a single documentation type.

Furthermore, Figure S3 illustrates the comparative impact of various featurization methods on the ensemble predictions. The data indicate that the efficacy of each featurization technique is highly dependent on the documentation source. Specifically, the unsummarized ClinicalBERT model achieves optimal performance when applied to Diagnoses texts. This is likely because these texts, which map directly to ICD-9 and ICD-10 codes, are inherently concise; therefore, summarization may inadvertently discard critical information. In contrast, for clinical documents such as History & Physical reports and Discharge Summaries which are among the longest (see Table 2 in the main manuscript), the application of summarization techniques prior to ClinicalBERT encoding demonstrably enhances predictive performance.

#### 4.2 Performance Metrics

Our pipeline achieves very high values in AUC, recall/sensitivity and specificity for the majority of AI-derived variables (Table S4). In contrast, precision is lower for some variables. This is an expected side-effect of our dual-thresholding strategy. We explicitly select the upper threshold $t_2$ to satisfy a desired level of sensitivity and the lower threshold $t_1$ to satisfy a desired level of specificity. Consequently, because sensitivity and specificity measure accuracy among actual positives and actual negatives respectively, our approach naturally optimizes the correct classification of both classes rather than precision. In addition to that, a large number of variables are imbalanced (Figure S4). Because precision is mathematically defined as the ratio of true positives to total predicted positives:

$$Precision = \frac{TP}{TP + FP}$$

it is highly sensitive to this low prevalence. Hence, even when a model demonstrates near-perfect sensitivity and specificity, the absolute number of false positives can outnumber the exceedingly rare true positives, mathematically driving down the precision score. This effect can clearly be seen in Figure S4, where models with near-perfect sensitivity (recall) or specificity, often exhibit low precision scores, especially under heavy class imbalance.

Furthermore, the operational impact of these errors needs to be contextualized. In direct clinical diagnostics, false positives incur immediate harms, including patient distress, unnecessary interventions, and significant monetary costs. Conversely, in a retrospective clinical registry, false positives and false negatives carry similar, indirect costs. Neither affects immediate patient care; instead, both equally degrade the integrity of downstream applications like hospital quality benchmarking and clinical research. Since the penalty for missing a condition is just as high as incorrectly flagging one, precision and the F1 score, which focus disproportionately on the positive class, are less ideal for our use case. Following guidance from the STS ACSD managers, the more appropriate metrics are sensitivity/recall (accuracy in the positives) and specificity (accuracy in the negatives), as they independently measure performance across both classes. Hence, by optimizing these two metrics, our dual-thresholding approach effectively navigates the class imbalance to maintain the strict accuracy standards required for downstream registry applications.

#### 4.3 Accuracy - Completion Tradeoff

Our dual-thresholding approach inherently balances prediction accuracy against completion rate. Because we strictly prioritize data integrity over full automation to meet the rigorous standards of the STS database, we selected an operating point that maintains an overall accuracy exceeding 99%. Consequently, this stringent requirement naturally caps our automated completion rate at 49.5% for MGB and 43.2% for HHC, leading to many variables being populated with very high accuracy but low completion rates (Table S5). Figure S5a explicitly illustrates this tradeoff: if we were able to tolerate an overall accuracy of 94% instead of >99%, relaxing the quality control thresholds would substantially increase the pipeline's overall completion rate to approximately 75% in MGB.

A primary driver of this disparity between accuracy and completion is the severe class imbalance inherent to many clinical variables. Highly imbalanced variables adversely affect automated completion rates, particularly when stringent performance thresholds are enforced. Figure S5b demonstrates this effect via the empirical cumulative distribution function (ECDF) of AI-derived variables: under strict quality control thresholds (e.g., 0.95), the completion curves for high-imbalance variables (minority class prevalence of 0%–25%) shift notably leftward compared to those with low imbalance (25%–50% prevalence). Similarly, Figure S5c highlights that the gap in average completion percentage between low and high imbalance variables is most pronounced at the highest confidence thresholds. Ultimately, because the models naturally produce fewer high-confidence predictions for rare clinical events, enforcing a near-perfect accuracy standard dictates a higher rate of manual deferral for these specific variables.

##### 4.4 Individual Variable Error Analysis

In Supplementary Sections 4.2 and 4.3, we highlight class imbalance as a primary driver of performance bottlenecks. While the volume of extracted variables precludes exhaustive manual failure analysis across all variables, we conducted a targeted, blinded expert review of discrepancies for two key variables: "history of diabetes" and "CABG". As noted in the main manuscript, expert review overturned the manual ground-truth label in favor of the AI's prediction in 17.9% of flagged diabetes cases and 9.1% of flagged CABG cases.

For true AI misclassifications, our analysis revealed specific error patterns. For diabetes, errors frequently arose from conflicting EHR modalities. For example, clinical notes occasionally documented a history of diabetes while concurrent laboratory results showed normal HbA1c levels, prompting the model to over-rely on textual sources and diagnoses codes over objective lab values (see SHAP Figure S2). Conversely, false negatives occurred when clinical signals were exceptionally weak, such as a single borderline lab value without corroborating documentation in any other source. For CABG procedures, misclassifications were primarily driven by temporal ambiguity; the model occasionally misidentified historical CABG procedures referenced in current clinical notes as having occurred during the index admission. Despite these edge cases, discriminative performance remained exceptionally high for these variables, with AUCs of 0.98 for diabetes and exceeding 0.99 for CABG.

##### 4.5 Excluded Variables

As mentioned in the main manuscript, 311 variables were excluded from our analysis (Figure 2 of the main manuscript) on the grounds of insufficient data and extreme sparsity. These variables were collected for less than 0.15% of patients in MGB (see Table S10 for more details).

The excluded variables predominantly fall into three distinct categories, representing atypical clinical edge cases or administrative overhead rather than core indicators: First, many of these variables seem to capture long-tail procedural accounting for highly complex surgeries, such as sequential anatomical targets beyond a sixth coronary bypass graft or a third ventricular assist device (VAD) explantation during the same surgery. Second, a substantial portion consists of hyper-granular intraoperative and anesthesiology maneuvers, including titrations of specific analgesics (e.g., intraoperative fentanyl dosing) or highly localized visceral vessel interventions that rarely apply to the broader operative cohort. Finally, another set of variables largely reflects supplementary supply-chain tracking—such as Unique Device Identifiers (UDIs) for specific aortic implants—and potentially redundant administrative billing codes, which serve primarily operational and logistical purposes.

Due to their very low collection rates, these variables were deemed unsuitable for automated data abstraction. Even if automated data abstraction were possible for these variables, the corresponding data collection burden reduction would be minimal due to their extreme levels of completion sparsity.
